# Exome-wide rare-variant associations vary across disease trajectories

**DOI:** 10.64898/2026.11.14.26368035

**Authors:** Ying Wang, Weijun Luo, Linfeng Hu, Tony Chen, Wenhan Lu, Connor Dowd, Cindy Wen, Wenjian Bi, Satoshi Koyama, Ida Surakka, Benjamin M. Neale, Mark Daly, Pradeep Natarajan, Alicia R. Martin, Wei Zhou

## Abstract

Genetic studies of chronic disease typically collapse longitudinal clinical histories into binary case-control outcomes, obscuring disease timing and differences in genetic association patterns across disease transitions. Time-to-event analysis can preserve this information, but rare-variant analysis of post-diagnosis progression is challenged by sparse carrier-events and extensive censoring. We developed GATE-GENE, a scalable gene-based framework that maintains calibration under these conditions. Across 300 onset phenotypes in UK Biobank (UKB), GATE-GENE identified 107 exome-wide significant germline associations involving 89 genes and 67 diseases. Across 95 multi-state trajectories comprising baseline (T0), an upstream disease state (T1) and a downstream disease state (T2), disease onset (T0→T1) and subsequent progression (T1→T2) showed limited detectable sharing of rare-variant association signals. Progression showed greater concordance of gene-level association signals with downstream phenotypes (T0→T2) when the analyses shared T2 events occurring after T1. Removing overlapping downstream events reduced the median correlation between gene-level association signals from 0.063 to 0.001, after accounting for background correlation. Null simulations reproduced the relationship between event overlap and concordance without genetic effects, whereas additional simulations showed that near-zero concordance could persist despite extensive underlying causal-gene sharing. This attenuation was reproduced across three ancestry groups in All of Us. Cross-biobank meta-analysis identified 12 additional exome-wide significant progression associations beyond three UKB discoveries, although clinical sensitivity analyses suggested that the strongest *PKD1* signals largely reflected underlying cystic kidney disease. Together, these findings establish GATE-GENE for investigating rare-variant associations with disease timing and progression, while demonstrating how overlapping clinical events and limited carrier-event information complicate inference about genetic sharing across disease transitions.

## Introduction

Many chronic diseases unfold through clinically distinct states and transitions that differ in biology, prognosis and treatment response^1,2^. Yet human genetic studies typically reduce these longitudinal histories to binary case-control outcomes, obscuring both when disease occurs and the sequence of disease states through which downstream outcomes are reached. Genetic associations with disease susceptibility can differ from those with subsequent progression, prognosis or survival^3–5^, suggesting that the genetics of becoming affected may not fully describe the genetics of later disease course. Longitudinal analyses that resolve successive clinical transitions may therefore reveal genetic relationships obscured by conventional case-control phenotypes.

Population-scale biobanks linking genomic data to longitudinal electronic health records (EHRs) now make such analyses possible at scale^6^. Time-to-event (TTE) models incorporate event timing and follow-up directly, preserving information lost when disease is reduced to binary case status^7–10^. Large-scale exome studies have established rare coding variation as a rich source of gene-level disease associations, but major phenome-wide resources have largely analysed disease status rather than longitudinal disease transitions^11,12^. Rare-variant TTE analyses additionally require gene-based aggregation and remain challenging when carrier-event counts are sparse and censoring is extreme, leaving few observed events among variant carriers. Gene-based rare-variant TTE methods such as GATE-STAAR^13^ have recently extended association testing to biobank scale, but calibration under more extreme censoring typical of post-diagnosis progression remains less well characterized, particularly because progression risk sets are substantially smaller than those for disease onset.

A further challenge concerns how longitudinal phenotypes are defined. A multi-state trajectory can be represented by a baseline state (T0), an upstream disease state (T1) and a downstream state (T2). Disease onset is represented by the T0→T1 transition, whereas progression follows individuals from T1 to T2 (T1→T2). A marginal downstream analysis follows individuals from T0 to T2 (marginal T0→T2) irrespective of whether T1 occurs first and can therefore include T2 events occurring after T1 that are also represented in the progression analysis. By contrast, a direct downstream analysis (direct T0→T2) considers only T2 events occurring before T1. Apparent concordance between progression and marginal downstream association signals may consequently arise from overlap in T2 events as well as from shared genetic effects. Multi-state models distinguish direct transitions from those occurring through an upstream state^14,15^, but whether and to what extent overlap in T2 events systematically influences genetic association patterns across disease trajectories remains unclear.

To characterize rare-variant associations across disease onset and progression, we developed GATE-GENE, a scalable gene-based rare-variant TTE framework for highly censored outcomes. We applied GATE-GENE in UK Biobank (UKB) to construct a phenome-wide atlas of rare coding associations with disease onset and to analyse 95 multi-state disease trajectories spanning onset, progression and marginal downstream risk and direct downstream transitions. We quantified gene-level association concordance across transitions and investigated how overlapping downstream events and clinical-route definitions influence observed association patterns, while examining how limited progression power constrains inference about underlying genetic sharing. We then evaluated the reproducibility of these patterns across European (EUR), African (AFR) and Admixed-American (AMR) ancestry groups in the All of Us Research Program (AoU) and performed cross-biobank meta-analysis to increase power for progression discovery. Together, these analyses show that overlapping downstream events can strongly influence observed rare-variant association concordance, while limited progression power constrains inference about underlying genetic sharing across disease trajectories.

## Results

### Study design and analytical framework

We studied rare coding variant associations with disease onset and progression in UKB and AoU using harmonized TTE phenotypes (**Fig. 1 ; Methods**). The study comprised two complementary components: a phenome-wide analysis of disease onset in UKB and a multi-state trajectory analysis in UKB and AoU. UKB whole-exome sequencing (WES) supported the primary discovery analyses, using a larger WES release and HES-restricted analytical population for the onset atlas and the interim approximately 200,000-participant WES release, without an HES-encounter restriction on controls, for multi-state trajectories, while AoU provided whole-genome sequencing data for cross-biobank analyses.

**Fig. 1.**
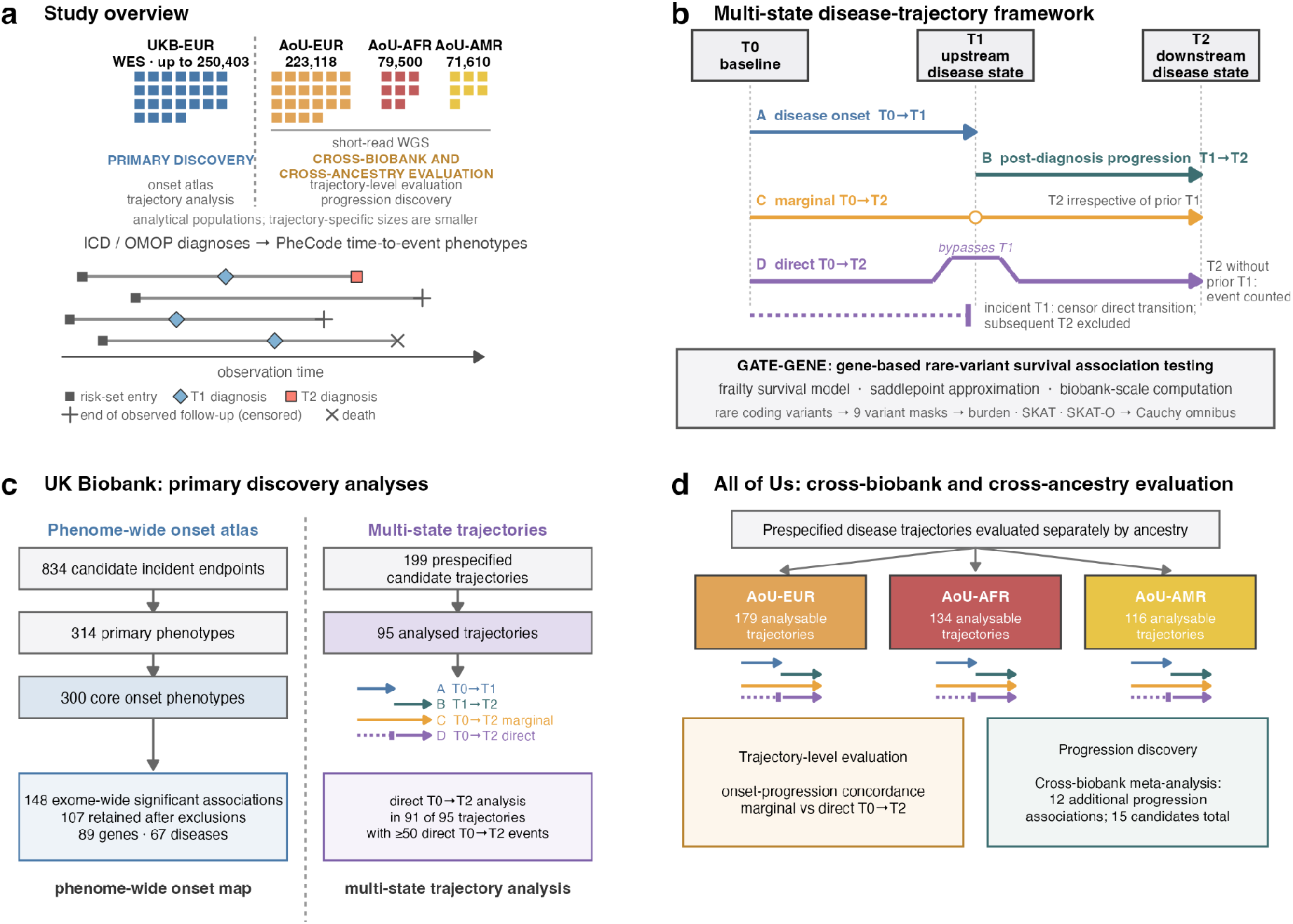
Study design and analytical framework of GATE-GENE a, Overview of the UK Biobank (UKB) and All of Us Research Program (AoU) cohorts used for rare-variant time-to-event analyses. UKB participants of European ancestry (EUR) with whole-exome sequencing (WES) served as the primary discovery cohort, whereas AoU participants with short-read whole-genome sequencing (WGS) were analysed separately in EUR, African (AFR) and Admixed American (AMR) ancestry groups. Longitudinal diagnoses were mapped to PheCode-defined phenotypes to construct disease-onset and progression outcomes. Sample sizes indicate the maximum analytical populations; trajectory-specific sample sizes were smaller. The timelines illustrate risk-set entry, disease diagnoses, censoring and death. **b,** Four complementary transitions were defined for each disease trajectory: disease onset (A, T0→T1), post-diagnosis progression (B, T1→T2), marginal downstream risk (C, T0→T2, irrespective of prior T1) and direct downstream risk (D, T0→T2, with follow-up censored at incident T1). Thus, the direct transition excludes T2 events occurring after T1 that contribute to both progression and marginal downstream analyses. GATE-GENE performs gene-based rare-variant survival association testing using a frailty model and saddlepoint approximation, with burden, SKAT and SKAT-O tests across nine prespecified variant masks and Cauchy combination across masks. **c,** UKB primary discovery comprised a phenome-wide disease-onset atlas and multistate trajectory analyses. Separately, 95 of 199 prespecified disease trajectories met the primary eligibility criteria; 91 had at least 50 direct T0→T2 events for the paired transition comparison. **d,** Prespecified disease trajectories were evaluated separately in AoU-EUR, AoU-AFR and AoU-AMR, yielding 179, 134 and 116 analysable trajectories, respectively. Trajectory-level analyses assessed association concordance between disease transitions across ancestry groups. Cross-biobank meta-analysis combining UKB and AoU identified 12 additional exome-wide significant progression associations beyond the three identified in UKB, yielding 15 statistical candidates in total.

For the onset atlas, we began with 834 PheCode-defined incident endpoints with ≥200 events in UKB-EUR. After excluding non-disease, administrative, and redundant endpoints and separating phenotypes for which first recorded diagnosis was an unreliable proxy for onset, we retained 314 primary phenotypes. Of these, 14 were classified as timing-limited because first recorded diagnosis in this middle-aged to older cohort may not adequately represent biological onset, leaving 300 core phenotypes for the primary analysis, with a median of 238,296 participants per phenotype (**Supplementary Table 1).**

For the trajectory analysis, we prespecified 199 candidate multi-state trajectories independently of genetic association results. These were drawn primarily from directional disease associations identified in population-scale registry studies and supplemented with clinically established disease sequences supported by epidemiological evidence^16–18^. We further filtered candidate trajectories to those with all three primary transition analyses available–upstream disease onset (T0→T1), post-diagnosis progression (T1→T2) and marginal downstream risk (T0→T2)–and at least 100 progression events. Applying these criteria in UKB-EUR yielded 95 trajectories, with a median progression risk-set size of 4,234 participants (**Supplementary Table 2**). A median of 10.6% of marginal T0→T2 events (IQR, 6.8-14.2%) occurred after incident T1. To investigate the influence of overlapping downstream events on association concordance, we additionally constructed cause-specific direct T0→T2 transitions, in which follow-up ended at incident T1. Of the 95 trajectories, 91 had at least 50 direct T0→T2 events and were retained for paired concordance analyses.

Applying the same eligibility criteria in AoU yielded 179, 134 and 116 analysable trajectories in EUR, AFR and AMR, respectively (**Supplementary Table 3**). Corresponding direct T0→T2 transitions were also constructed in each ancestry group. AoU enabled cross-ancestry evaluation of trajectory-level association patterns and was combined with UKB in cross-biobank meta-analysis to increase power for progression discovery. Across cohorts, T1→T2 analyses contained substantially less event information than T0→T1 and marginal T0→T2 analyses, with smaller risk sets, sparse carrier-event counts and high censoring.

### GATE-GENE remains calibrated under extreme censoring

Building on recent advances in gene-based rare-variant TTE testing, we developed GATE-GENE for scalable association analysis under highly censored biobank settings. GATE-GENE uses a mixed-effects frailty model^19^ and applies saddlepoint approximation (SPA)^20,21^ to sparse single-variant score statistics before gene-based aggregation, improving type I error control when carrier-event counts are limited (**Methods; Supplementary Note 1; Fig. 1).** We evaluated burden, SKAT^22^ and SKAT-O^23^ tests across nine prespecified rare-coding masks defined by three MAF cutoffs (1%, 0.1% and 0.01%) and three variant annotation categories (LoF only, LoF plus missense and LoF plus missense plus synonymous) and combined evidence across masks using the Cauchy method^24^.

We evaluated calibration using UKB exome genotypes from 100,000 White British participants under null phenotypes with 90%, 95% and 99% censoring (**Supplementary Note 2; Supplementary Figs. 1** **and 2; Supplementary Table 4**). GATE-GENE showed no statistically significant evidence of type I error inflation at the exome-wide threshold across the evaluated censoring levels. At the *α* = 2.5 × 10⁻⁶, pooled empirical type I error was 1.4 × 10⁻⁶ for both burden and SKAT (95% CI: 5.3 × 10^-7^ – 3.1 × 10^-6^), and 2.9 × 10⁻⁶ for SKAT-O (95% CI: 1.5 × 10^-6^ – 5.0 × 10^-6^), and 2.1 × 10⁻⁶ (95% CI: 9.9 × 10^-7^ – 4.1 × 10^-6^) for the per-gene Cauchy omnibus test. At the less stringent threshold of *α* = 0.001, residual inflation remained, with observed-to-expected exceedance ratios of 1.11-1.55 at. Without SPA, however, SKAT and SKAT-O became increasingly inflated at 95% and 99% censoring, whereas burden remained approximately calibrated.

**Fig. 2.**
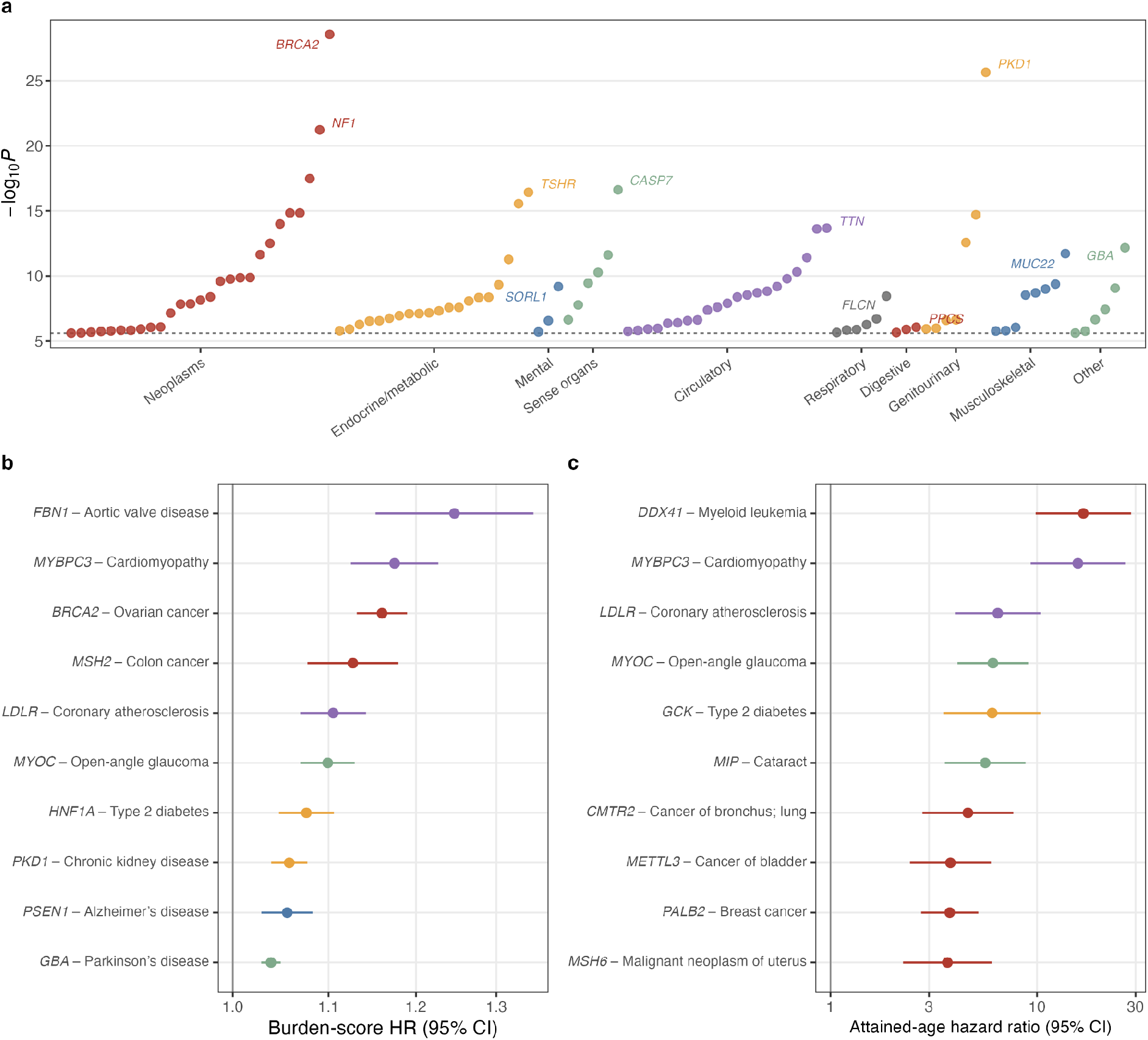
Phenome-wide rare coding associations with disease onset and timing in UK Biobank a, Phenome-wide landscape of the 107 germline-focused exome-wide significant gene-phenotype associations identified across 300 core incident-onset phenotypes in UKB participants of European ancestry. Points show gene-level Cauchy-combined association evidence and are grouped and coloured by clinical domain; selected genes are labelled. The dashed line denotes the exome-wide significance threshold (*P* = 2.5 × 10⁻⁶). **b,** Representative associations across clinical domains. Points show hazard ratios (HRs) per unit of the weighted burden score for the selected burden mask, with the horizontal lines indicating 95% confidence intervals. The vertical line denotes HR = 1. **c,** Carrier-based analyses of disease timing for selected associations with sufficient carrier-event counts. Points show HRs from Cox models using attained age as the time scale with delayed entry at cohort enrolment; horizontal lines indicate 95% confidence intervals. HR > 1 indicates a higher hazard of recorded diagnosis at a given attained age among rare-variant carriers.

For comparison, we evaluated GATE-STAAR^13^ using null phenotypes generated under the same simulation framework, with the comparison restricted to chromosome 19 (**Supplementary Fig. 3**). Although GATE-STAAR was simulation-evaluated at censoring levels up to 90%, its SKAT and STAAR-O tests showed substantial type I error inflation at 95% and 99% censoring in our simulations. At 99% censoring and *α* = 2.5 × 10⁻⁶, empirical type I error reached 1.7 × 10⁻^4^ for SKAT and 8.1 × 10⁻^5^ for STAAR-O, corresponding to 69.4-fold and 32.3-fold inflation, respectively, whereas the burden test remained near nominal at 3.0 × 10⁻^6^. A similar pattern was observed at *α* = 0.001.

Under simulated alternatives, GATE-GENE also gained modest but consistent power over matched incident case-control analysis using SAIGE-GENE+^24,25^. At the exome-wide threshold, SKAT-O power was 24.0%, 58.5% and 66.6% across the three causal architectures, compared with 22.1%, 54.9% and 61.5% for SAIGE-GENE+, respectively (paired *P* ≤ 1.2 × 10^-6^ in all scenarios; **Supplementary Fig. 4; Supplementary Tables 5 and 6**). Together, these results support the calibration of GATE-GENE at exome-wide significance thresholds under extreme censoring, with modest gains in power when event timing provides additional information.

### Phenome-wide mapping captures rare coding associations with disease timing in UK Biobank

Across 300 core UKB-EUR incident-onset phenotypes, GATE-GENE identified 148 exome-wide significant gene-phenotype associations at *P* < 2.5 × 10⁻⁶ per phenotype (**Supplementary Note 3**). After excluding associations attributable to clonal haematopoiesis, somatic neoplasia, extended-MHC effects or other somatic confounding, 107 germline-focused associations remained, involving 89 genes and 67 phenotypes (**Fig. 2a ; Supplementary Table 7**). Of these, 83 (78%) also met the study-wide FDR criterion across all gene-phenotype tests. Requiring the most informative burden mask to have MAC ≥20 retained 105 of 107 associations. Across the onset analyses, burden tests showed little inflation, with modest residual inflation for SKAT and SKAT-O concentrated among the most highly censored phenotypes (**Extended Data Fig. 1**).

The atlas recapitulated established associations including *BRCA2* with ovarian cancer, *PKD1* with chronic kidney disease (CKD), *MYBPC3* with primary cardiomyopathy, *GBA* with Parkinson’s disease and *MYOC* with primary open-angle glaucoma (**Fig. 2b)**. It also identified biologically plausible associations such as *TMTC2* with bronchiectasis and *SPP1* with bacterial pneumonia; *TMTC2* is involved in endoplasmic-reticulum calcium homeostasis and cadherin glycosylation, whereas *SPP1* has established roles in pulmonary antibacterial host defense^26,27^. Most associated genes were phenotype-specific, whereas a smaller subset showed pleiotropic association profiles concentrated within related clinical domains (**Supplementary Fig. 5, Supplementary Table 8**).

Gene-set enrichment analyses identified biological programmes enriched among rare-variant onset-associated genes, including DNA repair (Reactome; OR = 7.5, FDR = 3.8 × 10^-3^) and apoptosis (Hallmark; OR = 8.9, FDR = 5.1 × 10^-3^; **Supplementary Note 4**). Domain-specific analyses highlighted DNA repair and apoptosis in neoplasms, cardiac muscle contraction and coagulation in circulatory phenotypes, and ciliary assembly and intraflagellar trafficking in genitourinary phenotypes (**Extended Data Fig. 2)**. These enrichments remained significant after correction across the primary gene lists and pathway collections and in covariate-matched permutation analyses. However, only the neoplasm DNA-repair enrichments satisfied the prespecified leave-one-gene-out robustness criterion; cardiac and ciliary enrichments were sensitive to removal of *MYBPC3* and *PKD1*, respectively (**Supplementary Figs. 6 and 7 ; Supplementary Tables 9-14**).

We also examined convergence between rare- and common-variant associations. Rare-significant genes showed approximately 15-fold enrichment within 250 kb of phenotype-matched common-variant loci. To assess whether these signals persisted independently of nearby common-variant signals, we performed conditional analyses adjusting for common lead variants. Of 90 associations with stable conditional estimates, 81 remained exome-wide significant (**Extended Data Fig. 3; Supplementary Figs. 8 -10 ; Supplementary Tables 15-18; and 5; Supplementary Notes 4 and 5**).

We next evaluated the consistency of GATE-GENE onset findings with case-control analyses under different phenotype definitions. Matched SAIGE-GENE+ incident case-control analyses across 120 phenotypes showed highly concordant gene-level association evidence: the median Spearman correlation of −log10*P* was 0.968 (IQR, 0.944-0.979), median top-50 gene overlap was 43 (IQR, 40-45), and 87 of 98 exome-wide associations were shared (**Supplementary Figs. 11 and 12**). Thus, when incident phenotype definitions were aligned, TTE and case-control analyses recovered largely similar gene-level association signals. Comparison with GeneBass^11^, a UKB exome-based resource using case-control phenotypes that include prevalent and incident cases, provided additional cross-analysis support. In 74 of 99 matched phenotypes, top 200 genes ranked by GATE-GENE showed stronger GeneBass association evidence than the remaining genes after FDR correction (**Supplementary Note 3; Supplementary Figs. 13 and 14**).

Carrier-based survival analyses further characterized disease timing among rare-variant carriers. Among 80 associations with sufficient carrier-event counts for delayed-entry attained-age analysis, 57 remained significant after FDR correction and all showed higher age-specific incidence among rare-variant carriers (**Fig. 2c ; Extended Data Fig. 4; Supplementary Fig. 15**).

### Disease onset and progression show limited detectable rare-variant association sharing

Across 95 trajectories in UKB-EUR, progression (T1→T2) contained substantially fewer events than upstream onset (T0→T1) or marginal downstream (T0→T2) analyses, with a median of 243 T1→T2 events (IQR, 150-429), compared with 4,595 T0→T1 and 3,676 marginal T0→T2 events. Exome-wide discovery was correspondingly limited, particularly for progression. We identified 37, 20 and 3 significant gene-transition associations for T0→T1, marginal T0→T2, and T1→T2, respectively. Progression analyses had a median tail-based genomic control factor of *λ*_0.01_ = 1.05 (**Supplementary Fig. 16; Supplementary Table 19**).

To characterize the clinical relationship between each pair of disease states, we first tested the associations between incident T1 and subsequent T2 risk. Across 79 of 95 trajectories with estimable HRs, incident T1 was associated with a median 7.2-fold higher hazard of T2 (IQR, 4.2-16.4). This strong clinical relationship, however, did not translate to detectable sharing of gene-level association signals between disease onset and progression. Across 95 trajectories, the median Spearman correlation (*ρ*) of gene-level −log_10_*P* values between the two transitions was −0.001 (IQR, −0.009 to 0.009). Moreover, no gene-level association at *P* < 1 × 10⁻⁴ was shared between onset and progression or across all three transitions (**Supplementary Table 20**). Limited detectable sharing also extended beyond three-state trajectories. Across 155 pairs of adjacent transitions assembled from longer clinical chains, gene-level concordance was similarly low (median Spearman *ρ* = 0.005; IQR, −0.002 to 0.011) and no greater than that between disjoint transitions sharing no disease state.

We next investigated factors contributing to the low observed onset-progression concordance (**Extended Data Figs. 5 and 6**). After harmonizing the genes and variant masks tested in each transition, we compared observed concordance with empirical null distributions matched for event counts and available tests. Concordance increased with the number of progression events (Spearman *ρ* = 0.52, *P* = 2 × 10⁻⁷), consistent with greater power to detect shared association signals in larger progression risk sets. By contrast, concordance was not associated with the proportion of individuals with T1 who subsequently developed T2 (Spearman *ρ* = −0.15, *P* = 0.17). Analyses of burden-effect estimates, effect directions and overlap among top-ranked genes showed similar patterns.

To further assess how limited progression power affected the detectability of shared genetic effects, we performed simulations matched to the observed event counts and rare-variant carrier spectrum (**Extended Data Fig. 7; Supplementary Notes 6 and 7; Supplementary Table 21**). When 25% of progression-associated causal genes were also causal for onset, concordance as low as that observed occurred in 22% of simulations, compared with 1.1% when 50% were shared. These findings were conditional on the simulated effect sizes and genetic architectures and did not directly estimate the extent of underlying genetic sharing between onset and progression.

Against this background of low aggregate concordance, three progression associations reached exome-wide significance in UKB: *GSDME/DFNA5* with benign prostatic hyperplasia→urinary tract infection, *MARCHF7/MARCH7* with CKD→hyperkalaemia, and *F13A1* with coronary artery disease (CAD)→myocardial infarction (MI). None showed evidence of association with the corresponding T0→T1 transition (*P* > 0.36 for all), although differences in statistical power preclude interpreting this as definitive evidence of progression specificity. For *GSDME* and *MARCHF7*, progression burden-score HRs were >1 with 95% CIs excluding 1, whereas the corresponding T0→T1 CIs included 1. *F13A1*, which encodes the catalytic subunit of coagulation factor XIII involved in fibrin clot stabilization, emerged as a biologically plausible candidate for CAD→MI progression. However, this association was driven by variance-component SKAT tests and therefore did not establish a directional effect (**Extended Data Fig. 8**). The small number of progression discoveries was consistent with the limited carrier information in progression risk sets: only 39% of progression masks had a cumulative MAC ≥50, compared with 86% of onset masks.

We next investigated why progression showed greater concordance with marginal downstream risk (T0→T2) than with upstream onset (T0→T1), focusing on the contribution of overlapping T2 events to observed gene-level concordance.

### T2-event overlap shapes progression-downstream concordance

#### Progression-downstream concordance tracks overlap in T2 events

Although onset and progression showed little detectable sharing of gene-level association signals, progression (T1→T2) showed greater concordance with marginal downstream risk (T0→T2) than with onset (T0→T1). Gene-level concordance between T1→T2 and marginal T0→T2 was modest (median *ρ* of 0.064; IQR, 0.044-0.094; **Extended Data Fig. 6**), but exceeded the near-zero concordance between T0→T1 and T1→T2 in all 95 trajectories (paired two-sided Wilcoxon signed-rank test, *P* = 2.7 × 10^-17^; **Fig. 3a**). Progression-downstream concordance increased with the number of progression events (Spearman *ρ* = 0.39-0.45), consistent with greater power in larger progression risk sets. Nevertheless, progression remained more concordant with marginal downstream risk than with onset within event-count strata, suggesting that differences in event counts alone could not explain the greater concordance.

**Fig. 3.**
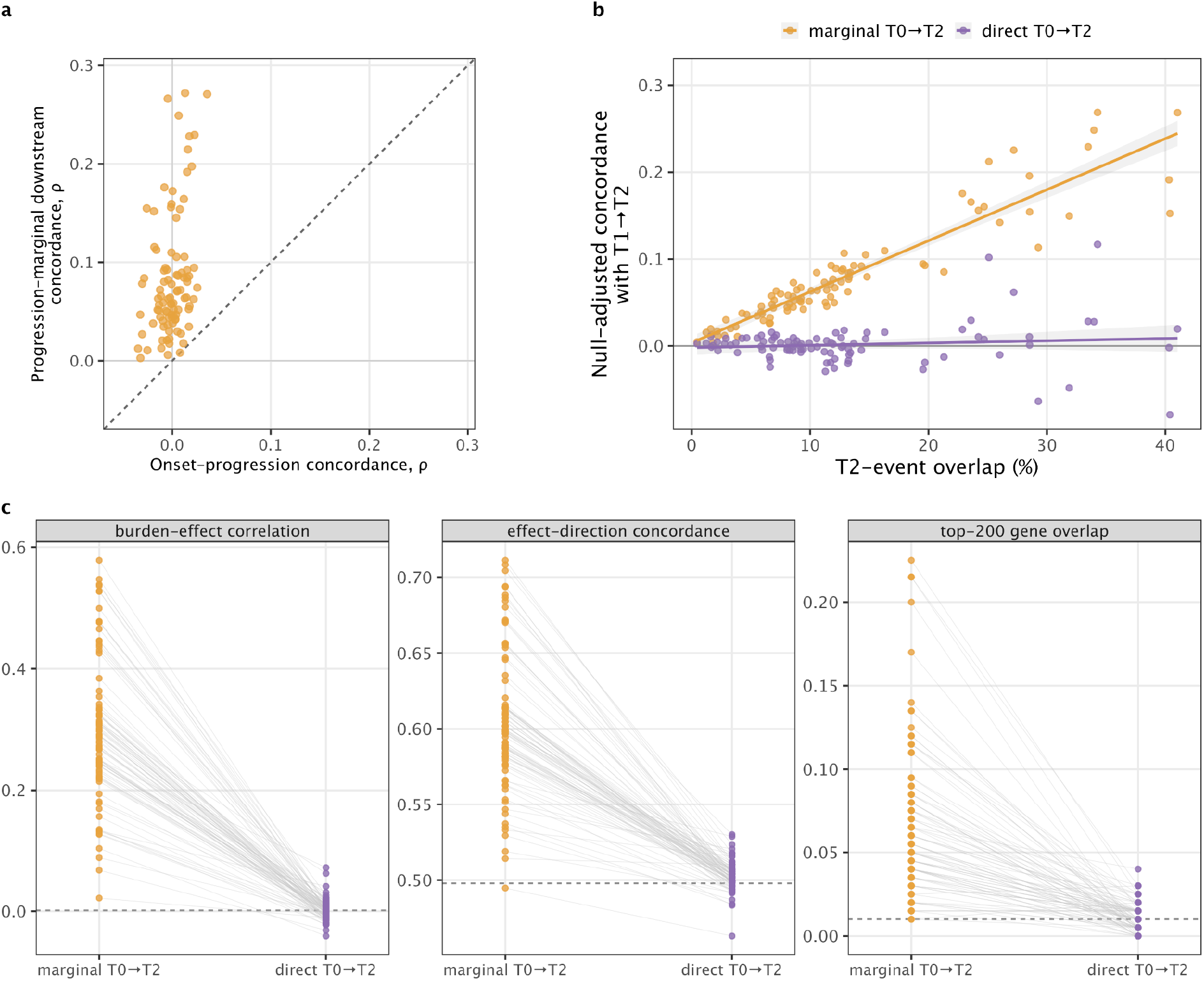
Rare-variant association concordance across disease transitions and T2-event overlap in UK Biobank a, Comparison of gene-level association concordance between progression (T1→T2) and upstream onset (T0→T1) versus concordance between progression and marginal downstream risk (marginal T0→T2) across 95 UKB-EUR trajectories. Each point represents one trajectory; the dashed line denotes the identity line. **b,** Relationship between T2-event overlap and null-adjusted progression-downstream concordance for marginal and direct T0→T2 transitions. T2-event overlap was defined as the proportion of marginal downstream T2 events occurring after incident T1. Null-adjusted concordance was calculated by subtracting the median gene-level Spearman correlation among matched disjoint transition pairs from the observed correlation. Each point represents one trajectory; lines show fitted linear regressions with 95% confidence bands. For both marginal and direct comparisons, the x-axis represents the T2-event overlap fraction in the original marginal downstream analysis **c,** Within-trajectory comparison of progression-downstream concordance between marginal and direct T0→T2 definitions using burden-effect correlation, effect-direction concordance and overlap among the 200 highest-ranked genes. Lines connect the same trajectory across downstream definitions. Dashed horizontal lines indicate the corresponding null expectations.

This comparison suggested another explanation. Both concordance measures included the same T1→T2 progression analyses, but only marginal T0→T2 included T2 events that occurred after T1 that also contributed to progression. We therefore investigated whether statistical dependence induced by overlapping T2 events contributed to the greater progression-downstream concordance, independently of any underlying genetic sharing. For each trajectory, we quantified T2-event overlap as the proportion of marginal T0→T2 events occurring after incident T1. We calculated null-adjusted concordance by subtracting the median gene-level Spearman correlation among matched pairs of transitions sharing no disease state from the observed correlation. For every 10-percentage-point increase in T2-event overlap, null-adjusted progression-downstream concordance increased by 0.059 (95% CI: 0.054-0.064). T2-event overlap explained 86% of the variation in concordance across trajectories (Spearman *ρ* = 0.95; **Fig. 3b**). The association remained robust after accounting for trajectories sharing the same T1 or T2 phenotype (two-way cluster-adjusted estimate, 0.059; 95% CI, 0.043-0.075; **Supplementary Table 22**).

We next tested this explanation using simulated null phenotypes with no genetic effects, while preserving each trajectory’s event counts, risk-set structure and proportion of T2 events occurring after T1. Even under the genetic null, shared events induced correlated sampling variation in association statistics. The simulated increase in null-adjusted progression-downstream concordance was 0.056 (95% CI, 0.052-0.059) for every 10-percentage-point increase in T2-event overlap, closely matching the observed estimate of 0.059. When we assumed that only 60% of incident T1 events were recorded, the relationship remained positive but was attenuated (0.030; 95% CI, 0.026-0.034). These findings show that overlapping T2 events can reproduce the observed concordance gradient without genetic effects, although they do not exclude underlying genetic sharing between transitions. We next examined whether progression-downstream concordance persisted after removing overlapping T2 events from the downstream analysis.

### Removing T2-event overlap reduces concordance to near-null levels

To exclude overlapping T2 events from downstream phenotypes, we replaced marginal T0→T2 with a cause-specific direct T0→T2 transition, in which follow-up ended at incident T1 (**Fig. 1b**). Of the 95 primary trajectories, 91 retained at least 50 direct T0→T2 events and were included in the paired comparison. Across these trajectories, median null-adjusted concordance with progression (T1→T2) fell from 0.064 (IQR: 0.043-0.092) for marginal T0→T2 to 0.001 (IQR: −0.006 to 0.009) for direct T0→T2. Concordance decreased in all 91 trajectories (paired two-sided Wilcoxon signed-rank test, *P* = 1.2 × 10^-16^; **Fig**. **3b****; Supplementary Table 23**).

Two complementary analyses further supported the role of T2-event overlap. First, among 66 trajectories spanning 19 downstream phenotypes represented by multiple T1 states, we held the marginal T0→T2 phenotype fixed while varying T1. For every 10-percentage-point increase in T2-event overlap, null-adjusted concordance increased by 0.076 for marginal T0→T2 but only 0.017 for direct T0→T2. Thus, even for the same downstream disease, greater T2-event overlap was associated with stronger concordance, whereas this relationship was substantially attenuated after excluding overlapping events. Second, in an exploratory analysis stratified by whether T1 and T2 belonged to the same disease-system category, the corresponding increase for direct T0→T2 was weakly positive within disease systems (0.012) but negative across disease systems (−0.014), despite similar distributions of T2-event overlap. This small residual association within disease systems could reflect shared biology, diagnostic entanglement or both.

The attenuation extended beyond correlations of gene-level *P*-value ranks (**Fig. 3c**). Replacing marginal T0→T2 with direct T0→T2 reduced median burden-effect correlation with progression from 0.287 to 0.002, effect-direction concordance from 59.9% to 50.2%, and overlap among top-200 genes from 5.5% to 1.0%. A stricter sensitivity analysis excluding all participants who ever had recorded T1 diagnoses yielded similarly near-zero null-adjusted progression-downstream concordance (0.0012), consistent with the direct-transition result. In additional illness-death simulations^28,29^ incorporating a cause-specific direct T0→T2 transition and varying the degree of causal-gene sharing, progression-direct downstream concordance remained near zero even when all simulated direct-transition causal genes were shared with progression. Although the simulations reproduced the observed association and burden-effect correlations approximately, they did not reproduce effect-direction concordance. These findings suggest that limited progression power can obscure substantial underlying genetic sharing (**Supplementary Note 7; Supplementary Table 24**).

Together, these findings indicate that the greater concordance between progression and marginal downstream analyses largely arises from overlapping T2 events. However, near-zero concordance after removing these events does not establish an absence of underlying genetic sharing, particularly given the limited power of rare-variant progression analyses. Moreover, direct T0→T2 represents a cause-specific transition rather than a causal decomposition of downstream genetic effects, because censoring at T1 changes the population remaining at risk for T2. Thus, observed rare-variant association concordance is strongly influenced by the clinical transitions represented in phenotype definitions and should not, by itself, be interpreted as a measure of underlying genetic sharing.

### Cross-biobank analyses reproduce route-dependent association patterns and expand progression discovery

We applied the same disease-trajectory framework in AoU to test whether the UKB findings were reproduced across ancestry groups. We then combined results across UKB and AoU to increase power for progression discovery (**Fig1d**; **Methods; Supplementary Note 8**).

### Route-dependent association patterns are reproduced across ancestry groups

As in UKB, AoU showed little detectable sharing of gene-level association signals between disease onset and subsequent progression. After adjustment for concordance among matched pairs of transition analyses sharing no disease state, median onset-progression concordance remained near zero in the EUR, AFR and AMR groups (−0.0008, −0.0021 and −0.0005, respectively; **Fig. 4a**; **Supplementary Fig. 17**).

**Fig. 4.**
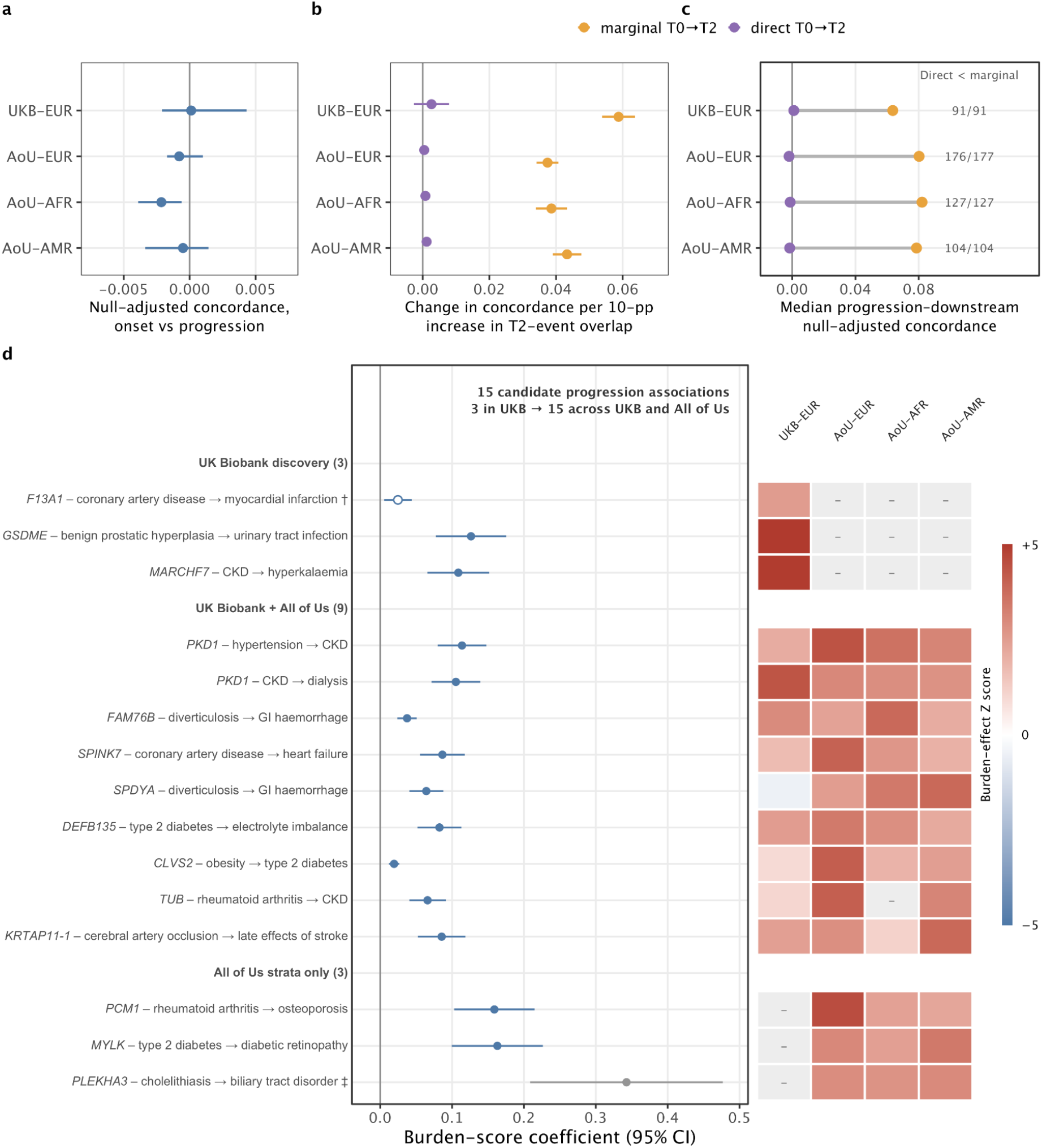
T2-event-overlap patterns reproduce across biobanks and meta-analysis expands progression discovery. a, Onset-progression association concordance in UK Biobank European-ancestry participants (UKB-EUR) and All of Us (AoU) European (EUR), African (AFR) and Admixed-American (AMR) ancestry groups. Points show null-adjusted gene-level concordance between upstream onset (T0→T1) and progression (T1→T2); horizontal bars indicate 95% confidence intervals. **b,** Association between T2-event overlap and null-adjusted progression-downstream concordance. Points show the estimated change in concordance per 10-percentage-point increase in the proportion of marginal downstream T2 events occurring after incident T1; horizontal bars indicate 95% confidence intervals. Orange denotes marginal T0→T2 analyses and purple denotes direct T0→T2 analyses. **c,** Attenuation of progression-downstream concordance after replacing marginal with direct T0→T2. Points show median concordance across trajectories; grey lines connect marginal and direct estimates within each cohort or ancestry group. Annotations indicate the number of trajectories with decreased concordance relative to the number included in the paired comparison. The vertical dashed line denotes zero null-adjusted concordance. **d,** Cross-biobank meta-analysis identified 12 additional exome-wide significant progression associations beyond the three identified in UKB, yielding 15 distinct statistical candidates across the UKB and combined discovery analyses; all 15 are shown. Forest plots show meta-analytic burden-score effects on the log-hazard-ratio scale with 95% confidence intervals. Associations are grouped by which strata contributed to the plotted meta-analysis: UK Biobank discovery, UK Biobank + All of Us, and All of Us strata only. †F13A1 was identified through a variance-component signal; the burden-score coefficient is shown for descriptive comparison and does not establish effect direction (open symbol). ‡PLEKHA3 is shown as exploratory (grey symbol) because the selected mask was supported by a small cumulative minor allele count. Heatmap cells show stratum-specific burden-effect Z scores for the selected variant mask; dashes indicate that no eligible estimate was available for that stratum at that mask, not that the gene was untested in that cohort.

By contrast, progression was more concordant with marginal T0→T2 than with onset, consistent with UKB. Among 177 EUR, 127 AFR and 104 AMR trajectories with results for both marginal and direct T0→T2, median null-adjusted progression-downstream concordance was 0.080, 0.082 and 0.079, respectively. Replacing marginal T0→T2 with direct T0→T2 reduced these values to −0.0019, −0.0012 and −0.0015, respectively. Concordance decreased in nearly all trajectories across the three ancestry groups (paired Wilcoxon signed-rank *P* < 1.0 × 10^-18^; **Fig. 4b ; Supplementary Fig. 18**).

The relationship between T2-event overlap and concordance was also reproduced across ancestry groups (**Fig. 4c)**. For every 10-percentage-point increase in T2-event overlap, null-adjusted progression-downstream concordance increased by 0.037-0.043 for marginal T0→T2, compared with 0.059 in UKB. T2-event overlap accounted for 68-80% of the between-trajectory variation in concordance (Spearman *ρ* = 0.95, 0.91 and 0.93 in EUR, AFR and AMR, respectively). After marginal T0→T2 was replaced with direct T0→T2, the corresponding increases fell to 0.0005-0.0012, with T2-event overlap accounting for no more than 4% of the variation. The smaller estimates in AoU than in UKB may reflect differences in longitudinal follow-up and disease ascertainment, although these contributions were not directly evaluated.

Together, these results reproduce two central UKB findings in an independent biobank across three ancestry groups: little detectable sharing of gene-level association signals between onset and progression, and the near-complete attenuation of progression-downstream concordance after excluding overlapping T2 events.

### Cross-biobank meta-analysis expands progression discovery

To increase power for progression discovery, we combined results from UKB and three ancestry groups in AoU (EUR, AFR and AMR) using fixed-effect meta-analysis of burden effects within each variant mask, followed by Cauchy combination of the resulting *P* values across up to nine prespecified masks (**Methods**). Cross-biobank meta-analysis identified 12 additional exome-wide significant progression associations beyond the three initially discovered in UKB, yielding 15 candidate associations across the two analyses (**Fig. 4d ; Supplementary Tables 25-27**). One additional association, *PLEKHA3*, was supported by very few minor alleles across contributing strata for the most significant variant mask and was considered exploratory. Among the remaining 11 associations, random-effects meta-analysis of the most significant mask yielded nominal significance for all 11 (*P* < 0.05), with seven retaining exome-wide significance (*P* < 2.5 × 10⁻⁶). Leave-one-stratum-out analyses showed generally similar mask-level burden-effect estimates, although only the two *PKD1* associations remained exome-wide significant after excluding any single contributing stratum (**Extended Data Fig. 9**).

The additional associations spanned several clinical trajectories including *MYLK* with type 2 diabetes→diabetic retinopathy, *PCM1* with rheumatoid arthritis→osteoporosis and *SPINK7* with CAD→heart failure. Some associations were supported by sparse rare-variant information or showed heterogeneous effects across cohorts. Notably, *MYLK* and *PCM1* had no eligible UKB results for their meta-analysis-selected masks, while alternative masks testable in UKB showed no significant association, indicating that their discovery signals were driven by AoU.

The strongest additional signals involved *PKD1* with hypertension→CKD (*P* = 2.1 × 10^-10^; burden-score HR = 1.12; 95% CI, 1.08-1.16) and CKD→dialysis (*P* = 4.9 × 10^-9^; HR = 1.11; 95% CI, 1.07-1.15). To assess the contribution of underlying polycystic kidney disease, we excluded participants with recorded cystic kidney disease. This removed only 1.4-5.6% of the corresponding risk sets while eliminating 56-100% of qualifying *PKD1* LoF alleles. The burden-meta association signals were consequently attenuated to *P* = 0.74 for hypertension→CKD and *P* = 0.30 for CKD→dialysis. Restricting the exclusion to cystic kidney disease diagnoses recorded within 90 days of progression risk-set entry produced less attenuation, consistent with delayed recognition of underlying cystic disease (**Supplementary Table 28**). These findings suggest that the *PKD1* associations may largely reflect underlying cystic kidney disease rather than progression-specific genetic effects, highlighting the importance of disease history and ascertainment when interpreting genetic associations with clinical progression.

Although aggregate trajectory-level association patterns were consistent across biobanks, support for individual progression associations was more variable (**Extended Data Fig. 10 Supplementary Figs. 19 and 20 ; Supplementary Table 29**). Among the three progression associations initially identified in UKB, *F13A1* with CAD→MI and *MARCHF7/MARCH7* with CKD→hyperkalaemia showed nominal support in AoU-EUR (one-sided *P* = 0.027) and AoU-AFR (one-sided *P* = 0.028), respectively. By contrast, *GSDME/DFNA5* with benign prostatic hyperplasia→urinary tract infection showed no corresponding association evidence in AoU despite relatively high estimated replication power. None of these three associations reached exome-wide significance in the combined meta-analysis.

## Discussion

We developed GATE-GENE to enable scalable gene-based rare-variant association analyses with disease timing and progression in longitudinal biobanks. GATE-GENE maintained type I error control under the sparse carrier-event counts and extreme censoring encountered in progression studies, including settings in which the evaluated GATE-STAAR variance-component and omnibus tests became inflated, and gained modest power over matched case-control analysis when event timing was informative. Applying GATE-GENE across UKB and AoU yielded a broad atlas of rare-variant associations with disease onset and subsequent progression, while demonstrating that observed progression-downstream association concordance was strongly influenced by overlapping outcome events. Together, these findings establish the feasibility of rare-variant survival analysis at biobank scale and highlight the importance of event structure and phenotype definition when comparing genetic associations across disease stages.

Previous studies have shown that genetic associations with disease susceptibility can differ from those with subsequent clinical outcomes or mortality^3–5^. Our results extend this question to rare-coding variants across a broad range of disease trajectories. Gene-level association concordance between onset and progression remained low after harmonizing tested genes and variant masks and accounting for differences in event information and test availability. Sensitivity analyses did not reveal systematically negative effect correlations suggestive of pervasive index-event bias, although they could not exclude its contribution. Importantly, progression analyses contained substantially fewer events and carrier-events than onset analyses, limiting the ability to detect shared associations. Simulations showed that the interpretation of low concordance depended on assumptions about underlying genetic architectures and effect sizes. Thus, the observed association profiles showed limited detectable concordance, but this does not establish that the underlying rare-variant genetic architectures are largely distinct.

The progression-downstream comparison illustrates an additional challenge in interpreting longitudinal genetic associations. A marginal association with a downstream disease combines individuals who reach that endpoint through different clinical sequences, whereas a progression analysis is restricted to individuals who have already entered an earlier disease state. Marginal T0→T2 and progression T1→T2 analyses can therefore share the same T2 events, inducing correlated association statistics even in the absence of shared genetic effects. Consistent with this mechanism, progression-downstream concordance increased almost linearly with T2-event overlap, the observed gradient was reproduced under genetically null simulations, and concordance fell to near-null levels when overlapping events were removed. These patterns were reproduced in UKB and across three AoU ancestry groups. Additional illness-death simulations showed that near-zero progression-direct downstream concordance could persist even under extensive causal-gene sharing in the simulated architectures. Thus, high concordance between progression and marginal downstream analyses can arise from shared events, whereas low concordance after their removal does not necessarily imply distinct underlying genetic effects.

The phenome-wide onset atlas provides a complementary resource for studying rare-variant associations with disease timing. GATE-GENE recovered established disease-gene relationships and showed broad agreement with matched incident case-control analyses, supporting the validity of the survival-analysis framework. By incorporating diagnosis timing and systematically curated longitudinal phenotypes, the atlas extends existing large-scale case-control rare-variant resources such as GeneBass^11^ and BRaVa^30^ beyond binary disease status alone. The onset atlas and multi-state trajectory analyses demonstrate the utility of GATE-GENE for investigating rare-variant associations with disease onset, subsequent progression and successive stages of a clinical course.

Cross-biobank meta-analysis expands the discovery of rare-variant progression associations, although individual signals showed variable support across cohorts and ancestry groups. Importantly, statistical associations with progression do not necessarily represent genetic effects specific to the transition under study. The *PKD1* findings illustrate how pre-existing disease susceptibility and incomplete diagnostic histories can complicate the interpretation of progression associations, while the accompanying loss of genetic information in sensitivity analyses limits definitive attribution. More broadly, sparse carrier information and heterogeneous cohort-specific effects emphasize the need for careful clinical characterization and independent validation before assigning progression-specific biological interpretations.

Several limitations remain. Disease timing was inferred from EHR and registry diagnoses, so first recorded diagnosis is an imperfect proxy for biological onset, and differences in healthcare utilization, coding practices and left truncation may affect transition timing and classification of clinical routes. Progression risk sets were substantially smaller than T0-based cohorts, with sparse carrier-events limiting not only association discovery but also the ability to distinguish limited from extensive underlying genetic sharing across disease transitions. Larger sequencing cohorts with more informative progression carrier-events and longer longitudinal follow-up will be needed to better resolve shared and transition-specific rare-variant genetic effects. Cross-biobank meta-analysis relied on gene-and mask-level summary statistics because variant-level score statistics could not be exchanged across research environments, limiting assessment of variant-level consistency and sources of heterogeneity. Primary discovery was anchored in UKB participants of European ancestry, and broader validation across ancestries and healthcare systems remains necessary. Death was treated as censoring rather than modelled as a competing transition, so the estimated cause-specific hazards do not directly represent cumulative incidence in the presence of competing mortality. Finally, our analyses focused on rare-coding variants, leaving the contribution of other classes of genetic variation across disease transitions to future investigation.

More broadly, our findings show that the clinical route encoded by a longitudinal phenotype is an important component of genetic study design. Marginal downstream associations may capture genetic influences operating through different clinical pathways, while progression analyses focus on transitions conditional on an earlier disease state. Distinguishing these routes and accounting for overlapping events are essential for interpreting observed association concordance without conflating it with underlying genetic sharing. By integrating disease history and transition structure into large-scale genetic studies, GATE-GENE provides a framework and resource for investigating how rare variants relate to disease susceptibility, timing and progression.

## Methods

### Study cohort and participant inclusion

The UK Biobank (UKB) is a population-based cohort of approximately 500,000 participants recruited at 40-69 years of age^31^. All participants provided written informed consent, and ethical approval was obtained from the National Research Ethics Service Committee. Whole-exome sequencing (WES) was performed by the UK Biobank Exome Sequencing Consortium for 454,787 participants^12^, including 390,971 White British participants. The phenome-wide onset atlas used this WES release, with incident diagnoses ascertained exclusively from Hospital Episode Statistics (HES) inpatient records and a median analytical sample size of 238,296 participants per phenotype. The multi-state analyses instead used the interim approximately 200,000-participant UKB WES release^32^ and included otherwise eligible controls regardless of whether they had a recorded HES encounter; the median progression risk-set size was 4,234. Simulations used 100,000 randomly selected White British participants.

All of Us Research Program (AoU) analyses used version 8 data, including short-read whole-genome sequencing (WGS) and longitudinal electronic health records^33,34^. Genetic ancestry assignments provided by AoU were based on a random-forest classifier trained on reference populations; participants with posterior probability >0.75 for a given ancestry were retained, and samples flagged for quality-control issues were excluded. Analyses were conducted separately in participants of European (EUR; n = 223,118), African (AFR; n = 79,500) and Admixed-American (AMR; n = 71,610) genetic ancestry.

### Phenotype definition and curation for the incident-onset atlas

For the UKB onset atlas, incident disease ascertainment used ICD diagnoses from linked HES inpatient records, mapped to PheCodes^35^. Incident phenotypes were defined as the first qualifying diagnosis recorded at least 30 days after cohort entry, with time measured from study entry to event or censoring. These phenotypes therefore represent time to first recorded diagnosis during follow-up rather than biological disease onset in all individuals. Cohort-specific procedures for detecting prevalent conditions, defining eligible prior observation, handling the 30-day landmark and establishing participant-specific follow-up end dates are given in **Supplementary Notes 3, 6 and 8**.

Phenotypes were curated in two stages. First, we applied a content-based filter to retain clinically meaningful disease-state phenotypes and exclude symptoms and signs, injuries and poisonings, procedure-, treatment-, history-or status-related codes, vague residual categories, and exact or near-exact duplicates. When parent and child phecodes overlapped, we retained a single representative phenotype in the primary analysis set, prioritizing the more clinically canonical and interpretable endpoint while also considering event count and redundancy. Second, we applied an onset-validity screen to distinguish phenotypes appropriate for the primary broad incident-onset analysis from those whose recorded event time was more likely to reflect delayed ascertainment, downstream complications, acute decompensation, congenital timing, or other context-dependent clinical events. After curation, 314 phenotypes were retained in the primary broad incident-onset analysis set, 337 were retained for supplementary or sensitivity analyses, and 183 were excluded. Of the 314 primary phenotypes, 14 were designated timing-limited because first recorded diagnosis in a middle-aged to older cohort may not adequately represent biological onset. The remaining 300 phenotypes constituted the core onset atlas used for primary analyses. All onset analyses used time since study entry as the analysis time scale and were adjusted for age at study entry, sex and the first ten genetic principal components; sex was omitted for sex-restricted phenotypes.

#### Sequencing data and rare-variant masks

The UKB onset atlas used the full 450k whole-exome sequencing OQFE callset on GRCh38, whereas the multi-state trajectory analyses used the interim approximately 200k UKB exome release. Variants were assigned to genes using the gene-level group files previously constructed for SAIGE-GENE+ analyses of the UKB exomes^12,24^, which classify variants into loss-of-function (LoF), missense and synonymous functional categories. We did not perform additional sample-or variant-level sequencing quality control or reannotation beyond that incorporated into the released UKB callset and previously generated group files. The two UKB datasets were analysed separately; no participant-level pooling was performed between the onset atlas and the trajectory analyses. Exact source release identifiers, ancestry filters and variant-group file provenance should be reported for each component after verification.

AoU analyses used exonic regions of the version 8 short-read whole-genome sequencing callset and corresponding gene-level group files^34^. Functional classes comprised predicted LoF, missense including low-confidence LoF and synonymous variants. Gene and annotation definitions were harmonized between UKB and AoU for cross-cohort analyses.

Rare coding variants were evaluated using nine prespecified masks comprising three maximum minor-allele-frequency thresholds (1%, 0.1% and 0.01%) crossed with three nested functional classes: LoF, LoF plus missense and LoF plus missense plus synonymous variants. Rare variants were weighted using Beta(MAF; 1,25) weights, and ultra-rare variants with minor allele count (MAC) ≤10 were collapsed before set-based testing following the SAIGE-GENE+ strategy^24^.

#### GATE-GENE rare-variant time-to-event association testing

GATE-GENE is a scalable gene-based rare-variant association framework for time-to-event (TTE) phenotypes. For each phenotype, GATE-GENE first fits a mixed-effects frailty model including fixed covariates and a Gaussian frailty term parameterized by a genetic relationship matrix, thereby accounting for censoring, relatedness and population structure^19,36^. Variant score statistics and their covariance are then used for gene-based burden, SKAT^22^ and SKAT-O^23^ testing.

To improve calibration when carrier-event counts are sparse, GATE-GENE applies saddlepoint approximation (SPA) to single-variant score statistics before gene-based aggregation^37–39^. A variance-ratio approximation is used to reduce the computational cost of matrix-adjusted score-variance estimation. Detailed derivation of the frailty model, score statistics, variance approximation, ultra-rare-variant collapsing and SPA adjustment is provided in **Supplementary Note 1**.

Within each annotation-by-frequency mask, burden, SKAT and SKAT-O statistics were calculated. Unless otherwise specified, primary gene-level evidence was based on SKAT-O, with mask-specific *P* values combined using the Cauchy combination method^40,41^ to obtain a per-gene omnibus *P* value. Burden estimates were used to aid interpretation of effect direction. GATE-GENE supports user-defined functional annotations and maximum minor-allele-frequency thresholds; in this study, we evaluated nine prespecified masks defined by three maximum MAF thresholds crossed with three nested functional classes. Exponentiated burden coefficients are interpreted as hazard ratios per unit of the Beta(1,25)-weighted genetic burden score, not as per-allele or per-carrier hazard ratios (**Supplementary Note 6**).

Exome-wide significance was defined as *P* < 2.5 × 10⁻⁶ within each phenotype. Because this threshold controls testing within rather than across phenotypes, we additionally applied Benjamini-Hochberg false-discovery-rate control^42^ across all gene-phenotype tests in the 300-phenotype onset atlas as a study-wide multiplicity sensitivity analysis. UKB Progression, native AoU and cross-biobank meta-analyses were treated as separate testing families (**Supplementary Tables 27 and 29**). FDR correction was applied to the Cauchy-combined gene-level omnibus *P* values for the onset atlas, harmonized association *P* values for progression and region-level omnibus *P* values for native AoU. Thus, the onset-atlas FDR analysis applied an additional multiple-testing correction to the same statistic used for discovery, rather than providing independent evidence. For cross-biobank meta-analysis, FDR correction instead used Stouffer-combined omnibus *P* values, whereas discovery used Cauchy-combined *P* values; consequently, the two analyses were not based on identical test statistics.

#### Calibration, power simulations and method comparison

Calibration and power were evaluated using exome genotypes from 100,000 randomly selected White British UKB participants, preserving realistic population structure and relatedness. Null TTE phenotypes were simulated under censoring rates of 90%, 95% and 99%, and GATE-GENE was evaluated with and without saddlepoint approximation (SPA). Power was evaluated at 90% censoring under three rare-variant causal architectures differing in causal-variant proportions and effect sizes and was compared with SAIGE-GENE+ applied to corresponding incident binary phenotypes using the same participants, genotypes and variant masks. Full generative models, causal architectures and simulation parameters are provided in **Supplementary Note 2**.

To benchmark calibration against an existing gene-based rare-variant TTE method, GATE-STAAR was applied to the same simulated null phenotypes. Because its pipeline required method-specific genotype conversion and substantially larger intermediate files, the controlled comparison was restricted to chromosome 19. We evaluated LoF, missense and synonymous variant groups at maximum MAF thresholds of 1%, 0.1% and 0.01% across 200 null phenotype replicates per censoring level. GATE-STAAR was run using its implemented default ultra-rare-variant collapsing threshold (MAC ≤ 20) and default burden, SKAT and STAAR-O settings. Participants, phenotype replicates, censoring mechanisms, covariates, eligible variants and tested genes were matched between methods wherever the corresponding masks permitted. Empirical type I error was estimated separately by test, MAF threshold and censoring level, with exact binomial 95% confidence intervals. Exact software releases and commits, container identifiers, command-line settings, SPA configuration, weighting schemes, input-conversion procedures, numbers of variants, genes and tests, and the distinction between the implemented ultra-rare threshold and that described in the original GATE-STAAR report are provided in **Supplementary Note 2**.

### Curation of germline-focused associations

Because UKB exome sequencing was performed using blood-derived DNA, exome-wide significant associations were screened for signals attributable to clonal haematopoiesis, haematologic neoplasia and other somatic processes. A curated set of 42 genes recurrently implicated in clonal haematopoiesis or myeloid or lymphoid neoplasia was used together with phenotype context and variant-level association patterns to classify associations as germline-likely, somatic-likely, somatic-suspect, dual germline-somatic or haematologic-fingerprint signals^32,43–50^. Associations classified as somatic-likely or somatic-suspect were excluded from the primary germline-focused atlas. Associations involving genes with both germline predisposition and recurrent somatic mutation were evaluated at the variant level, and extended-MHC signals were additionally examined for long-range linkage-disequilibrium confounding. The complete gene list and per-association classifications are provided in **Supplementary Tables 7 and 30**.

### Evaluation of phenome-wide incident-onset associations

To assess the contribution of event-time modelling beyond the incident case definition, we compared GATE-GENE with matched SAIGE-GENE+ incident case-control analyses for 120 onset phenotypes selected to span censoring levels, onset-age distributions and PheCode categories. Case-control analyses used the same samples, phenotype definitions and variant masks. Concordance was summarized using the Spearman correlation of gene-level −log10*P* values, overlap among the top 50 genes and sharing of exome-wide significant associations.

For the 107 germline-focused exome-wide associations, carrier-level disease timing was evaluated using Cox proportional-hazards models with attained age as the time scale and delayed entry at cohort enrolment. Rare LoF carriers were compared with non-carriers, with adjustment for sex and the first 20 genetic principal components; a sensitivity analysis additionally adjusted for birth year. Associations with sufficient carrier-event counts for stable estimation were tested, with Benjamini-Hochberg correction across analysable associations.

GATE-GENE onset results were also compared with phenotype-matched GeneBass15 prevalent case-control exome results. Ninety-nine core phenotypes had results available in both resources. Genome-wide concordance was evaluated over genes tested in both resources, and GeneBass evidence among the 200 genes most strongly prioritized by GATE-GENE was compared with that among the remaining genes within each phenotype. Because both resources use UKB exome data, this analysis was interpreted as cross-analysis support rather than independent replication. Detailed phenotype mapping and analyses are provided in **Supplementary Note 3**.

### Functional enrichment and rare-common convergence

Functional enrichment among germline-focused onset-associated genes was evaluated using Hallmark, Reactome and Gene Ontology Biological Process gene sets^51–53^, together with GTEx v8 tissue-expression gene sets^54^ (**Supplementary Table 31**). Over-representation was tested against the universe of genes with valid association results using one-sided Fisher’s exact tests with false-discovery-rate correction. Robustness was evaluated using gene sets matched on characteristics influencing rare-variant testability and through leave-one-gene-out analyses.

Rare-common convergence was evaluated at phenotype, locus and pathway levels. Common-variant survival association analyses were performed for the 300 core onset phenotypes using GATE, and independent common-variant loci were defined by LD clumping^55^. At the phenotype level, we tested whether the number of genome-wide significant common-variant loci correlated with the number of germline-focused rare-variant gene discoveries, with permutation and count-model sensitivity analyses accounting for differences in discovery opportunity. At the locus level, we tested whether rare-significant genes occurred more frequently than expected near phenotype-matched common-variant loci using a covariate-matched permutation null; the extended MHC was excluded from the primary analysis. Rare-variant associations were also refitted conditioning on phenotype-specific common lead variants. At the pathway level, common-variant gene-set enrichment was performed using MAGMA^56^ and compared with rare-variant pathway enrichment. Full definitions, permutation procedures, conditional-fit diagnostics and sensitivity analyses are provided in **Supplementary Notes 4 and 5**.

### Definition of multi-state disease trajectories

Disease trajectories were represented by baseline (T0), an upstream disease state (T1) and a downstream disease state (T2). For each trajectory, we defined three primary transition-specific association analyses: upstream disease onset (A; T0→T1), post-diagnosis progression (B; T1→T2) and marginal downstream risk (C; T0→T2) that followed participants to T2 irrespective of whether T1 occurred first. A fourth, cause-specific direct downstream transition (D; T0→T2, censoring at incident T1) was constructed to evaluate the contribution of overlapping T2 events to observed concordance.

Candidate trajectories were assembled before examination of genetic association results using clinically established disease sequences, the disease-trajectory literature and curated candidate lists^16–18^. We prioritized clinically interpretable ordered disease pairs with epidemiological support and sufficient endpoint counts for transition-specific analysis. Of 199 UKB candidate trajectories, 95 had all three primary transitions available and at least 100 T1→T2 progression events and were retained for the primary analysis.

All transition analyses used time since transition-specific risk-set entry as the analysis time scale. For T0→T1 and marginal T0→T2, the time origin was UKB recruitment; participants with prevalent endpoint disease were excluded and a 30-day landmark was applied. For T1→T2, the risk set comprised participants with incident T1 who remained free of T2 at the T1 date, and the time origin was reset to the incident T1 date. Participants with T2 before or concurrent with T1 were excluded. The time scales and risk-set definitions distinguish direct T0→T2 censoring at incident T1 from the stricter T1-free sensitivity analysis, which excludes participants with T1 recorded at any time (**Supplementary Note 6**).

All transition analyses were adjusted for age at risk-set entry, sex and the first ten genetic principal components, with sex omitted for sex-restricted phenotypes. Progression analyses were additionally adjusted for birth year. Death was treated as censoring; transition estimates therefore represent associations with the cause-specific transition hazard among participants remaining alive and under observation rather than effects on cumulative incidence in the presence of competing mortality.

### T2-event overlap and direct downstream transitions

For each trajectory, T2-event overlap was defined as the proportion of marginal T0→T2 events in which incident T1 occurred after baseline and before T2. This quantity measures the fraction of downstream events that traversed the same T1→T2 clinical route represented by the progression analysis. This is an observed shared-event fraction, not a causal estimate of how disease progressed. The broader distribution of T2 diagnoses by prior and subsequent T1 history is referred to as clinical-route composition.

To exclude T2 events shared with the progression analysis from the downstream comparison, we constructed a cause-specific direct T0→T2 transition in which participants exited the direct-transition risk set at incident T1. Direct-transition analyses were retained when at least 50 downstream events remained. As a stricter T1-free sensitivity analysis, we also repeated the downstream analysis after excluding participants who recorded T1 at any time. Additional tie handling, post-T1 washout analyses and route decomposition are described in **Supplementary Note 6**. Same-day first records of T1 and T2 were treated as having indeterminate ordering and were not counted as a direct T0→T2 event. This route-specific exclusion is not equivalent to excluding all participants with an observed T1 diagnosis (**Supplementary Note 6**).

### Phenotypic relationship between paired disease states

To provide clinical context for the gene-level concordance analyses, we quantified the relationship between incident T1 and subsequent T2 for each of the 95 trajectories using the same phenotype definitions, baseline-entry criteria and censoring dates as the corresponding transition analyses. Participants were required to be free of both T1 and T2 at recruitment, such that these analyses compared incident disease states.

For each trajectory, we modeled time to T2 using a Cox proportional-hazards model with incident T1 as a time-varying exposure. Participants contributed unexposed person-time until T1 onset and exposed person-time thereafter. Models used attained age as the analysis time scale, with left truncation at age at recruitment, and were adjusted for sex. T1 was considered to precede T2 only when its recorded date was earlier than the T2 date. Trajectories with ≤5 T2 events among participants without incident T1 were summarized separately because the corresponding hazard ratios could not be estimated stably; 79 of 95 trajectories contributed to the summary of T1–T2 hazard ratios. We additionally calculated, for each trajectory, the proportion of participants with incident T1 who subsequently developed T2.

To test whether the strength of the clinical T1–T2 relationship explained onset–progression concordance, we related the transition proportion to the Spearman correlation of gene-level −log₁₀(*P*) values between T0→T1 and T1→T2 across trajectories. We used the transition proportion for this analysis because it was not correlated with progression event count, whereas hazard-ratio and incidence-rate estimates were. Associations were evaluated using Spearman correlation and in sensitivity analyses using rank regression adjusted for progression event count.

### Gene-level association concordance across transitions

Gene-level association concordance between transition-specific analyses was quantified using Spearman correlation of gene-level –log10*P* values. Because the rarest MAF cells were frequently unavailable in the smaller progression risk sets, the primary concordance analyses used a harmonized omnibus statistic constructed from the same six annotation-by-frequency cells in every transition: the three functional masks at maximum MAF thresholds of 1% and 0.1%. Only genes with valid harmonized omnibus results in the relevant transitions were retained.

For each trajectory, we calculated progression-onset concordance, *ρ*(T1→T2, T0→T1), and progression-downstream concordance, *ρ*(T1→T2, T0→T2), and compared these paired correlations across trajectories using Wilcoxon signed-rank tests. We additionally calculated progression-direct downstream concordance for trajectories with an eligible cause-specific D transition and compared B-C against B-D concordance within the same 91 UKB trajectories. Both contrasts used the same progression transition, whereas their downstream event definitions differed.

To account for differences in statistical power and pair composition, null-adjusted concordance was defined relative to an empirical null constructed from disjoint transition pairs sharing no disease state. Null pairs were matched on minimum event-count stratum and pair composition, defined by the number of progression analyses in the pair. Null-adjusted concordance was calculated as the observed Spearman correlation minus the median correlation of matched disjoint pairs. Burden-effect correlation, effect-direction concordance and overlap among the top-ranked 200 genes were evaluated as complementary metrics that did not rely on *P*-value ranks. Tail calibration of trajectory analyses was summarized using the genomic-control factor at the 1st percentile of the gene-level *P*-value distribution (*λ*_0.01_).

### T2-event overlap and dependence-aware analyses

We regressed null-adjusted progression-downstream gene-level concordance on the fraction of marginal T0→T2 events recorded after incident T1. Regression coefficients were expressed per 10-percentage-point increase in this T2-event overlap fraction; confidence intervals accounting for repeated T1 and T2 phenotypes were obtained by two-way clustering.

Because progression analyses condition on reaching T1, we additionally evaluated index-event bias diagnostically by testing whether onset-progression burden-effect correlations were systematically negative; this analysis was not used as a formal correction for index-event bias. To test the T2-event-overlap interpretation without relying on simulation, we performed two empirical control analyses. First, in a within-endpoint analysis, we refitted the T2-event-overlap regression with fixed effects for the downstream phenotype, thereby holding the marginal T0→T2 endpoint constant among endpoints represented by multiple trajectories while allowing the intermediate T1 state to vary. Second, in a cross-system analysis, trajectories were stratified according to whether the intermediate and downstream PheCodes belonged to the same PheCode block, used as a proxy for clinical relatedness; T2-event-overlap distributions were compared between strata, and the analysis was repeated after restriction to their common overlap range. The sign-correlation diagnostic does not remove or estimate index-event bias and cannot establish that selection into the T1 risk set is unbiased.

Because trajectories reused intermediate and downstream phenotypes, they were not treated as independent observations. We therefore evaluated the T2-event overlap regression using four dependence-aware approaches: cluster-robust variance estimation with one-way and two-way clustering by intermediate and downstream phenotype; cluster bootstrap resampling of downstream phenotypes, intermediate phenotypes or disease families (10,000 replicates); a crossed random-intercept model including intermediate-and downstream-phenotype effects; and block permutation of T2-event overlap fractions between endpoint blocks while preserving shared-endpoint structure under the null (10,000 permutations). Leave-one-intermediate, leave-one-downstream and leave-one-disease-family-out analyses further assessed whether the association was driven by individual reused states or disease groups. The same dependence-aware analyses were applied separately within each All of Us ancestry group. Full empirical-null construction, harmonization procedures and sensitivity analyses are provided in **Supplementary Note 6**.

### Longer clinical chains and simulation analyses

To evaluate whether limited cross-transition sharing extended beyond three-state trajectories, the analysed disease states and progression transitions were assembled into a directed graph and simple clinical paths of two to five edges were enumerated. Gene-level concordance between adjacent progression transitions was evaluated using the same harmonized concordance framework and compared with disjoint transition pairs.

Illness-death simulations were used to evaluate whether the observed cross-transition concordance patterns could arise from limited progression power, shared susceptibility effects, index-event selection or incomplete recording of the intermediate disease state. Simulations considered susceptibility-only, shared-effect and transition-specific architectures and were analysed using the same concordance pipeline as the empirical data. Additional simulations varied the fraction of progression causal genes shared with onset to evaluate partially shared architectures. For each sharing fraction, we calculated the proportion of simulated replicates yielding concordance at or below the observed value; sharing fractions for which this proportion was <0.05 were considered inconsistent with the observed data under the simulated architecture. To complement the simulations empirically, trajectories were ranked by progression event count and analyses were repeated in the top 50%, 25% and 10% of trajectories. Onset and marginal downstream comparisons were evaluated within the same trajectory subsets so that progression power was held constant between comparisons (**Supplementary Table 21**). Complete simulation models, parameter settings and interpretation are provided in **Supplementary Note 7**. We interpreted the resulting architecture contrasts conditional on the modelled causal effects, carrier frequencies, event rates and ascertainment; no sharing fraction was inferred directly from an observed near-zero correlation. We distinguish simulations of onset-progression sharing from those of progression-direct sharing: these contrasts have different event structures and cannot be interpreted as estimates of the same causal-gene overlap parameter.

To quantify concordance generated by shared-event structure alone, we additionally constructed T2-event-overlap-preserving null simulations in which all genetic effects were set to zero. Each replicate was matched to an observed trajectory and reproduced its onset, progression and downstream event counts, risk-set structure and fraction of downstream events reached through incident T1. We simulated three replicates for each of 91 trajectories under two intermediate-state ascertainment settings. Excess progression-downstream concordance was calculated relative to the same disjoint-transition null used in the empirical analyses and regressed on the realized T2-event fraction, yielding the T2-event-overlap gradient expected in the absence of genetic effects. We analysed the susceptibility-only architecture in parallel to assess the additional contribution of index-event selection when genetic effects acted only on T0→T1.

To specifically test whether weak progression-direct concordance could coexist with shared rare-variant causal genes, an additional illness-death simulation varied the fraction of direct-transition causal genes shared with progression (0-100%), introduced a cause-specific direct T0→T2 arm censored at incident T1, and calibrated the effect scale to observed progression–marginal association and burden-effect correlations. At the evaluated event information and effect scale, progression-direct correlations remained near zero across sharing fractions, including complete sharing. This analysis did not reproduce observed effect-direction concordance and was treated as a detectability sensitivity analysis rather than a calibrated model of all concordance metrics (**Supplementary Note 7**).

### All of Us trajectory analyses

AoU TTW phenotypes were constructed from OMOP-mapped electronic health records using PheCode v1.2b1 definitions and the same four-transition framework as UKB. Unlike UKB, which used an approximately 30-day landmark after recruitment, AoU required 365 days of prior EHR observation before baseline-entry follow-up began. This longer observation-history requirement was intended to reduce misclassification of pre-existing conditions as incident diagnoses, given the more limited retrospective clinical history available in AoU. Person-time and diagnoses during the initial 365 days were not included as incident follow-up events. Progression follow-up began at incident T1 among eligible participants free of T2. Trajectories with at least 100 progression events within an ancestry group were included, with direct-transition eligibility assessed separately. Event-ordering and censoring definitions were otherwise harmonized across cohorts.

Gene-based testing used GATE-GENE as implemented in SAIGE v1.5.2 on exonic regions of the AoU version 8 WGS callset. Null models were fitted separately in EUR, AFR and AMR using ancestry-specific sparse genetic relationship matrices and were adjusted for age at risk-set entry, sex and the first ten genetic principal components, with birth year additionally included for progression analyses. The cause-specific direct T0→T2 transition was reconstructed within each ancestry using the same definition as in UKB. The UKB and AoU relatedness-matrix cutoffs and the observation-period definitions differ and are specified by cohort in **Supplementary Notes 6 and 8**.

Cross-cohort analyses used exact PheCode-pair matching. Gene symbols were harmonized using a curated HGNC alias map, and ambiguous or untestable mappings were excluded. Variant-mask definitions were harmonized between cohorts. Cross-ancestry reproduction of observed gene-level association-concordance patterns was evaluated using the same concordance framework and transition-pair-composition-matched empirical null as in UKB, constructed separately within each ancestry. Additional AoU phenotype-fitting rules, direct-transition construction and calibration analyses are described in **Supplementary Note 8.**

### Replication and cross-biobank meta-analysis

UKB trajectory-level exome-wide associations were deduplicated to distinct gene-transition associations before evaluation in AoU. For each association, the UKB-selected mask was defined as the annotation-by-frequency cell with the strongest discovery burden signal. Primary replication required concordant burden-effect direction and a one-sided *P* < 0.05 for the same mask in AoU. Benjamini-Hochberg correction was additionally applied across carried-forward associations within each ancestry. Cross-ancestry AoU replication was evaluated using fixed-effect inverse-variance meta-analysis, with heterogeneity summarized using Cochran’s *Q* and *I*^2^.

For combined cross-biobank discovery, burden log-hazard-ratio estimates were meta-analysed within each mask across UKB and AoU EUR, AFR and AMR using fixed-effect inverse-variance meta-analysis. Mask-level meta-analysis *P* values were then combined using the Cauchy method to obtain a gene-level cross-biobank burden-meta omnibus P value. SKAT and SKAT-O statistics were not meta-analysed. Each mask contributed whichever strata had a finite positive standard error, so the contributing stratum set differs between masks of the same gene; a gene-level omnibus *P* value therefore does not imply four-stratum support for an association, and some masks contributing to it have no UKB estimate. Random-effects and leave-one-stratum-out analyses are selected-mask burden analyses rather than omnibus statistics. Cross-biobank meta-analysis was treated as a separate discovery family, with exome-wide significance defined as *P* < 2.5 × 10⁻⁶ per analysis. Leave-one-stratum-out, random-effects and alternative P-value-combination analyses were used to evaluate robustness of new progression associations. Detailed replication-power, winner’s-curse and meta-analysis sensitivity analyses are provided in **Supplementary Note 8**.

### Clinical sensitivity analyses of progression associations

Targeted clinical sensitivity analyses were performed for progression associations requiring additional interpretation. For *PKD1* renal trajectories, participants with recorded cystic kidney disease were excluded to assess whether the associations reflected previously unrecognized polycystic kidney disease rather than progression of common hypertension or chronic kidney disease. A temporally restricted analysis excluded cystic-kidney-disease diagnoses recorded by 90 days after entry into the progression risk set to evaluate delayed recognition.

Selected UKB downstream endpoints were additionally corroborated using linked procedure, prescription and cause-of-death records. Carrier-level models evaluated robustness to pre-entry healthcare-contact intensity and to conservative censoring of downstream events lacking independent corroboration. Code definitions, corroboration windows and sensitivity meta-analyses are provided in **Supplementary Note 8**, and per-endpoint corroboration rates in **Supplementary Table 32**.

## Data availability

Individual-level genetic and longitudinal phenotypic data from the UK Biobank are available to approved researchers through the UK Biobank (https://www.ukbiobank.ac.uk); this study was conducted under application 31063. Individual-level data from the All of Us Research Program are available to authorized researchers through the All of Us Researcher Workbench (https://www.researchallofus.org). This study used the All of Us Controlled Tier dataset release 8. Access to both resources is subject to the respective programmes’ approval procedures and data-use requirements, and individual-level data cannot be redistributed by the authors.

Gene-level association results from the phenome-wide disease-onset analyses and multistate disease-trajectory analyses in UK Biobank and All of Us can be explored through the GATE-GENE Browser (https://gategene.org). All All of Us-derived results are disseminated in accordance with the All of Us Data and Statistics Dissemination Policy, including restrictions on reporting participant counts of 1-20 or statistics from which such counts could be inferred.

Publicly available resources used in this study include GeneBass association summary statistics (https://app.genebass.org), Molecular Signatures Database (MSigDB) gene sets (https://www.gsea-msigdb.org/gsea/msigdb/) and PheCode mappings (https://phewascatalog.org).

## Code availability

GATE-GENE is implemented within the SAIGE R package and is publicly available under an open-source license at https://github.com/ZhouLabGenetics/GATE-GENE. Installation instructions, input and output specifications, and usage examples are available at https://zhoulabgenetics.github.io/GATE-GENE-doc/. The repository includes example data and a reproducible workflow for fitting a survival null model and performing gene-based rare-variant association tests.

UK Biobank phenome-wide onset analyses were performed using SAIGE v1.5.0.2, whereas the multistate trajectory analyses in UK Biobank and All of Us used v1.5.2. The corresponding Docker images were wzhou88/saige:1.5.0.2 and wzhou88/saigetest:1.5.2, respectively.

## Data Availability

Individual-level genetic and health-record data from UK Biobank and the All of Us Research Program are available to approved researchers through their respective controlled-access procedures and cannot be redistributed by the authors. UK Biobank data are accessible through the UK Biobank Access Management System, and All of Us data are available through the All of Us Researcher Workbench, subject to applicable access requirements. Aggregate association results and supporting analyses are provided in the manuscript and supplementary materials.

https://www.ukbiobank.ac.uk/use-our-data/apply-for-access/

## Acknowledgements

We gratefully acknowledge All of Us and UK Biobank participants for their contributions, without whom this research would not have been possible. Y.W. is supported by NIH-NHGRI R00HG013969. W.Z. is supported by NIH-NHGRI R00HG012222 and R01HG014518.

## Extended Data Figures

**Extended Data Fig. 1.**
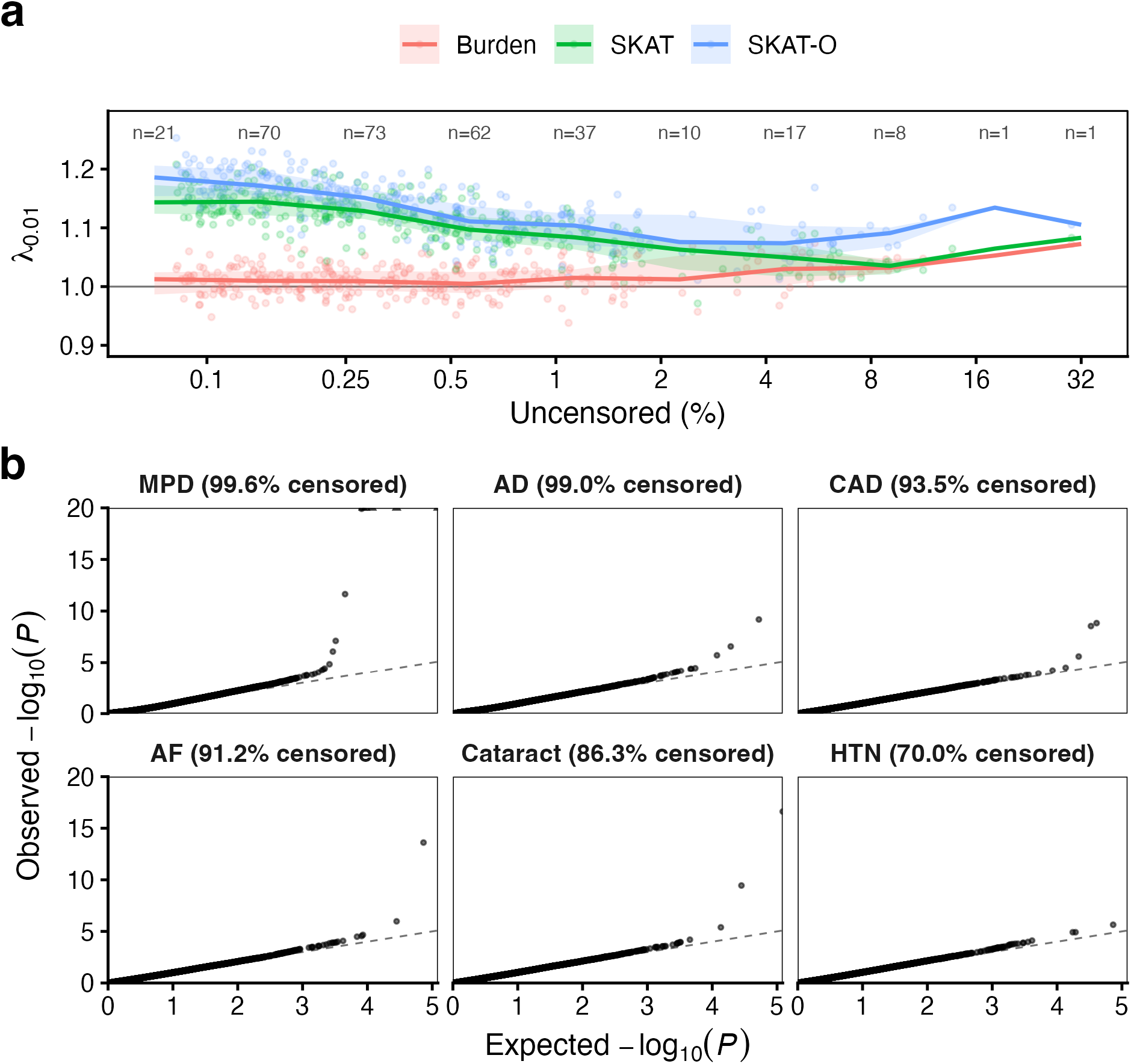
Calibration of phenome-wide gene-based time-to-event association testing in UK Biobank. **a)** Tail-based genomic control factor (*λ*_0.01_) for gene-level tests as a function of event information across the 300 core incident-onset phenotypes in UKB-EUR participants. Each point represents a phenotype-specific *λ*_0.01_ computed from gene-level *P*-values for the Cauchy-combined test, shown separately for Burden, SKAT and SKAT-O. The x-axis shows the uncensored proportion (100 − censoring rate; log scale). Solid lines and shaded bands denote the binned median and interquartile range of *λ*_0.01_ within uncensored-percentage bins; numbers above the panel indicate the number of phenotypes per bin. The horizontal line marks *λ*_0.01_ = 1. **b)** Quantile-quantile (QQ) plots for representative onset endpoints spanning a range of censoring proportions. Expected and observed -log₁₀(P) values are shown for the Cauchy-combined SKAT-O test. The dashed line denotes the null expectation. Observed values were capped at −log₁₀(*P*) = 20 for visualization; points exceeding the cap are indicated by triangles. Phenotype abbreviations: AD, Alzheimer’s disease; AF, atrial fibrillation; CAD, coronary artery disease; HTN, essential hypertension; MPD, myeloproliferative disease.

**Extended Data Fig. 2.**
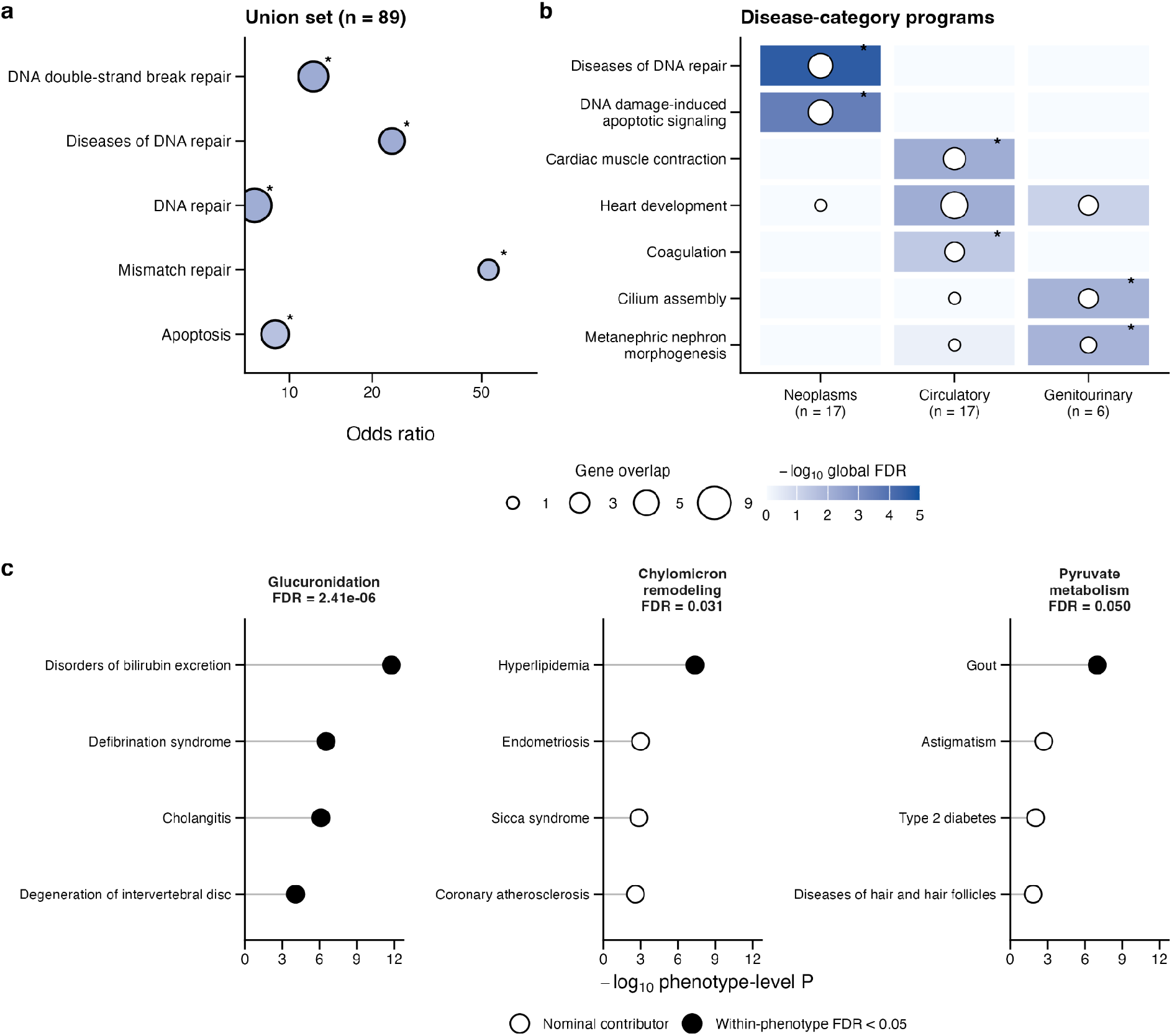
Functional convergence among onset-associated genes. a, Over-representation analysis of selected pathways among the union of 89 genes implicated by the 107 retained gene-phenotype associations. The x-axis shows the enrichment odds ratio on a logarithmic scale. Point area represents the number of overlapping genes and fill represents −log_10_(global FDR), calculated across the prespecified primary gene-list-pathway analyses. Asterisks indicate empirical global FDR < 0.05 in permutations using control genes matched on gene size, variant availability and testability. **b,** Enrichment of representative pathways among genes associated with neoplasm, circulatory-system and genitourinary-system phenotypes. Tile intensity represents −log_10_(global FDR) and point area represents the number of overlapping genes. Asterisks indicate empirical global FDR < 0.05 in matched-gene permutation analyses. Category sizes are shown below each column. **c,** Leading phenotype-level contributors to pathways identified by the ranked gene-set analysis across the 300 core phenotypes. Pathway-level FDR values were obtained after combining phenotype-specific evidence using Cauchy combination and correcting across all tested pathways. The x-axis shows the phenotype-level ranked-analysis *P* value. Filled circles indicate associations passing within-phenotype, within-collection FDR < 0.05; open circles indicate the remaining displayed contributors. The four strongest phenotype-level contributors are shown for each pathway.

**Extended Data Fig. 3.**
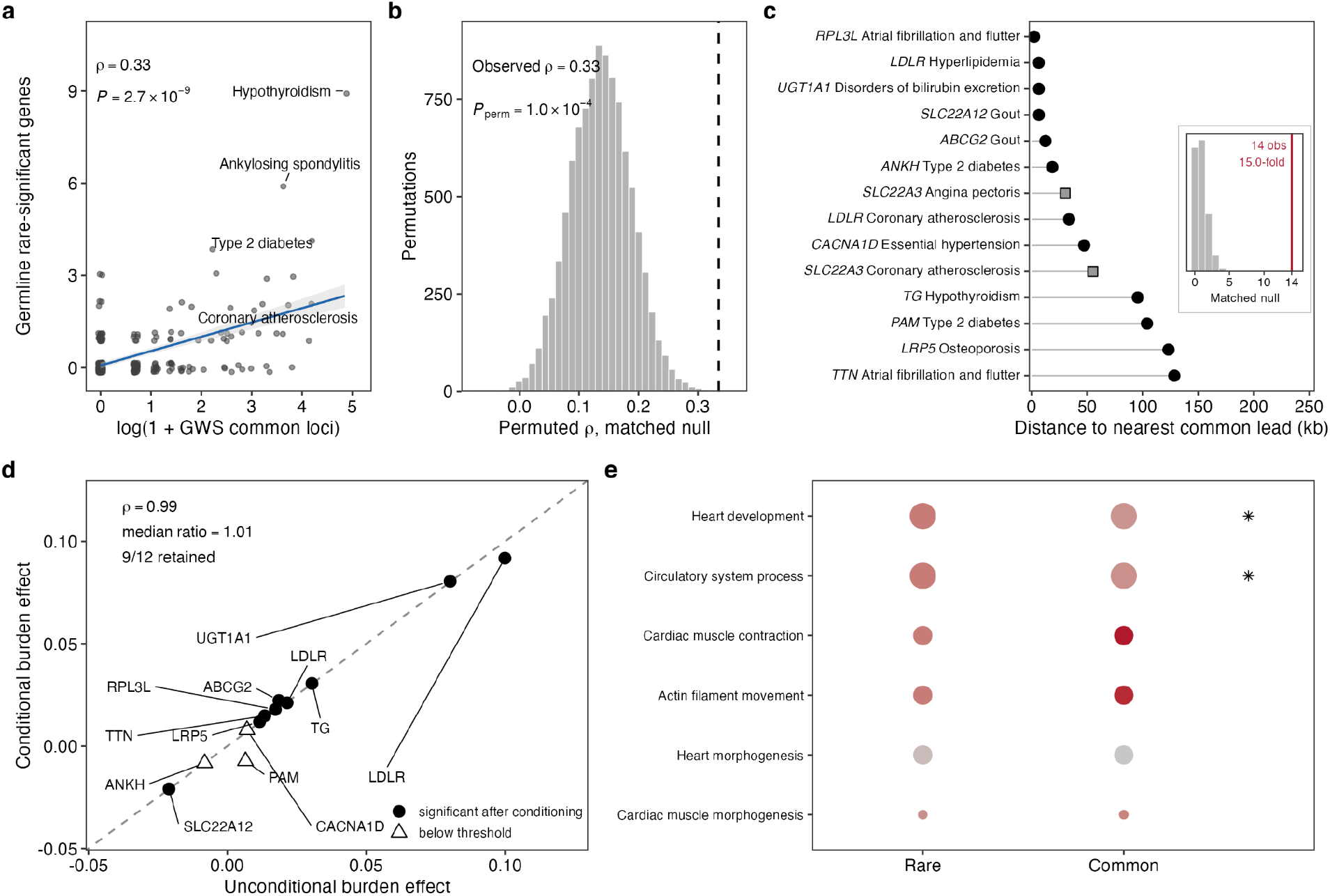
Overlap and functional convergence of rare- and common-variant associations with disease onset. a, Germline-likely rare-significant gene count against genome-wide significant clumped common-locus count across 300 incident-onset phenotypes; points are jittered, line is a linear fit with 95% confidence band. **b,** Null distribution of the Spearman correlation under 10,000 permutations of rare-gene counts within strata jointly defined by event-count quintile and censoring-rate tertile; dashed line, observed value. **c,** The 14 positional overlaps, showing the distance from each rare-significant gene to the nearest genome-wide significant common lead variant for the same phenotype. Symbols indicate whether the rare association remained exome-wide significant after conditioning on that lead (filled circle), fell below threshold (open triangle), or had an unstable conditional fit (grey square). Inset, matched permutation distribution of positional overlaps; red line, observed overlap count. **d,** Conditional against unconditional burden effect estimates for the 12 positionally overlapping gene-phenotype pairs with stable conditional fits; dashed line, identity. **e,** The six non-redundant pathway clusters shared between the rare- and common-variant analyses in circulatory phenotypes. Dot size indicates the number of contributing genes and colour indicates enrichment significance. Asterisks denote clusters retaining at least two genes after removing *TTN*, *MYH7* and *MYBPC3*; these three genes contributed 30 of 42 rare gene-term memberships.

**Extended Data Fig. 4.**
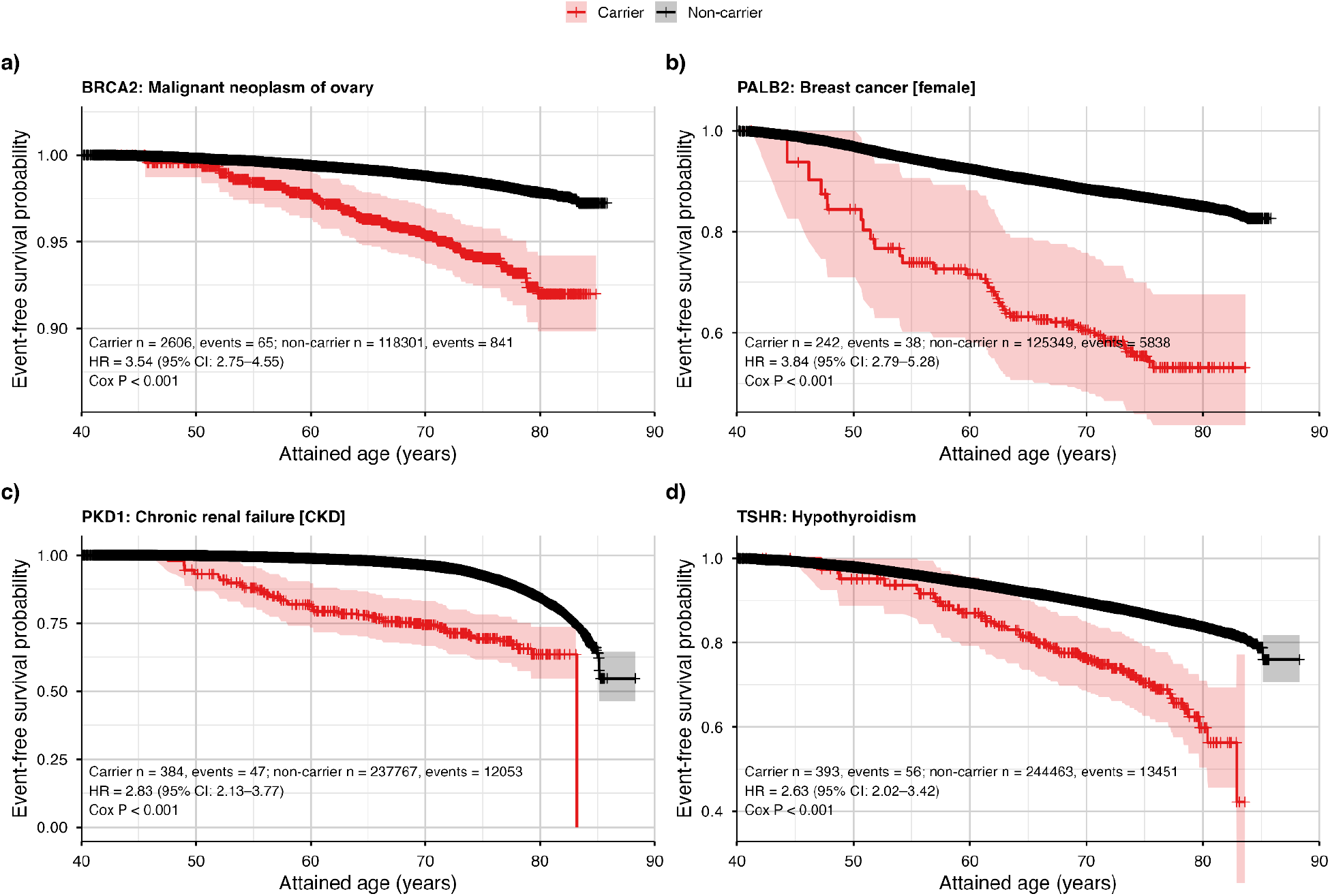
Representative attained-age delayed-entry survival curves stratified by rare-variant carrier status. Attained-age event-free survival curves are shown for **a**, *BRCA2*-malignant neoplasm of ovary; **b**, *PALB2*-breast cancer in females; **c**, *PKD1*-chronic renal failure; and **d**, *TSHR*-hypothyroidism. Red and black curves denote rare-variant carriers and non-carriers, respectively; shaded bands indicate 95% confidence intervals, and tick marks denote censoring. Curves were estimated using attained age as the time scale with delayed entry at cohort enrolment. Hazard ratios, 95% confidence intervals and two-sided Wald-test *P* values are from Cox proportional-hazards models including carrier status and available covariates for sex, birth year and the first 20 genetic principal components. Numbers of participants and incident events in each carrier group are shown within each panel.

**Extended Data Fig. 5.**
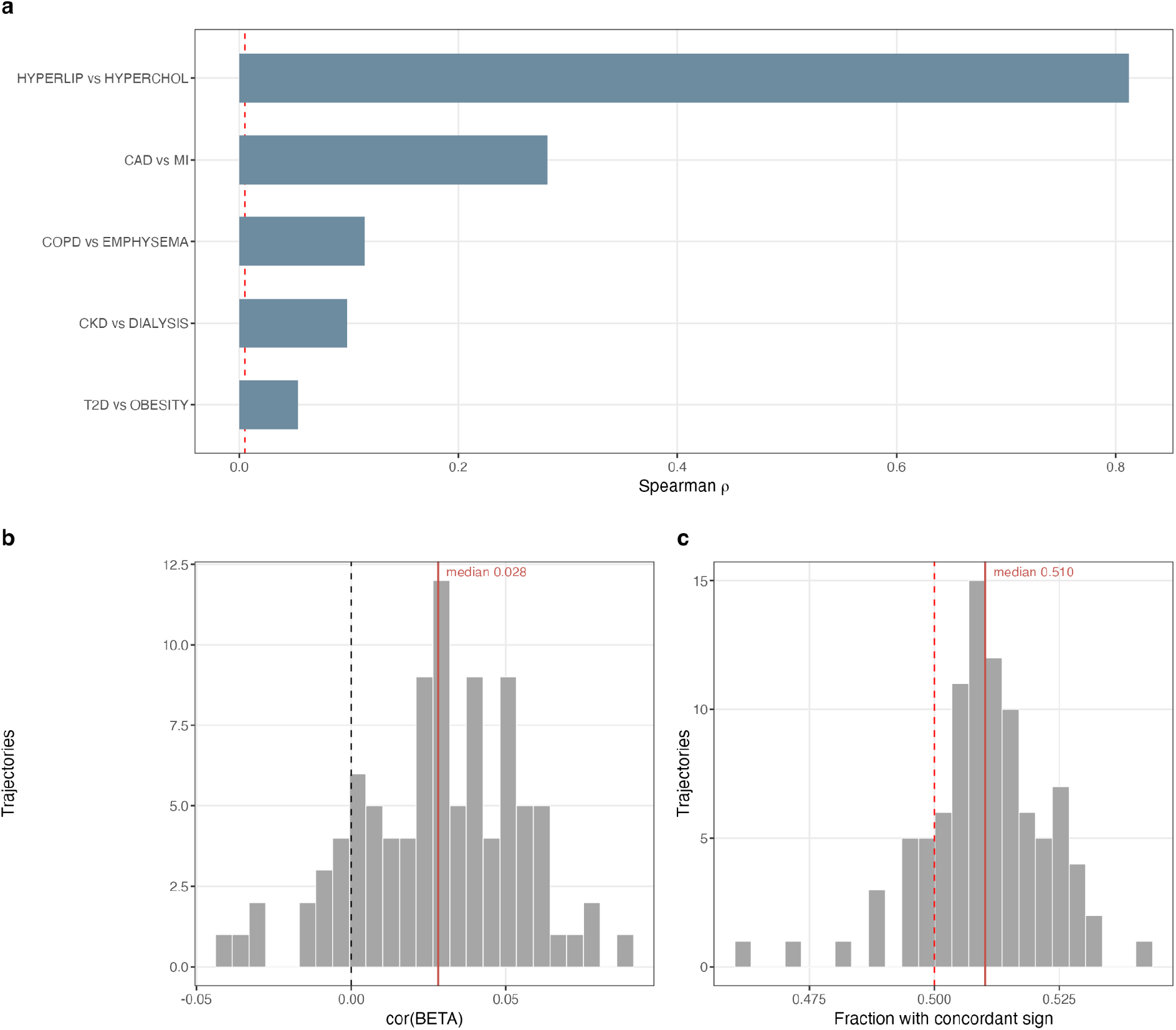
Controls and complementary measures of gene-level association concordance. a, Gene-level Spearman correlations for positive-control pairs of related onset phenotypes with expected shared association signals; the dashed red line indicates the median correlation among disjoint-pair controls. **b,** Distribution of gene-level effect-size correlation between T1→T2 and T0→T1 across trajectories; red line, median. **c,** Fraction of genes with concordant effect sign; red dashed line, 0.5 expected under independence; red line, median.

**Extended Data Fig. 6.**
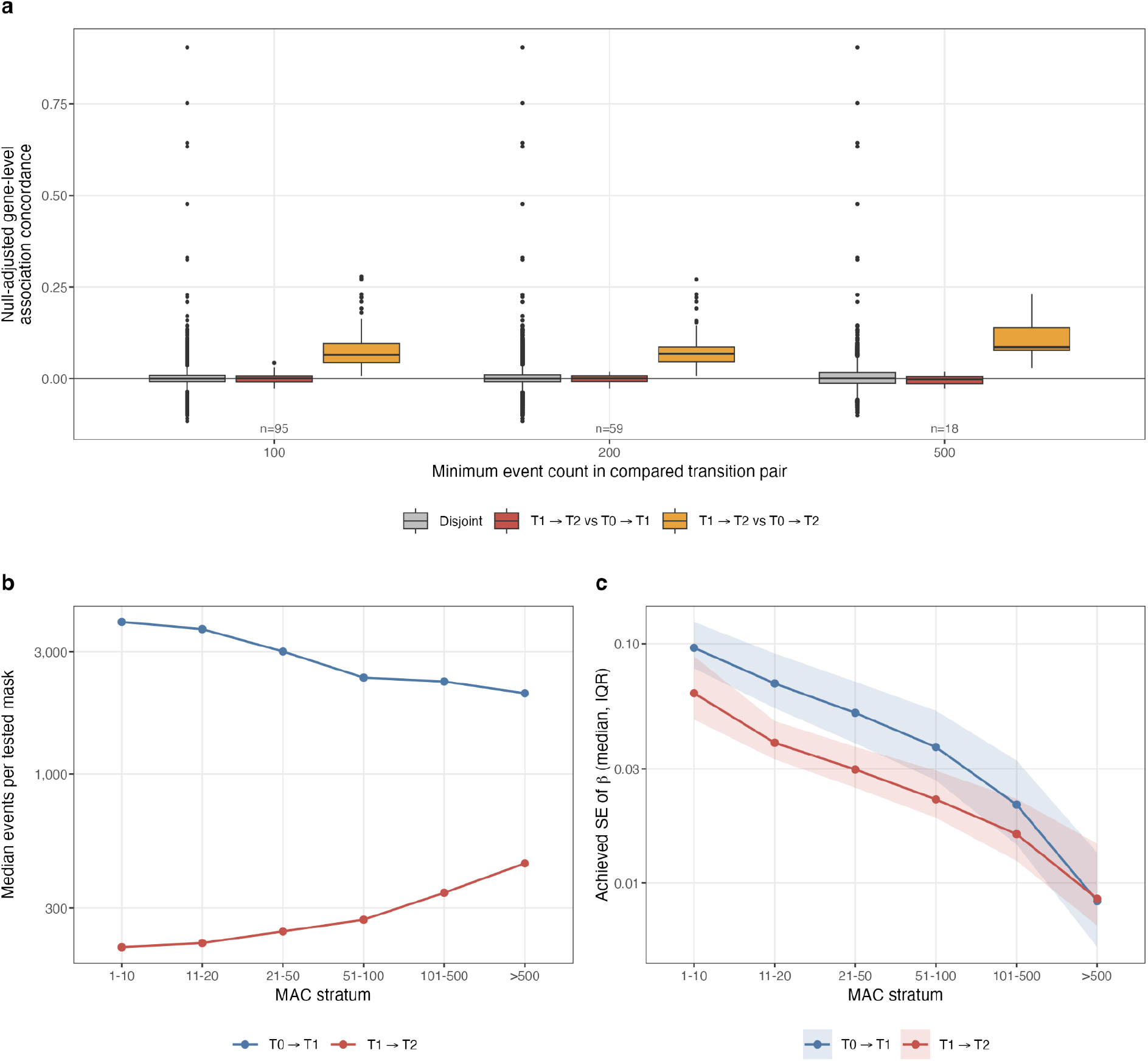
Statistical information and sensitivity of rare-variant progression analyses. a, Null-adjusted gene-level association concordance for transition pairs with minimum event counts of 100, 200 and 500, showing the distribution across comparison pairs. n above the axis gives T1→T2 vs T0→T1 pairs at each threshold. **b,** Median events per tested gene-mask by minor-allele-count stratum and transition class. **c,** Minimum detectable |*β*| by MAC stratum, estimated from achieved standard errors; shaded band, interquartile range across masks. Note that MAC is an allele COUNT: because progression risk sets are roughly twenty-fold smaller, the same MAC stratum corresponds to a much higher allele frequency in T1→T2 than in T0→T1, so panel b is not a like-for-like power comparison between transition classes and should be read within a class, not across. Panel a gives the between-class comparison.

**Extended Data Fig. 7.**
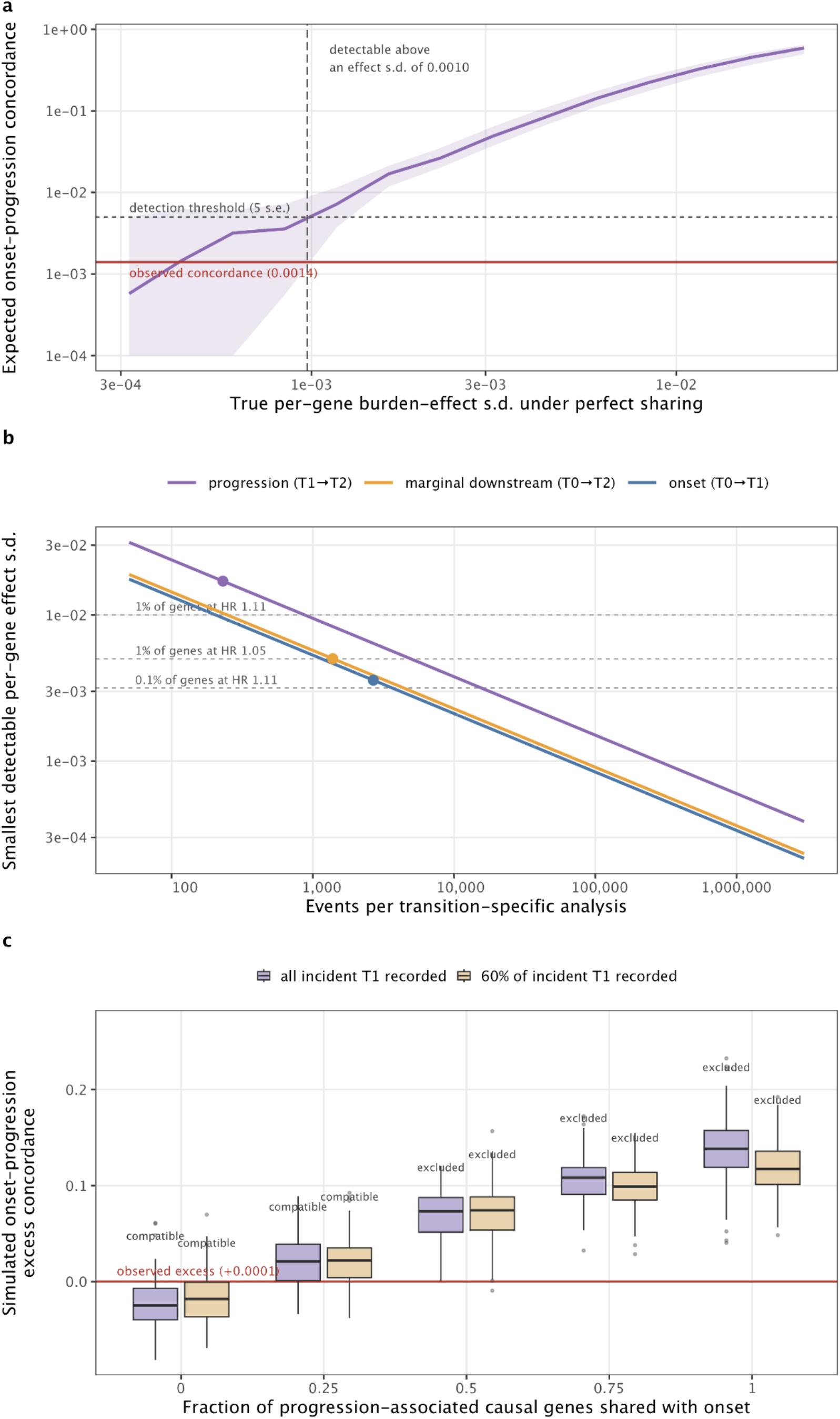
Detectability of shared onset-progression effects. a, Concordance expected under perfect sharing as a function of the true per-gene burden-effect s.d. **b,** Smallest detectable per-gene effect s.d. as a function of events per transition with the observed transitions marked. The reference line in a represents observed unadjusted onset-progression concordance (*ρ* = 0.0014), whereas that in c represents null-adjusted concordance (0.0001). **c,** Partial-sharing continuum. Box plots show simulated null-adjusted onset-progression concordance across 95 replicates per cell, for true shared-gene fractions of 0, 0.25, 0.5, 0.75 and 1, under complete and 60% ascertainment of incident T1. Boxes give the median and interquartile range, whiskers 1.5× the interquartile range, and the red line the observed value (+0.0001). A sharing fraction is labelled excluded where fewer than 5% of replicates yield an excess at or below the observed value. Under the evaluated simulation assumptions, sharing fractions up to 0.25 were compatible with the observed concordance, whereas sharing fractions of 0.5 or greater produced values at or below the observed estimate in fewer than 5% of replicates.

**Extended Data Fig. 8.**
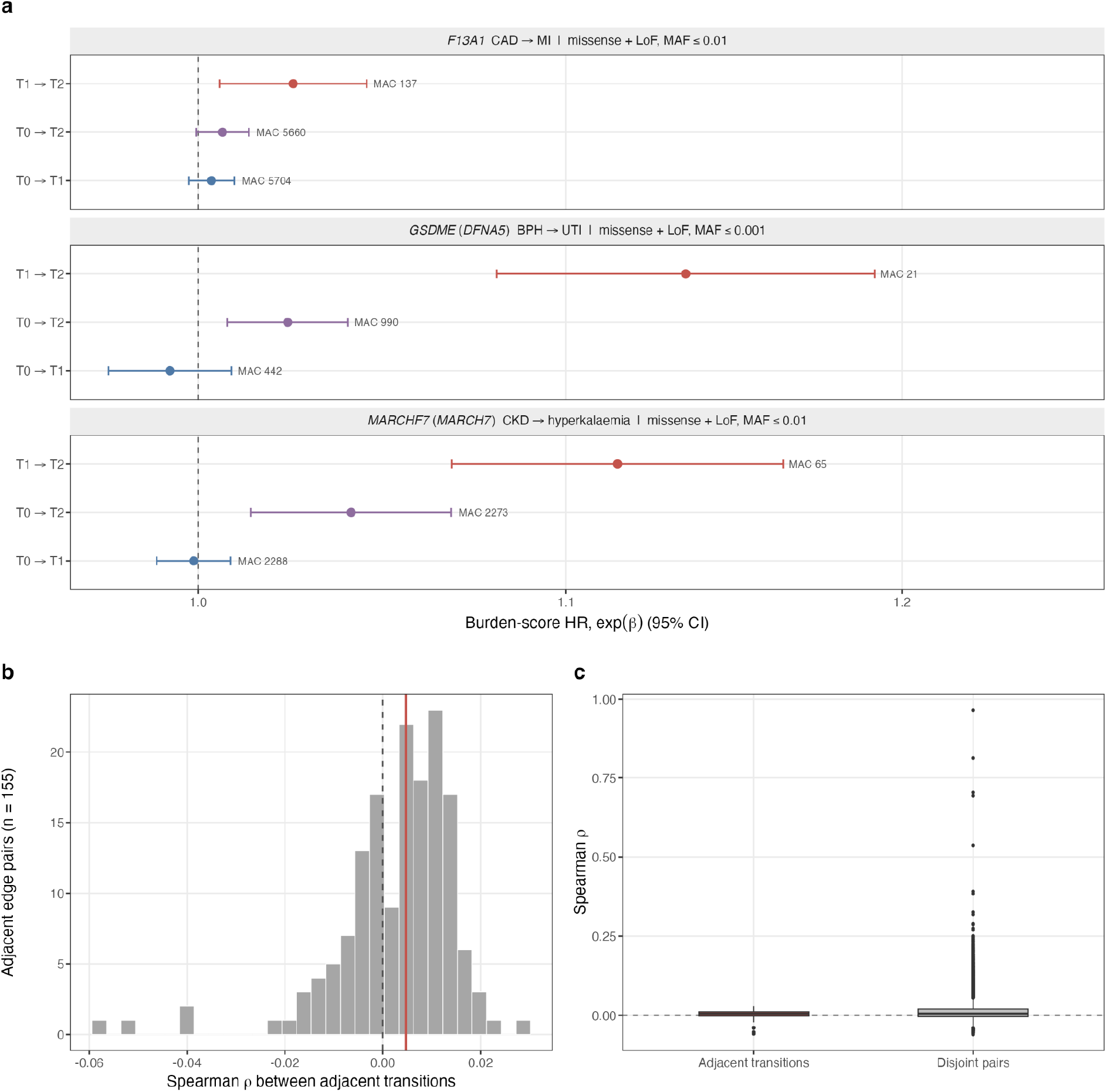
Rare-variant progression associations and concordance across longer clinical chains. a, Burden-score hazard ratios with 95% confidence intervals for the selected variant mask of each UKB exome-wide significant progression association, evaluated using the same mask in the onset, marginal downstream and progression analyses of the corresponding trajectory. The F13A1 association was identified through a variance-component test; its burden-score hazard ratio is displayed for descriptive comparison. **b,** Distribution of gene-level concordance between adjacent transitions in chains of two to five edges; dashed line, zero; red line, median. **c,** The same adjacent-edge concordance against disjoint pairs, which share no disease state, as a non-adjacent baseline.

**Extended Data Fig. 9.**
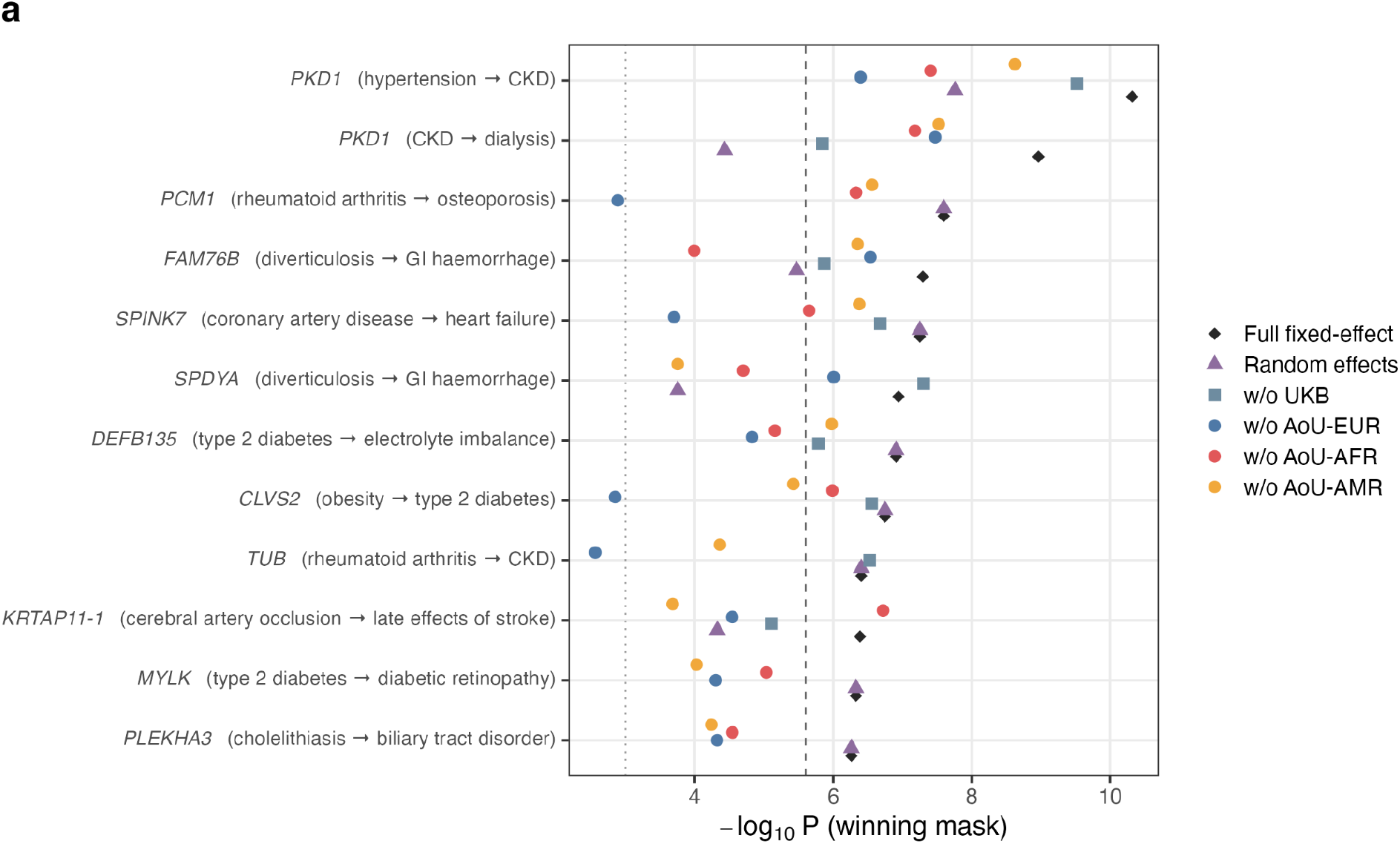
Sensitivity analyses of 12 additional cross-biobank progression associations. a, −log₁₀ *P* at the selected mask for each of the 12 associations under the full fixed-effect UKB+AoU meta-analysis (diamonds), the DerSimonian-Laird random-effects estimate (triangles) and each leave-one-stratum-out fixed-effect meta-analysis (squares and circles, by omitted stratum). Among the 12 additional candidates, *PLEKHA3* was considered exploratory because of sparse rare-variant support. Excluding this association, all 11 remaining candidates retained nominal significance under random-effects meta-analysis, with seven remaining exome-wide significant. The dashed line marks exome-wide significance (*P* = 2.5 × 10^-6^) and the dotted line *P* = 1 × 10^-3^. Leave-one-out estimates retained *P* < 1 × 10⁻³ for every omitted stratum in 9 of 12 associations (maximum leave-one-out *P* = 2.7 × 10^-3^ , on omission of AoU-EUR); random-effects estimates remained at *P* < 1 × 10⁻⁴ for 11 of 12. At the thresholds quoted in the main text, every association remained nominally significant (*P* < 0.05) under every omission, 4 to 8 of 12 remained exome-wide significant depending on which stratum was omitted, and the random-effects estimates retained all 12 at *P* < 0.05 with 8 exome-wide significant. **b,** Burden-score log-hazard ratios (points) with 95% confidence intervals (horizontal lines) at the selected mask for UKB-EUR, AoU-EUR, AoU-AFR and AoU-AMR, with the UKB+AoU fixed-effect meta-analytic estimate (diamonds), for progression from hypertension to chronic kidney disease and from chronic kidney disease to dialysis.

**Extended Data Fig. 10.**
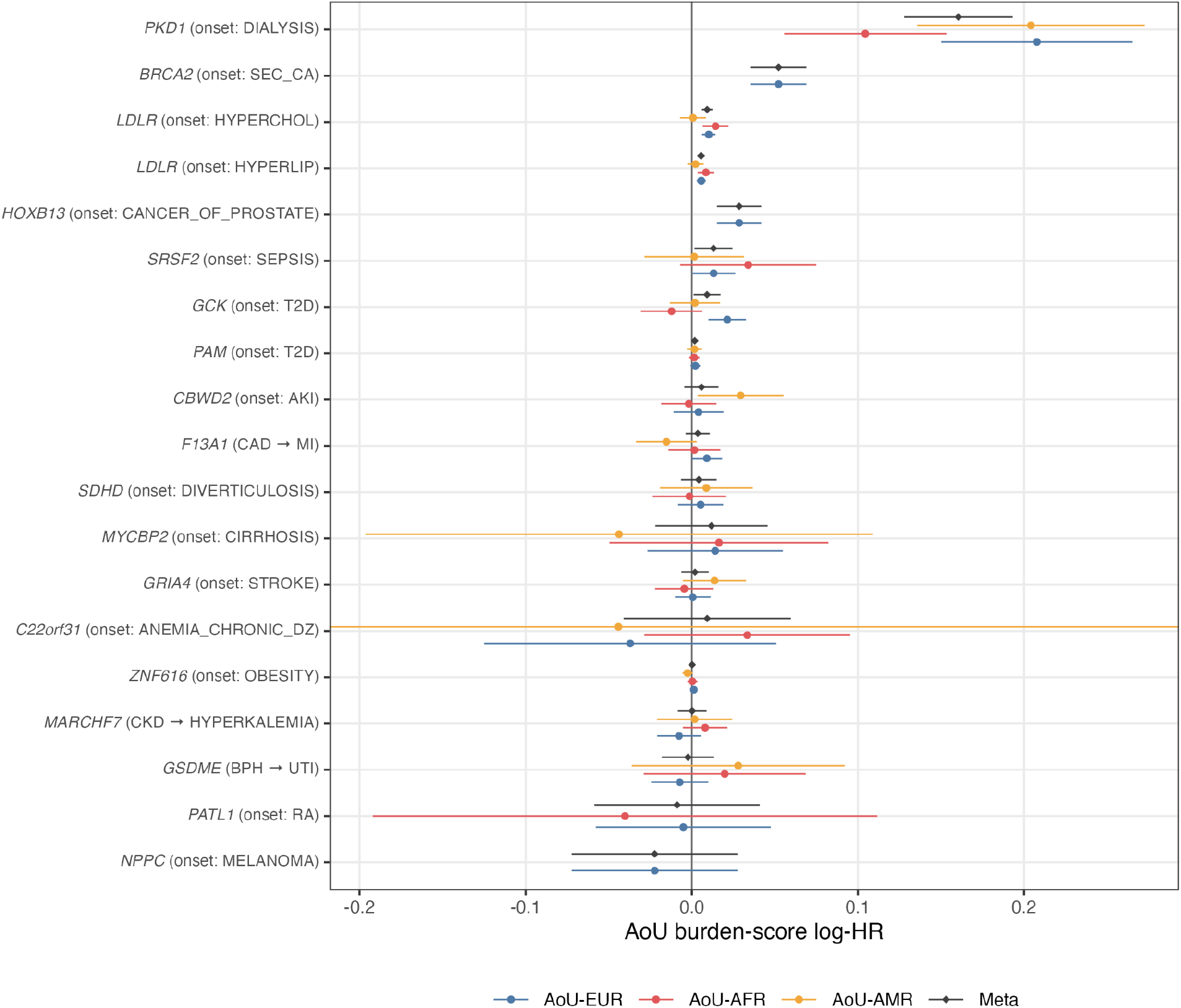
Cross-ancestry evaluation of 20 distinct UK Biobank exome-wide associations in All of Us. Burden-score log-hazard ratios (points) with 95% confidence intervals (horizontal lines) for 20 distinct gene-phenotype associations identified in UK Biobank (UKB) trajectory analyses and evaluated in the All of Us Research Program (AoU). Of these, 19, 16 and 15 associations were testable in the European (EUR), African (AFR) and Admixed American (AMR) ancestry groups, respectively. Diamonds indicate fixed-effect meta-analytic estimates across AoU ancestry groups. Effects are oriented to the UKB discovery direction, and associations are ordered by cross-ancestry meta-analysis *P* value. Associations not testable in a given ancestry group are omitted.

## References

1. Jack, C. R., Jr et al. Hypothetical model of dynamic biomarkers of the Alzheimer’s pathological cascade. Lancet Neurol. 9, 119–128 (2010).

2. Hanahan, D. & Weinberg, R. A. Hallmarks of cancer: the next generation. Cell 144, 646–674 (2011).

3. Yang, Z. et al. Limited overlap between genetic effects on disease susceptibility and disease survival. Nat. Genet. 1–9 (2025).

4. Liu, G. et al. Genome-wide survival study identifies a novel synaptic locus and polygenic score for cognitive progression in Parkinson’s disease. Nat. Genet. 53, 787–793 (2021).

5. Lee, J. C. et al. Genome-wide association study identifies distinct genetic contributions to prognosis and susceptibility in Crohn’s disease. Nat. Genet. 49, 262–268 (2017).

6. Urbut, S. M. et al. A Bayesian framework for longitudinal EHR and genetic discovery. Nature 1–12 (2026).

7. He, L. & Kulminski, A. M. Fast algorithms for conducting large-scale GWAS of age-at-onset traits using Cox mixed-effects models. Genetics 215, 41–58 (2020).

8. Bi, W., Fritsche, L. G., Mukherjee, B., Kim, S. & Lee, S. A Fast and Accurate Method for Genome-Wide Time-to-Event Data Analysis and Its Application to UK Biobank. Am. J. Hum. Genet. 107, 222–233 (2020).

9. Dey, R. et al. Efficient and accurate frailty model approach for genome-wide survival association analysis in large-scale biobanks. Nat. Commun. 13, 5437 (2022).

10. Pedersen, E. M. et al. ADuLT: An efficient and robust time-to-event GWAS. Nat. Commun. 14, 5553 (2023).

11. Karczewski, K. J. et al. Systematic single-variant and gene-based association testing of thousands of phenotypes in 394,841 UK Biobank exomes. Cell Genom. 2, 100168 (2022).

12. Backman, J. D. et al. Exome sequencing and analysis of 454,787 UK Biobank participants. Nature 599, 628–634 (2021).

13. Song, S., Li, X., Zhou, H., Li, Z. & Lin, X. Scalable and accurate rare-variant association tests for whole genome sequencing time-to-event analysis in large biobanks. Proc. Natl. Acad. Sci. U. S. A. 123, e2525288123 (2026).

14. Urbut, S. M. et al. MSGene: a multistate model using genetic risk and the electronic health record applied to lifetime risk of coronary artery disease. Nat. Commun. 15, 4884 (2024).

15. Tassistro, E., Bernasconi, D. P., Rebora, P., Valsecchi, M. G. & Antolini, L. Modeling the hazard of transition into the absorbing state in the illness-death model. Biom. J. 62, 836–851 (2020).

16. Siggaard, T. et al. Disease trajectory browser for exploring temporal, population-wide disease progression patterns in 7.2 million Danish patients. Nat. Commun. 11, 1–10 (2020).

17. Jensen, A. B. et al. Temporal disease trajectories condensed from population-wide registry data covering 6.2 million patients. Nat. Commun. 5, 1–10 (2014).

18. Westergaard, D., Moseley, P., Sørup, F. K. H., Baldi, P. & Brunak, S. Population-wide analysis of differences in disease progression patterns in men and women. Nat. Commun. 10, 1–14 (2019).

19. Hougaard, P. Frailty models for survival data. Lifetime Data Anal. 1, 255–273 (1995).

20. Daniels, H. E. Saddlepoint Approximations in Statistics. Ann. Math. Stat. 25, 631– 650 (1954).

21. Zhou, W. et al. Efficiently controlling for case-control imbalance and sample relatedness in large-scale genetic association studies. Nat. Genet. 50, 1335–1341 (2018).

22. Wu, M. C. et al. Rare-variant association testing for sequencing data with the sequence kernel association test. Am. J. Hum. Genet. 89, 82–93 (2011).

23. Lee, S., Wu, M. C. & Lin, X. Optimal tests for rare variant effects in sequencing association studies. Biostatistics 13, 762–775 (2012).

24. Zhou, W. et al. SAIGE-GENE+ improves the efficiency and accuracy of set-based rare variant association tests. Nat. Genet. 54, 1466–1469 (2022).

25. Zhou, W. et al. Scalable generalized linear mixed model for region-based association tests in large biobanks and cohorts. Nat. Genet. 52, 634–639 (2020).

26. van der Windt, G. J. W., Hoogerwerf, J. J., de Vos, A. F., Florquin, S. & van der Poll, T. Osteopontin promotes host defense during Klebsiella pneumoniae-induced pneumonia. Eur. Respir. J. 36, 1337–1345 (2010).

27. Sunryd, J. C. et al. TMTC1 and TMTC2 are novel endoplasmic reticulum tetratricopeptide repeat-containing adapter proteins involved in calcium homeostasis. J. Biol. Chem. 289, 16085–16099 (2014).

28. Putter, H., Fiocco, M. & Geskus, R. B. Tutorial in biostatistics: competing risks and multi-state models. Stat. Med. 26, 2389–2430 (2007).

29. Andersen, P. K. & Keiding, N. Multi-state models for event history analysis. Stat. Methods Med. Res. 11, 91–115 (2002).

30. Palmer, D. S. et al. The Biobank Rare Variant consortium powers the discovery of rare genetic associations through global collaboration. medRxiv (2026) doi:10.64898/2026.05.21.26353759.

31. Bycroft, C. et al. The UK Biobank resource with deep phenotyping and genomic data. Nature 562, 203–209 (2018).

32. Kar, S. P. et al. Genome-wide analyses of 200,453 individuals yield new insights into the causes and consequences of clonal hematopoiesis. Nat. Genet. 54, 1155–1166 (2022).

33. All of Us Research Program Investigators et al. The “All of Us” Research Program. N. Engl. J. Med. 381, 668–676 (2019).

34. Lu, W. et al. Systematic common and rare variant association testing in 392,030 whole genomes in All of Us. medRxiv (2026) doi:10.64898/2026.05.08.26350964.

35. Wu, P. et al. Mapping ICD-10 and ICD-10-CM Codes to Phecodes: Workflow Development and Initial Evaluation. JMIR Med Inform 7, e14325 (2019).

36. Vaupel, J. W., Manton, K. G. & Stallard, E. The impact of heterogeneity in individual frailty on the dynamics of mortality. Demography 16, 439–454 (1979).

37. Dey, R., Schmidt, E. M., Abecasis, G. R. & Lee, S. A fast and accurate algorithm to test for binary phenotypes and its application to PheWAS. Am. J. Hum. Genet. 101, 37– 49 (2017).

38. Kuonen, D. Miscellanea. Saddlepoint approximations for distributions of quadratic forms in normal variables. Biometrika 86, 929–935 (1999).

39. Barndorff-Nielsen, O. E. Approximate interval probabilities. J. R. Stat. Soc. Series B Stat. Methodol. 52, 485–496 (1990).

40. Li, X. et al. Dynamic incorporation of multiple in silico functional annotations empowers rare variant association analysis of large whole-genome sequencing studies at scale. Nat. Genet. 52, 969–983 (2020).

41. Liu, Y. & Xie, J. Cauchy combination test: a powerful test with analytic p-value calculation under arbitrary dependency structures. J. Am. Stat. Assoc. 115, 393–402 (2020).

42. Benjamini, Y. & Hochberg, Y. Controlling the false discovery rate: A practical and powerful approach to multiple testing. J. R. Stat. Soc. Series B Stat. Methodol. 57, 289–300 (1995).

43. James, C. et al. A unique clonal JAK2 mutation leading to constitutive signalling causes polycythaemia vera. Nature 434, 1144–1148 (2005).

44. Meggendorfer, M. et al. SRSF2 mutations in 275 cases with chronic myelomonocytic leukemia (CMML). Blood 120, 3080–3088 (2012).

45. Montes-Moreno, S. et al. Clinical molecular testing for ASXL1 c.1934dupG p.Gly646fs mutation in hematologic neoplasms in the NGS era. PLoS One 13, e0204218 (2018).

46. Jaiswal, S. et al. Clonal hematopoiesis with somatic mutations is a common, age-related condition associated with adverse outcomes. Blood 124, 840–840 (2014).

47. Klco, J. M. & Mullighan, C. G. Advances in germline predisposition to acute leukaemias and myeloid neoplasms. Nat. Rev. Cancer 21, 122–137 (2021).

48. Khoury, J. D. The 5th edition of the WHO Classification of Haematolymphoid Tumours: Myeloid and Histiocytic/Dendritic Neoplasms. Leukemia.

49. Horton, R. et al. Gene map of the extended human MHC. Nat. Rev. Genet. 5, 889– 899 (2004).

50. Genovese, G. et al. Clonal hematopoiesis and blood-cancer risk inferred from blood DNA sequence. N. Engl. J. Med. 371, 2477–2487 (2014).

51. Liberzon, A. et al. The Molecular Signatures Database (MSigDB) hallmark gene set collection. Cell Syst. 1, 417–425 (2015).

52. Gillespie, M. et al. The reactome pathway knowledgebase 2022. Nucleic Acids Res. 50, D687–D692 (2022).

53. The Gene Ontology Consortium. The Gene Ontology resource: enriching a GOld mine. *Nucleic Acids Research* 49, D325–D334 (2021).

54. GTEx Consortium. The GTEx Consortium atlas of genetic regulatory effects across human tissues. Science 369, 1318–1330 (2020).

55. Chang, C. C. et al. Second-generation PLINK: rising to the challenge of larger and richer datasets. Gigascience 4, 7 (2015).

56. de Leeuw, C. A., Mooij, J. M., Heskes, T. & Posthuma, D. MAGMA: generalized gene-set analysis of GWAS data. PLoS Comput. Biol. 11, e1004219 (2015).

