## Supplementary Notes and Figures for "Exome-wide rare-variant associations vary across disease trajectories"

#### Supplementary Note 1. GATE-GENE statistical framework

##### Frailty model

In a study with sample size $N$, the phenotype of the $i$th individual is denoted by the pair $(\delta_i, t_i)$, where $\delta_i$ is the censoring indicator, with 1 for event and 0 for censored. The time $t_i$ represents the observed event or censoring time. Let $X_i$ represent $p$ covariates, $G_i$ represent the allele counts (0, 1, or 2 ) for $q$ variants to be tested. The conditional hazard function in the frailty model[52] can be written as, where $\lambda_0(t)$ is the baseline hazard function at time $t$, which works as the intercept in a frailty model. $b_i$ is the Gaussian frailty following $MVN(0, \tau\psi)$, where $\psi$ is $N \times N$ genetic relationship matrix (GRM), and $\tau$ is the additive genetic variance parameter. $\alpha$ is a coefficient vector of fixed effects and $\beta$ is a coefficient vector of the genetic effect.

##### Estimate variance component and other model parameters (Step 1)

Same as in the original GATE[9], to fit the null frailty model in GATE-GENE, the likelihood for the event-time data pairs $(\delta, t)$ can be expressed as a modified Poisson mixed effects model (GLMM)[53] likelihood with a weighted mean function. Suppose there are $Z$ unique event times: $t_1^* < t_2^* < \ldots < t_Z^*$ in increasing order. The mean function is weighted by the cumulative baseline hazard (CBH) estimated using the Breslow's estimator[54] $\hat{\Lambda}_0(t) = \sum_{i=1}^{N} \frac{\delta_i I(t_i \le t)}{\sum_{j \in R(t_i)} exp(\hat{\eta}_j)}$, where $I(t_i \le t)$ is indicator variable, $R(t_i)$ is the set of individuals at risk at $t_i$ and $\hat{\eta}_j = X^T{}_i \hat{\alpha} + G_i \hat{\beta} + \hat{b}_i$ . Thus, under the null hypothesis $\beta = 0$, penalized quasi-likelihood (PQL)[55] method and the computational efficient average information restricted maximum likelihood (AI-REML)[56] algorithm are used to iteratively estimate $(\hat{\tau}, \hat{\alpha}, \hat{b})$. At iteration $k$, let $(\hat{\tau}^{(k)}, \hat{\alpha}^{(k)}, \hat{b}^{(k)})$ be estimated $(\hat{\tau}, \hat{\alpha}, \hat{b})$, $\hat{\mu}_i{}^{(k)}$ be the estimated mean and $\hat{\mu}_i{}^{(k)} = \hat{\Lambda}_0(t) exp(X^T{}_i \hat{\alpha}^{(k)} + \hat{b}^{(k)})$ and $\hat{\Sigma}^{(k)} = \widehat{W}^{(k)^{-1}} + \hat{\tau}^{(k)}\psi$ be an $N \times N$ matric of the variance of working vector $\tilde{y}_i = X^T{}_i \hat{\alpha}^{(k)} + \hat{b}^{(k)} + (\delta_i - \hat{\mu}_i{}^{(k)}) / \{\hat{\mu}_i{}^{(k)}\}$ and $\widehat{W}^{(k)} = diag(\hat{\mu}_i{}^{(k)})$. To obtain the log quasi-likelihood and average information at each iteration, GATE and GATE-GENE use the preconditioned conjugate gradient approach (PCG)[57] to obtain the product of inverse of $\hat{\Sigma}^{(k)}$ and any other vector by iteratively solving a linear system with $\hat{\Sigma}^{(k)}$.

##### Set-based association tests (Step 2)

Test statistics of the Burden, SKAT[30], and SKAT-O[31] tests for a variance set can be conducted based on score statistics from marginal models for individual variants in the set. Suppose there are $q$ variants in the set to test. The score statistics for variant $j$ $(j = 1, \ldots, q)$ under $H_0 : \beta_j = 0$ is $T_j = g_j{}^T(\delta - \hat{\mu})$, where $g_j$ and $\delta$ are $N \times 1$ genotype and phenotype (event status) vectors, respectively, and $\hat{\mu}$ is the estimated mean of $\delta$ under the null hypothesis.

Let $s_j$ denote a threshold indicator or weight for variant $j$ and $S = diag(s_1,\dots,s_q)$ be a diagonal matrix with $s_j$ as the $jth$ element. Following the original SKAT and SKAT-O methods and the SAIGE-GENE+ tool, to upweight rare variants, the default setting in GATE-GENE is $u_j = Beta(MAF_j, 1, 25)$, which upweights rarer variants. Let $\tilde{G} = G - \tilde{X}(\tilde{X}^T\widehat{W}\tilde{X})^{-1}\tilde{X}^T\widehat{W}G$ be the covariate-and-intercept adjusted genotype matrix, where $G = (g_1,\dots,g_q)$ is $N \times q$ genotype matrix for the $q$ variants, $\tilde{X} = [1\,X]$ is the intercept-augmented covariate matrix, $\hat{Q} = \hat{S}^{-1} - \hat{S}^{-1}X(X^T\hat{S}^{-1}X)^{-1}\hat{S}^{-1}$, and $\hat{S} = (\widehat{W} - \widehat{U})^{-1} + \hat{\tau}\Psi$. Note that as shown in the original GATE paper, $\widehat{U} = \Gamma R^T D R \Gamma$, which is a rank $Z$ matrix to account for the loss of information due to estimating the baseline hazard function. $D_{Z\times Z} = diag\{\frac{d_Z}{\sum_{j\in R(t_z*)} exp(\hat{\eta}_j)}\}$, where $z = 1,\dots,Z$ unique event times and $d_z = \sum_{j:t_j=t_z} \delta_j$ is the number of events at time $t_z$. $\Gamma_{N\times N} = diag\{exp(\hat{\eta}_i)\}$, where $i = 1,\dots,N$ individuals, and $R$ is an $Z \times N$ matrix with $R_{zi} = 1$ if $i \in R(t_z*)$ and otherwise, 0.

The Burden and SKAT test statistics can be written as $T_{Burden} = (\sum_{j=1}^{q} s_j T_j)^2$ and $T_{SKAT} = \sum_{j=1}^{q} s_j{}^2 T_j{}^2$. Under the null hypothesis of no genetic effects, $T_{Burden}$ follows $\lambda_B \chi_1{}^2$, where $\lambda_B = J^T S\tilde{G}^T\hat{Q}\tilde{G}SJ$, $J$ is a $q \times 1$ vector with all elements being unity and $\chi_1{}^2$ is a chi-square distribution with 1 degree of freedom and $T_{SKAT}$ follows a mixture of chi-square distribution $\sum_{j=1}^{q} \lambda_{Sj}\chi_1{}^2$, where $\lambda_{Sj}$ is the $j$th eigenvalue of $S\tilde{G}^T\hat{Q}\tilde{G}S$. The SKAT-O test uses a linear combination of the Burden and SKAT test with test statistics $T_{SKAT-O} = (1-\rho)T_{SKAT} + \rho T_{Burden}$ with $0 \le \rho \le 1$. Following the SKAT-O test paper, we use a grid of eight values of $\rho = (0, 0.1^2, 0.2^2, 0.3^2, 0.4^2, 0.5^2, 0.5, 1)$ to find the minimum p-value for the SKAT-O test.

#### Incorporating multiple MAF thresholds and functional annotations

GATE-GENE allows users to incorporate multiple maximum MAF cutoffs and functional annotations in the set-based tests, which has been previously demonstrated to improve power for rare-variant tests of binary and quantitative phenotypes compared to a single MAF cutoff and functional annotation. Similar to SAIGE-GENE+, for each set (e.g. gene or region), SKAT-O p-values correlating to each mask of the combination of a MAF cutoff and a functional annotation are combined using the Cauchy combination method. Note that the Cauchy tes[36,58]t does not work well when one or more p-values equal to 1 or 0, which the minimum p-value

#### Approximate $\tilde{G}^T\hat{Q}\tilde{G}$

For each set of variants, given $\hat{Q}$, the calculation of $\tilde{G}^T\hat{Q}\tilde{G}$ requires PCG for each variant in the set, which can be computationally very expensive. Suppose $\tilde{g}$ is the covariate-and-intercept adjusted genotype vector for a single variants, an approximation approach has been successfully used in GATE to approximate $\tilde{g}^T\widehat{Q}\tilde{g}$ as the product of $\tilde{g}^T\widehat{W}\tilde{g}$ and the ratio estimated between the two using a small subset of randomly selected genetic markers, denoted as $\hat{r}$. This is because the ratio has been shown to be approximately constant for all variants both through theoretical proof and using the real data in the original GATE paper. Zhou et al.[17] have shown that the variance ratio may vary for ultra-rare variants with MAC < 10, so SAIGE-GENE estimates variance ratios for different MAC categories. Later SAIGE-GENE+[18] proposed collapsing ultra-rare variants with MAC < 10 and then testing the collapsed variant with other rare variants to reduce type I error and improve computation efficiency, which also enables a single variant ratio estimated using non-ultra rare variants to be used. GATE-GENE follows the SAIGE-GENE+ to first collapse ultra-rare variants, so $\tilde{G}^{T}\hat{Q}\tilde{G}$ can be calculated as $\hat{r}\tilde{G}^{T}\widehat{W}\tilde{G}$, where $\widehat{W}$ is a diagonal matrix and $\tilde{G}^{T}\widehat{W}\tilde{G}$ can be computed in the $O(N)$ instead of requiring $O(N^{1.5})$ to compute $\tilde{G}^{T}\hat{Q}\tilde{G}$. The saving of the computation can be substantial when the sample size is large, e.g. 500k samples in the UK Biobank.

#### Robust adjustment for $\hat{r}\tilde{G}^{T}\widehat{W}\tilde{G}$ to account for heavy censoring

To account for heavy censoring of time-to-event (TTE) phenotypes in set-based tests, GATE-GENE first applied saddlepoint approximation (SPA) to single variant score test statistics to obtain well calibrated p-values for each variant in the testing set following the approach used in GATE. After calculating p-values of $T_j$ for $j = 1, \ldots, q$ and the variance of $T_j$, denoted as $Var(T_j)_{woSPA} = \tilde{g}_j^{T}\widehat{Q\tilde{g}_j}$ as is calibrated by inverting the corresponding p-value to be $Var(T_j)_{SPA}$. Let $f_j = Var(T_j)_{SPA}/Var(T_j)_{woSPA}$ be the scaling factor for the $jth$ variant and $F$ is the $q \times 1$ vector with $f_j$ be the $jth$ element. GATE-GENE then adjust the $\tilde{G}^{T}\hat{Q}\tilde{G}$ based on $F$ to be $F^{1/2}(F^{1/2})^{T} \odot \tilde{G}^{T}\hat{Q}\tilde{G}$ to compute robust p-values for the set-based tests.

#### Collapsing ultra-rare variants

To fully address the heavy censoring of TTE phenotypes and the sparsity in the data in rare variants tests, GATE-GENE follows the similar approached used in SAIGE-GENE+ to collapse variants with MAC≤10 and then tests the collapsed variant together with all other variants with MAC>10.

### Supplementary Note 2. Simulation design and method benchmarking

We performed a series of simulation studies to evaluate the performance of GATE-GENE, with a focus on both type I error control and statistical power. To closely mimic the population structure and sample relatedness typical of large-scale biobank cohorts, we leveraged real genotype data from UKB. Specifically, we randomly selected 100,000 White British individuals with available exome sequencing data. Phenotypes were simulated based on a subset of $L = 100{,}000$ variants drawn from a pool of 250,656 directly genotyped and LD-pruned markers (window size = 500 bp, step-size = 50 bp, $r^2 < 0.2$). All selected markers had a minor allele frequency (MAF) ≥ 0.01 and were randomly sampled from the odd chromosomes.

These 100,000 markers were also used to construct a sparse genomic relationship matrix (GRM), applying a sparsity threshold of 0.05 to the pairwise relatedness coefficients across all analyses. For each phenotype set, the sparse GRM was used for variance ratio estimation after a null frailty model fitting in Step 1. In Step 2, we performed gene-based association testing using GATE-GENE under three maximum MAF thresholds (1%, 0.1% and 0.01%) and three variant inclusion criteria based on functional annotation: (i) loss-of-function (LoF) variants only, (ii) LoF plus missense variants; and (iii) LoF, missense, and synonymous variants. For each gene, association tests were performed using the Burden, SKAT and SKAT-O method. Results were then aggregated using either the Cauchy combination test [35,36] or minimum *P* value approach. Variant annotations were obtained using ANNOVAR[59]. The LoF variants include those annotated as frameshift deletions/insertions, non-frameshift deletions/insertions, splicing, stop-gain and stop-loss mutations.

#### Type I error rates simulations

Null phenotypes were simulated following the mixed model: $\lambda_i(t) = \lambda_0(t)exp(X_I\beta + G_i\gamma + b_i$. We selected $\lambda$, the mean of the exponential hazard function, corresponding to different censoring rates: 90%, 95% and 99%, respectively, and repeated the simulation for 20 times for each different censoring rate. Event times were generated from an exponential model. For each censoring level, 20 independent phenotype replicates were generated and analysed genome-wide. Null models included the simulated covariates and the first four genetic principal components.

Each replicate was analysed using GATE-GENE with and without saddlepoint approximation (SPA). Empirical type I error was evaluated for burden, SKAT, SKAT-O and the per-gene Cauchy omnibus statistic, including at the exome-wide threshold of $P = 2.5 \times 10^{-6}$ and at $P = 0.001$. Quantile-quantile plots were additionally used to evaluate departures from the expected null distribution across censoring levels and MAF thresholds.

Empirical type I error at the exome-wide threshold ($2.5 \times 10^{-6}$) was evaluated separately for every combination of test (burden, SKAT, SKAT-O), censoring level (90%, 95%, 99%) and maximum MAF threshold ($1 \times 10^{-4}$, $1 \times 10^{-3}$, $1 \times 10^{-2}$), giving 27 cells each based on approximately 463,000 gene-level tests. With saddlepoint approximation, empirical error ranged from 0 to $4.0 \times 10^{-6}$ at most 1.7-fold the nominal rate, and the exact binomial 95% confidence interval contained the nominal value in 27 of 27 cells. Without saddlepoint approximation the same cells showed inflation up to 60-fold (SKAT at 99% censoring, maximum MAF 0.001), confirming that the calibration gain is attributable to the saddlepoint step rather than to the aggregation scheme. Per-cell counts, empirical rates and confidence intervals are released with the analysis code.

#### GATE-STAAR calibration comparison

To determine whether calibration under extreme censoring differed for an existing gene-based rare-variant time-to-event framework, we applied GATE-STAAR to the same simulated null phenotypes used for the GATE-GENE calibration analyses. The comparison was restricted to chromosome 19 because the GATE-STAAR workflow required method-specific genotype conversion and substantially larger intermediate files. Within chromosome 19, the two methods were evaluated using the same participants, phenotype replicates, censoring mechanisms, covariates, eligible variants and tested genes wherever the corresponding variant masks could be matched.

We evaluated loss-of-function (LoF), missense and synonymous variant groups at maximum minor-allele-frequency (MAF) thresholds of 1%, 0.1% and 0.01%, spanning a range of variant sparsity. For each of the 90%, 95% and 99% censoring levels, 200 independently simulated null phenotype replicates were analysed. GATE-STAAR was run using release v0.1. Saddlepoint approximation was enabled only when the non-SPA asymptotic p-value is nominally significant under the evaluated conditions.

For burden and SKAT testing, GATE-STAAR used its implemented default ultra-rare-variant collapsing threshold of MAC ≤ 20 and Beta(MAF; 1,25) rare-variant weighting. The implemented threshold differs from the MAC threshold of 10 described in the original GATE-STAAR report; therefore, we retained the software implementation default throughout and reported this distinction explicitly. STAAR-O, the omnibus test implemented in GATE-STAAR, combined burden, SKAT and ACAT-V statistics across the weighting schemes implemented by the software, including Beta(MAF; 1,25) and Beta(MAF; 1,1), using Cauchy combination. Exact mask definitions, weighting schemes and software options are provided in **Supplementary Table 33**.

After harmonization, the benchmark included 562,255 variants (43,326 loss-of-function, 342,916 missense and 176,013 synonymous) on chromosome 19, 1,388 genes and 3,881 gene-level tests per phenotype replicate. Empirical type I error was calculated separately for each test, MAF threshold and censoring level and is reported with exact binomial 95% confidence intervals based on 220,983 to 271,200 contributing tests per cell. Quantile-quantile plots were additionally used to assess departures from the expected null distribution across the full range of association statistics.

Because this comparison was restricted to chromosome 19, the resulting calibration estimates should be interpreted as describing performance under the evaluated high-censoring conditions on that chromosome rather than as a genome-wide characterization of GATE-STAAR.

#### Power simulations

To assess the empirical power of GATE-GENE, we focused on the simulation scenario with a high censoring rate of 90%. In addition to including 100,000 variants to simulate random effects, we selected 10 genes as causal, *APOB*, *BRCA2*, *IGSF9B*, *GPRIN1*, *DDR1*, *DBH*, *CFB*, *HLA-DRB1*, *IL33* and *GPSM3*, spanning 168 to 4,394 qualifying coding variants per gene, to generate phenotypes under the alternative hypothesis and evaluate power. Phenotypes were generated by adding the summed causal-gene effects to the linear predictor, with the censoring rate tuned to give a 10% event prevalence. Given that functionally deleterious variants are more likely to exert large effects on disease risk, we modeled both the proportion and effect sizes of causal variants according to functional annotations. For variants with minor allele count (MAC) > 10, we considered two causal proportion settings: (1) 20% for predicted LoF, 10% for missense, and 2% for synonymous variants; and (2) 30% for LoF, 10% for missense, and 2% for synonymous variants. Since rare deleterious variants tend to be under stronger selective constraint, we assumed that causal proportions among ultra-rare variants (MAC ≤ 10) were three times higher than those among less rare variants (MAC > 10) within each functional category.

To further assess power under realistic genetic architectures, we evaluated two settings for the absolute effect sizes of causal variants: $|0.25 \times \log_{10}(\mathrm{MAF})|$ and $|0.15 \times \log_{10}(\mathrm{MAF})|$ for missense and synonymous variants, respectively. For LoF variants, which are expected to be more impactful, effect sizes were doubled: $|0.5 \times \log_{10}(\mathrm{MAF})|$ and $|0.3 \times \log_{10}(\mathrm{MAF})|$. Two scenarios were considered for the direction of effects among causal variants: (1) all causal variants had effects in the same direction (i.e., all increasing disease risk); and (2) a mixed direction scenario in which 100% of LoF, 80% of missense, and 50% of synonymous causal variants increased disease risk, with the remaining variants having protective effects. In this second scenario, we maintained a consistent effect direction for all LoF variants, reflecting the expectation that true LoF variants are less likely to have opposing effects.

In total, three simulation scenarios were constructed based on different combinations of causal variant proportions and effect size distributions across functional annotations (**Supplementary Table 4**). For each scenario, we conducted 100 simulation replicates, yielding 1,000 *P* values in total (10 causal genes × 100 replicates per gene).

### Supplementary Note 3. Onset-atlas curation and validation

#### Phenotype curation

The UK Biobank phenome-wide onset analysis began with 834 PheCode-defined incident time-to-event phenotypes with more than 200 incident events. An incident event was defined as the first qualifying diagnosis recorded at least 30 days after recruitment among participants without the corresponding phenotype before baseline. The UKB onset atlas used the full WES release and HES inpatient diagnoses for outcome ascertainment. Its smaller HES-restricted analytical population and control eligibility rules were specific to the onset atlas and were not applied to the interim-WES multistate analyses. Control eligibility for the onset atlas was restricted to participants with at least one PheCode-mappable diagnosis in the linked HES inpatient table; participants never hospitalised with a mappable code were not eligible as controls. This restriction removes 133,953 of the 420,542 participants of inferred European ancestry carrying a genotype-quality-control record (31.9%), leaving 286,589 from which the per-phenotype analytical denominators were drawn after exclusion of prevalent cases and application of the landmark.

Phenotypes were curated in two stages. First, we applied a content-based filter to retain clinically meaningful disease-state phenotypes and exclude symptoms and signs, injuries and poisonings, procedure-, treatment-, history- or status-related codes, vague residual categories and exact or near-exact duplicate phenotypes. When strongly overlapping parent and child PheCodes were present, a single representative phenotype was retained for the primary analysis, prioritizing clinical interpretability while considering event count and redundancy.

Second, we evaluated whether first recorded diagnosis during UK Biobank follow-up provided an interpretable approximation of disease onset. Phenotypes were set aside from the core atlas when recorded timing was likely to reflect delayed ascertainment, a downstream complication or acute decompensation, congenital disease timing or another strongly context-dependent clinical event. After curation, 314 phenotypes were retained in the primary broad incident-onset set, 337 clinically meaningful phenotypes were retained for supplementary analyses and 183 phenotypes were excluded. Fourteen of the 314 primary phenotypes were classified as timing-limited, leaving 300 phenotypes in the core onset atlas. Per-phenotype classifications and reasons for exclusion are provided in **Supplementary Table 1**.

#### Curation of germline-focused associations

UK Biobank exome sequencing was performed on blood-derived DNA, and therefore variants arising through clonal haematopoiesis or haematologic neoplasia can appear in exome data alongside inherited variants. To distinguish germline-focused rare-variant associations from signals likely to reflect somatic processes, we curated 42 genes recurrently implicated in clonal haematopoiesis and myeloid or lymphoid neoplasia from established sources[37–45]. This set comprised 34 recurrent somatic driver genes and eight genes with both germline predisposition and recurrent somatic mutation.

Each exome-wide significant gene-phenotype association was evaluated using three complementary sources of information: membership of the curated somatic-driver set; the clinical phenotype involved, particularly myeloid or lymphoid neoplasia, cytopenias, splenomegaly and iron-metabolism phenotypes; and the underlying single-variant architecture. Variant-level review distinguished associations driven by recurrent somatic hotspots, including *JAK2* p.Val617Phe, *SRSF2* p.Pro95 and *ASXL1* c.1934dup, from signals distributed across rare variants consistent with inherited variation. Carrier patterns and case-versus-censored allele-frequency enrichment were additionally considered.

Associations were classified as germline-likely, somatic-likely, somatic-suspect, dual germline-somatic or haematologic-fingerprint signals. Somatic-likely and somatic-suspect associations were excluded from the primary germline-focused atlas. Associations involving dual germline-somatic genes were evaluated at the variant level; for example, established germline founder alleles were retained when the observed carrier pattern was consistent with inherited variation. Signals involving fingerprint genes in the extended MHC were additionally screened for long-range linkage-disequilibrium confounding. The complete gene list, unfiltered association set and per-association classifications are provided in **Supplementary Tables 7 and 30**.

#### Study-wide multiplicity

Study-wide multiplicity was examined as a separate sensitivity analysis from per-phenotype discovery. The primary exome-wide discovery threshold was $P < 2.5 \times 10^{-6}$ for the per-gene Cauchy omnibus assembled from mask-specific SKAT-O $P$ values within each phenotype. For the 300 core onset phenotypes, 5,518,026 gene-phenotype tests were included in the pooled study-wide Benjamini-Hochberg analysis. For the onset atlas the study-wide FDR analysis was applied to the same Cauchy omnibus statistic used for discovery, so the FDR discoveries are a re-thresholding of the discovery statistic rather than independent evidence. Stouffer-combined omnibus P values were used for FDR control in the cross-biobank meta-analysis family only. The expected number of false discoveries under the complete null was calculated separately. Progression, native All of Us and cross-biobank meta-analysis results were treated as independent testing families.

#### Matched incident case-control benchmark

To isolate the contribution of modelling event time beyond the incident disease definition itself, we compared GATE-GENE results with matched incident case-control analyses using SAIGE-GENE+. A panel of 120 UKB-EUR onset phenotypes was selected from the broader onset transition to span censoring proportions, onset-age distributions and PheCode disease categories.

For each phenotype, the case-control analysis used the same participants, incident phenotype definition and variant masks as the corresponding GATE-GENE analysis, but reduced the phenotype to incident case status. Concordance between frameworks was summarized using the Spearman correlation of gene-level $-\log_{10}P$ values, overlap among the 50 highest-ranked genes and sharing of associations reaching $P < 2.5 \times 10^{-6}$ in either framework.

#### Carrier-level attained-age analyses

For each of the 107 germline-focused exome-wide associations, we evaluated whether rare-variant carriers differed from non-carriers in the timing of diagnosis. Carriers were defined using rare LoF variants in the associated gene. Cox proportional-hazards models used attained age as the time scale with delayed entry at cohort enrolment, so that participants entered the risk set at their age at recruitment and were followed until diagnosis or censoring.

Models were adjusted for sex and the first 20 genetic principal components; sex was omitted where inappropriate, and a sensitivity model additionally adjusted for birth year. A discovery-aligned Cox model using follow-up time from cohort entry was also fitted for comparison. Associations with sufficient carrier-event counts for stable estimation were retained, yielding 80 analysable associations. Benjamini-Hochberg correction was applied across these associations.

Because the carrier analyses followed up associations selected in the primary discovery analysis, they were interpreted as characterization of disease timing among already-discovered associations rather than as independent confirmation.

#### GeneBass cross-resource comparison

To assess cross-resource consistency between incident TTE gene-based associations and prevalent case-control exome analyses, we compared GATE-GENE onset results in UKB with phenotype-mapped GeneBass results. Among the 300 core incident-onset phenotypes analyzed by GATE-GENE, 99 were represented in both resources. For each phenotype, comparisons were restricted to genes with valid association *P* values in both GATE-GENE and GeneBass, yielding a median of 17,196 intersecting genes per phenotype (IQR, 17,160-17,224; range, 17,093-17,490) and 1,702,543 matched phenotype-gene results in total.

Global agreement in gene-level association evidence was modest. Across the 99 matched phenotypes, the median Spearman correlation between GATE-GENE and GeneBass -$\log_{10}$P values was 0.069 (IQR, 0.040-0.100), and the median Pearson correlation was 0.092 (Supplementary Fig. 7a ). Exact overlap among top-ranked genes was also limited: the median number of shared genes was 0 (IQR, 0-1) among the top 20, 1 (IQR, 0-2) among the top 50 and 8 (IQR, 5-13) among the top 200. These patterns are consistent with differences between incident-onset and prevalent case definitions, TTE and case-control modeling frameworks, and the variant masks used in the two resources.

Despite modest global rank concordance, genes prioritized by the incident TTE analysis showed systematic enrichment for GeneBass evidence. For each phenotype, we compared GeneBass association evidence among the 200 genes most strongly prioritized by GATE-GENE with that among the remaining genes in the corresponding phenotype-specific intersection. GATE-GENE top-200 genes had significantly stronger GeneBass evidence in 74 of 99 phenotypes (74.7%) after Benjamini-Hochberg correction across phenotypes (FDR < 0.05; Supplementary Fig. 7b). The median difference in GeneBass -$\log_{10}P$ values between GATE-GENE top-200 genes and the remaining intersecting genes was 0.090. Thus, GeneBass support was detectable as a distributional enrichment even when exact gene rankings differed between resources.

We next evaluated the 107 exome-wide significant GATE-GENE associations classified as germline-likely. Fifty-one associations occurred in 27 phenotypes represented in both analyses. Five of these involved genes without an available GeneBass result for the corresponding phenotype, leaving 46 associations across 26 phenotypes directly comparable between GATE-GENE and GeneBass. Associations without a matched phenotype or gene-level result were treated as unavailable for comparison rather than as unsupported.

Among the 46 jointly comparable associations, GATE-GENE and GeneBass association strengths showed moderate concordance (Spearman $\rho$ = 0.342; Pearson $r$ = 0.345). Thirty associations (65.2%) had GeneBass $P < 0.05$, 28 (60.9%) had $P < 0.01$, 24 (52.2%) had $P < 10^{-3}$

and 18 (39.1%) reached $P < 2.5 \times 10^{-6}$. Thirty associations also remained significant after Benjamini-Hochberg correction across the 46 jointly comparable associations (FDR < 0.05).

Together, these analyses show that incident TTE and prevalent case-control gene rankings are only modestly correlated across the exome, but that concordance becomes more apparent among genes prioritized by GATE-GENE and among core exome-wide significant associations. Because both resources are derived from UKB exome data, this comparison should be interpreted as cross-analysis support rather than independent cohort replication.

### Supplementary Note 4. Functional enrichment and rare-common convergence analyses

#### Functional enrichment of onset-associated genes

We evaluated functional enrichment among the 107 germline-focused gene-phenotype associations, comprising 89 unique associated genes. The primary union list contained all 89 genes. We additionally constructed disease-domain-specific lists containing genes associated with at least one phenotype in each PheCode category, a pleiotropic list containing genes associated with at least three phenotypes and a union-without-neoplasm list excluding genes represented in the neoplasm category. A gene associated with phenotypes from more than one disease domain could contribute to multiple domain-specific lists. The background universe comprised 18,399 genes with a valid Cauchy-combined gene-level association *P* value in the 300 core onset phenotypes.

Hallmark, Reactome and Gene Ontology Biological Process gene sets were obtained from MSigDB using msigdbr. After intersection with the tested-gene universe, Hallmark and Reactome gene sets containing 10-500 genes and Gene Ontology sets containing 20-800 genes were retained. GTEx v8 tissue-expression sets were constructed from the 500 genes with the highest median expression in each tissue.

For each associated-gene list and gene set, over-representation was tested using a one-sided Fisher's exact test with the tested-gene universe as the background. Odds ratios, overlap counts and contributing genes were retained. The primary analyses comprised the full union, union-without-neoplasm and pleiotropic lists and the neoplasm, circulatory-system and genitourinary-system domain lists. Benjamini-Hochberg correction was applied jointly across pathway tests for these primary lists; secondary within-family FDR values were additionally calculated for individual gene-list-collection combinations.

Because rare-variant discovery probability depends on gene characteristics, enrichment robustness was evaluated using covariate-matched gene-set permutations. For each associated gene, control genes were matched on coding length, number of qualifying variants, cumulative minor allele count, median number of masks tested across phenotypes and the proportion of phenotypes with a valid SKAT-O result. We generated 10,000 matched control sets with the same number of genes as each observed list and calculated empirical P values as the proportion of matched sets with pathway overlap at least as large as observed. Empirical P values were adjusted across pathways and primary gene lists using the Benjamini-Hochberg procedure.

We additionally performed leave-one-gene-out analyses for the neoplasm, circulatory-system, genitourinary-system, pleiotropic and union-without-neoplasm lists. A pathway was considered robust to leave-one-gene-out analysis when it remained significant at within-family FDR <0.05 in at least 80% of iterations.

As a threshold-free sensitivity analysis, Cauchy-combined gene-level $P$ values from all 300 core phenotypes were converted to capped $-\log_{10}P$ scores, rank-normalized within phenotype and residualized for coding length, qualifying-variant count, cumulative minor allele count and gene-level testability. For each pathway, we tested whether member genes had a positive mean residual association score. Pathway-level $P$ values were then combined across phenotypes using the aggregated Cauchy association test with equal phenotype weights, and global FDR was calculated across all Hallmark, Reactome and Gene Ontology pathways.

#### Common-variant locus burden per phenotype

We performed genome-wide common-variant survival association analysis for each of the 300 core incident-onset phenotypes using GATE implemented in SAIGE. Genome-wide summary statistics were clumped with PLINK[50] using --clump-p1 1e-6, --clump-p2 1e-6, --clump-r2 0.1 and --clump-kb 500. Each clumped common-variant locus was represented by its lead variant. Two locus counts were retained per phenotype: the number of clumped loci reaching $P < 1 \times 10^{-6}$ and the number reaching genome-wide significance, defined as $P < 5 \times 10^{-8}$. The genome-wide significant locus count was used as the primary common-variant association-burden measure. The $P < 1 \times 10^{-6}$ locus count was used as a sensitivity measure. Phenotypes for which no lead variant reached $P < 1 \times 10^{-6}$ produced no PLINK clump output and were assigned a clumped-locus count of zero rather than being dropped.

#### Phenotype-level convergence and permutation nulls

The primary phenotype-level convergence test was the Spearman rank correlation between genome-wide significant common-locus count and germline-focused rare-significant gene count across the 300 core phenotypes. Significance was assessed both asymptotically and by permutation. In addition to unrestricted label permutation, we constructed four stratified permutation nulls, each with 10,000 permutations in which rare-gene counts were shuffled only within strata: event-count quintile; event-count quintile crossed with analysis batch; event-count quintile crossed with censoring-rate tertile; and PheCode disease domain. Strata containing a single phenotype were left unpermuted. These stratified permutation analyses tested whether common-variant association burden predicted rare-gene discovery yield beyond what was observed among phenotypes with similar discovery opportunity. Empirical P values were computed as (b + 1)/(B + 1), with B = 10,000 permutations, giving a minimum attainable value of $9.999 \times 10^{-5}$, reported as $P_{perm} < 1 \times 10^{-4}$ when no permutation exceeded the observed statistic.

Stability of the phenotype-level correlation was evaluated by leave-one-phenotype-out refitting across all 300 phenotypes, leave-one-disease-domain-out refitting across 13 disease domains, exclusion of the five phenotypes with the largest number of common-variant loci, restriction to the 67 rare-positive phenotypes and exclusion of genes in the extended MHC region.

#### Count models for rare-gene discovery yield

Because germline-focused rare-gene discovery counts were non-negative integers and 233 of 300 phenotypes had zero germline-focused rare-gene discoveries, the relationship between common-variant and rare-variant association burden was modelled as a count process rather than by linear regression on a transformed outcome. A negative binomial model with log link was fitted for rare-gene count as a function of log(1 + genome-wide significant common-locus count), adjusted for log(1 + event count). The negative binomial model was preferred to a Poisson model by likelihood-ratio test. Sensitivity models additionally adjusted for censoring rate, log(1 + sample size), case fraction and log(number of genes tested). Rate ratios were reported per one-unit increase in log(1 + genome-wide significant common-locus count).

Because of the large number of zero counts, the count process was additionally decomposed using a hurdle model. The binary component used logistic regression to model whether a phenotype yielded any germline-focused rare-significant gene among all 300 phenotypes. The positive-count component used a zero-truncated negative binomial model for the number of rare-significant genes among the 67 rare-positive phenotypes. The zero-truncated component was fitted by direct maximum likelihood, with standard errors obtained from the numerical Hessian, because an ordinary negative binomial model fitted only to positive phenotypes is not a hurdle component and can underestimate the association.

Dependence among related endpoints was addressed in three ways. First, block permutation within the PheCode disease domain was included among the phenotype-level permutation nulls. Second, we repeatedly sampled one phenotype per domain over 2,000 draws and refitted the phenotype-level association. Third, domain-stratified negative binomial models were fitted and combined using DerSimonian-Laird random-effects meta-analysis across the eight disease domains with at least ten phenotypes and at least three rare-positive phenotypes. Heterogeneity was summarized using Cochran's Q and $I^2$.

#### Positional overlap and matched permutation null

A germline-focused rare-significant gene was counted as positionally overlapping a common-variant association when its gene body fell within $\pm$250 kb of a genome-wide significant clumped common-variant locus for the same phenotype. The extended MHC region, defined as chr6:25 to 34 Mb, was excluded from the primary analysis because long-range linkage disequilibrium and high gene density can place many genes under a single common-variant association signal. MHC-retained analyses and alternative windows of $\pm$100 kb and $\pm$500 kb were performed as sensitivity analyses. To count independent regions rather than individual lead variants, clumped lead variants within 500 kb of one another on the same chromosome were merged.

Rare-significant genes were systematically larger and more variant-rich than the average tested gene, and larger genes are more likely to intersect a common-lead window by chance. A hypergeometric test against the full tested-gene background was therefore considered anti-conservative. The primary positional-overlap null was a covariate-matched permutation that preserved gene-level discoverability. Within each phenotype, rare-gene labels were permuted among that phenotype's tested genes while holding fixed both the number of rare-significant genes and the observed phenotype-specific common-variant loci. Permuted genes were sampled uniformly from the same joint covariate stratum as the gene being replaced. Strata were defined by chromosome crossed with genome-wide tertiles of coding length, number of qualifying variants and cumulative minor allele count. This approach used discrete binning rather than caliper or nearest-neighbour matching. When a chromosome-specific joint stratum contained fewer than five candidate genes, sampling fell back to the corresponding covariate-only stratum. If no covariate-only stratum was available, sampling fell back to any tested gene for that phenotype. Genes lacking covariate annotation were assigned to their own stratum, ensuring that they were matched to one another rather than dropped. Fallback rates were reported. The matched null was evaluated over 10,000 permutations.

#### Conditional analysis of rare-variant signals

For each germline-focused gene-phenotype pair, rare-variant burden and omnibus models were refitted after conditioning on phenotype-specific common lead variants. Independent common signals were selected using LD clumping and carried into conditional GATE step 2. For each pair, the burden mask reported in the unconditional analysis was the mask selected from the unconditional analysis. The identical mask, defined by variant annotation group and maximum minor allele frequency, was evaluated in the conditional analysis so that unconditional and conditional estimates were directly comparable. The omnibus statistic was the Cauchy combination of SKAT-O *P*-value across masks, and exome-wide significance was defined as $P < 2.5 \times 10^{-6}$ for this statistic.

Conditional fits were screened for degeneracy before summary statistics were computed. A fit was classified as unstable if the conditional estimate or standard error was missing, if the conditional standard error was zero or smaller than one tenth of the unconditional standard error, or if the absolute conditional effect estimate exceeded 5. These filters were applied because degenerate conditional fits can appear as spuriously strengthened associations and inflate apparent retention. Effect stability, effect ratios and retention rates were summarized only among fits that converged and passed these stability criteria.

#### Pathway-level rare-common convergence

Common-variant gene-based and gene-set statistics were computed using MAGMA. Variants were annotated to genes using NCBI37.3 coordinates with a 35-kb upstream and 10-kb downstream window. Gene-based *P* values were computed using a European linkage disequilibrium reference with fixed N = 361,144. Competitive gene-set analyses were performed using MSigDB C5 Gene Ontology Biological Process terms, version 2025.1, with Entrez identifiers. Significant common-variant pathway-phenotype associations were defined using a within-phenotype false discovery rate of 5%.

Rare-variant pathway enrichment was assessed using over-representation analysis of germline-focused rare-variant gene lists against the same Gene Ontology Biological Process collection. Ten rare-variant gene lists were prespecified: the full rare-gene union, a pleiotropic gene list containing genes associated with three or more phenotypes, a phenotype-specific gene list containing genes associated with exactly one phenotype and seven disease-domain-specific lists. For each list, overlap between rare-enriched terms and common-variant enriched terms was tested against a hypergeometric null over the Gene Ontology Biological Process terms tested in the rare-variant analysis. Benjamini-Hochberg correction was applied across all ten prespecified lists. The four lists yielding no significant rare-side terms contributed P = 1 to the correction family. A pooled summary across lists was constructed post hoc for descriptive purposes and was excluded from the corrected inference.

Shared pathway terms were collapsed into non-redundant clusters using the gene-set similarity clustering procedure from the rare-variant enrichment pipeline. A cluster was considered supported if at least one term retained at least two genes contributing to the rare-variant enrichment. Concentration of rare-common pathway convergence on a small gene core was quantified by counting gene-term memberships and by recounting supported clusters after removing *TTN* alone and after removing *TTN*, *MYH7* and *MYBPC3*.

#### Heritability quality control

LD score regression (LDSC) was run for all 300 core phenotypes using summary statistics munged against HapMap3 variants and precomputed European LD scores[61]. A phenotype was considered to pass heritability quality control if the heritability estimate and standard error were positive, the heritability z score exceeded 1.5 and mean $\chi^2$ exceeded 1.02. 63 of 300 phenotypes passed these criteria, and 69 had mean $\chi^2$ below 1, with median mean $\chi^2$ of 1.01 and median intercept of 1.00; all 300 intercepts lay between 0.95 and 1.05. Clumped common-variant locus counts were used as the primary convergence measure.

### Supplementary Note 5. Rare-common convergence diagnostics

#### Conditional-analysis diagnostics

All 107 germline-focused gene-phenotype pairs were evaluated in conditional rare-variant models. Seventeen pairs produced degenerate conditional fits: ten had a conditional standard error of exactly zero, five had a conditional standard error more than tenfold smaller than in the unconditional fit, one had an implausibly large conditional effect estimate (*ACVRL1* in disease of capillaries, $\beta$ = 16.5), and one returned no conditional estimate (*SLC22A3* in coronary atherosclerosis). These patterns were treated as numerical failures rather than biological findings. Because collapsed standard errors can make associations appear artificially strengthened after conditioning, retention and effect-stability summaries were reported only among fits that converged and passed these diagnostic filters. When all 107 pairs were included, 94 remained exome-wide significant after conditioning (87.9%); among the 90 pairs with stable fits, 81 remained exome-wide significant (90.0%). The latter value is reported in the main text. All excluded pairs and their failure modes are listed in **Supplementary Table 17**.

Two of the 14 positional rare-common overlaps had degenerate conditional fits, both involving *SLC22A3* in angina pectoris and coronary atherosclerosis. Conditional-effect summaries for positionally overlapping loci were therefore calculated over 12 pairs rather than all 14 overlaps. Among the 90 stable conditional fits, conditioning had little effect on the typical rare-variant estimate.

The median change in $-\log10P$ was −0.01 for the omnibus statistic and −0.05 for the selected burden mask. The median conditional-to-unconditional burden-effect ratio was 1.00 (IQR, 0.98 to 1.02). Two pairs changed effect direction and were excluded from the ratio summary; among the remaining 88 pairs, four had ratios below 0.8 and none had ratios below 0.5.

#### Interpretation of below-threshold conditional associations

Nine stable pairs fell below the exome-wide significance threshold after conditioning. An initial interpretation was that these losses reflected reduced precision after conditioning on correlated common variants. The data did not support this explanation. Among these nine pairs, the median ratio of conditional to unconditional standard error was 0.99, and the standard error increased in only two pairs: *HLA-C* in ankylosing spondylitis (ratio, 1.94) and *AP2A1* in emphysema (ratio, 1.02). Effect direction was preserved in eight of the nine pairs, with *HLA-C* in ankylosing spondylitis being the exception.

Instead, below-threshold status was driven primarily by weaker aggregate evidence across masks and tests. The omnibus Cauchy statistic weakened more than the selected burden mask in eight of the nine pairs, with a median reduction in $-\log10P$ of 2.81 for the omnibus statistic compared with 0.45 for the selected burden statistic (Supplementary Fig. 11f). Because exome-wide significance was declared on the omnibus statistic, a pair could fall below threshold while the selected burden-effect estimate remained largely unchanged. This pattern was most pronounced for *HOXB13* in prostate cancer (omnibus reduction, 8.9; burden reduction, 3.6), *HLA-C* in ankylosing spondylitis (8.7 versus 1.8) and *MUC22* in ankylosing spondylitis (6.1, while the selected burden statistic strengthened). The latter two pairs lie in the extended MHC, where long-range linkage disequilibrium can contribute signals across multiple masks and tests.

#### Phenotype dependence

The 300 core onset phenotypes are not statistically independent, because related endpoints can share cases, risk factors and genetic architecture. We used three analyses to evaluate whether phenotype dependence explained the observed correlation between common-variant locus burden and germline-focused rare-gene discovery yield. First, block permutation within PheCode disease domain preserved significance of the phenotype-level correlation ($P_{perm} = 1 \times 10^{-4}$). Second, repeated subsampling of one random phenotype per disease domain over 2,000 draws gave a median Spearman correlation of 0.31, matching the full-data estimate. This subsampling analysis had limited power because each draw included only 13 phenotypes, and the distribution was correspondingly wide (IQR, 0.07 to 0.50), with 22% of draws at or below zero. We therefore interpret it as a robustness analysis of the median effect rather than as an independent significance test.

Third, count-based models that accounted for disease-domain structure supported the same conclusion. Adding disease-domain fixed effects to the negative binomial model gave a rate ratio of 1.85 ($P = 4.6 \times 10^{-5}$), similar to the estimate from the primary event-count-adjusted model. Fitting the model separately within disease domains and pooling estimates by random-effects meta-analysis gave a rate ratio of 2.08 (95% CI, 1.14 to 3.80; $P = 0.017$), again close to the primary estimate of 2.03.

Heterogeneity across disease domains was moderate ($I^2 = 50\%$; Cochran's $Q = 18.7$ on 7 degrees of freedom). Domain-specific rate ratios ranged from below 1 in neoplasms and genitourinary disease to above 4 in sense-organ and digestive phenotypes, with wide confidence intervals in domains containing few rare-positive phenotypes. Leave-one-domain-out refitting showed that no single domain drove the overall phenotype-level correlation: Spearman $\rho$ ranged from 0.28 to 0.36 across the 13 refits, with all $P < 3.6 \times 10^{-6}$.

#### Extended MHC sensitivity

The extended MHC region, defined as chr6:25 to 34 Mb, inflated positional rare-common overlap statistics. When the MHC was retained, we observed 27 positional overlaps across 11 phenotypes. Thirteen of these overlaps lay within the MHC and arose from only two phenotypes, hypothyroidism and ankylosing spondylitis. After excluding the extended MHC, 14 positional overlaps remained across 10 phenotypes. Because high gene density and long-range linkage disequilibrium can place multiple genes under a single common-variant association signal in the MHC, MHC-derived overlaps were not treated as independent observations and the extended MHC was excluded from the primary positional-overlap analysis.

The phenotype-level rare-common correlation was not sensitive to MHC handling. Removing MHC genes from rare-gene counts gave $\rho = 0.32$ ($P = 1.6 \times 10^{-8}$), and removing the two MHC-driven phenotypes entirely gave $\rho = 0.31$ ($P = 3.1 \times 10^{-8}$), compared with $\rho = 0.335$ in the full data.

#### Concentration of the pathway signal

Pathway-level convergence was detectable but narrow. Of the ten prespecified rare-variant gene lists, only the circulatory disease-domain list showed rare-common overlap exceeding chance. All ten rare-enriched circulatory terms were also enriched in the common-variant analysis, corresponding to a 7.8-fold enrichment over expectation (hypergeometric $P = 3.8 \times 10^{-9}$; $q = 3.8 \times 10^{-8}$ after Benjamini-Hochberg correction across the ten prespecified lists). By contrast, the full rare-gene union did not show excess overlap with common-variant enriched terms (three observed terms versus 5.8 expected; $P = 0.94$). Thus, a union-level analysis would not have supported a broad pathway-convergence claim.

Within the circulatory result, the ten shared terms collapsed into six redundancy clusters and were dominated by a small sarcomeric gene core. *TTN*, *MYH7* and *MYBPC3* appeared in all ten shared terms and accounted for 30 of 42 rare gene-term memberships (71%). Removing *TTN* alone left all six clusters supported, whereas removing all three genes left two supported clusters. Three of the eight genes contributing to the pathway overlap, *TTN*, *CACNA1D* and *SLC22A3*, also appeared among positional rare-common overlaps. Thus, the pathway- and locus-level convergence results are partially shared, but not identical. The pathway result should therefore be interpreted as convergence concentrated in a small sarcomeric gene core rather than as a broadly distributed cardiovascular program.

##### Deflation of survival summary statistics

LD score regression indicated systematic deflation of the common-variant survival statistics. Of the 300 core phenotypes, 295 had mean $\chi^2$ below 1, with a median mean $\chi^2$ of 0.85 and a median LDSC intercept of 0.86. Only three phenotypes passed heritability quality control. This pattern is consistent with the use of time-to-event statistics for incident endpoints with modest event counts and indicates that LDSC-derived heritability and mean $\chi^2$ are not reliable measures of common-variant signal in this setting.

All rare-common convergence analyses therefore used clumped common-variant locus counts rather than LDSC-derived quantities. This choice is conservative for the present question: deflation reduces the number of common loci passing fixed significance thresholds and would be expected to attenuate, rather than create, the observed rare-common convergence.

#### Supplementary Note 6. UK Biobank multi-state trajectory analyses

##### Cause-specific direct downstream transition and clinical-route composition

The cause-specific direct T0→T2 transition was rebuilt post hoc from the existing baseline-entry phenotype files, so rows, covariates and the time scale are unchanged and only the exit time, the event indicator and the prevalent-T1 exclusion differ. Participants carrying the T1 PheCode at baseline were excluded as not at risk. When T1 and T2 were first recorded on the same date their ordering was treated as indeterminate and the observation contributed an event to neither transition; no wider ordering window was applied, because same-day recording reflects the resolution of the record rather than a clinical interval. Arms retaining fewer than 50 events were not analysed, leaving 91 of the 95 trajectories. The route decomposition underlying these analyses is as follows: across trajectories a median of 26.6% of downstream events occurred in participants with the upstream code recorded at any time, comprising 4.0% with prevalent T1 at baseline, 10.6% reached through incident T1 before the downstream event–the genuinely T2-event overlap–and 9.3% in whom the upstream code appeared only after the downstream event. Only the middle component is removed by the direct transition. Direct T0→T2 is defined by censoring at incident T1, not by excluding all participants who ever develop T1. A T2 event following T1 is ineligible as a direct event; the separate T1-free sensitivity excludes participants with any observed T1. The principal quantity in the main Results is the T2-event overlap fraction: marginal T2 events following incident T1 divided by eligible marginal T2 events. This descriptive measure must be distinguished from the wider clinical-route composition and from any causal pathway decomposition.

#### Trajectory definition and inclusion criteria

A disease trajectory was defined as an ordered pair of clinical states, consisting of an upstream state T1 and a downstream state T2. Each state was defined using phecode groupings of ICD-9 and ICD-10 codes from linked UK Biobank hospital inpatient records, death registry records and primary care data. For every trajectory, we generated three transitions: T0→T1, representing onset of the upstream state from baseline; T0→T2, representing onset of the downstream state from baseline; and T1→T2, representing progression from the upstream state to the downstream state. These trajectory analyses used the interim approximately 200,000-participant UKB WES release and did not require a recorded HES encounter for inclusion as an otherwise eligible event-free control. This is distinct from the HES-restricted phenotype and control selection used in the full-WES onset atlas (Supplementary Note 3).

Candidate trajectories were curated into clinical tiers before interpreting genetic association results. Tier 1 comprised clinically established disease sequences, whereas lower tiers captured broader or less specific relationships used for sensitivity analyses. Of 199 candidate trajectories in the European-ancestry exome-sequencing analysis set, 95 entered the primary analysis. Trajectories were excluded when at least one of the three component transitions was unavailable (n = 72), when the progression transition accrued fewer than 100 T1→T2 events (n = 24), or when the progression event count could not be determined (n = 8). The full inclusion funnel, including per-transition sample size, event count, censoring, follow-up and attained age at diagnosis for every candidate trajectory, is provided in **Supplementary Table 2**. The 95 analysed trajectories comprised 165 distinct transitions, because a single baseline-entry transition could serve as the T0→T1 arm of one trajectory and the T0→T2 arm of another.

#### Time scale, risk sets and covariates

GATE does not directly accommodate left truncation. We therefore analysed each transition using time since transition-specific risk-set entry, with follow-up beginning when individuals became eligible for the corresponding analysis. This approach allowed transition-specific entry dates without introducing an artificial event-free interval.

For baseline-entry transitions (T0→T1 and marginal T0→T2), the initial reference date was UK Biobank recruitment. Individuals with a qualifying diagnosis recorded before recruitment were excluded. An 30-day onset landmark was applied, with the analysis time origin shifted accordingly. Individuals diagnosed or censored within this interval were excluded, ensuring that all retained participants were at risk from the analysis time origin without requiring delayed entry. For progression (T1→T2), the risk set comprised individuals with incident T1 who remained free of T2 at the T1 diagnosis date; those with T2 before or concurrent with T1 were excluded. Follow-up began at incident T1 and measured time since the upstream diagnosis, without an additional washout period. Attained age at an event or censoring could be recovered by adding follow-up time to age at risk-set entry.

Diagnoses were ascertained exclusively from linked UK Biobank hospital inpatient records, with ICD-9 and ICD-10 codes mapped to PheCodes (v1.2b1). The available inpatient history was used to identify prevalent disease; no fixed retrospective lookback window was imposed. Follow-up was censored at death or the nation-specific administrative cutoff, whichever occurred first: 31 March 2023 for England, 31 August 2022 for Scotland and 31 May 2022 for Wales. Diagnoses recorded after these cutoffs were not counted as events. Death-registry information was used for censoring, whereas cause-of-death diagnoses, primary care, cancer registry and self-reported conditions were not used for phenotype ascertainment.

All transition analyses were adjusted for age at risk-set entry, sex and the first ten genetic principal components. Sex was omitted for sex-restricted phenotypes because it was constant within those analyses. Progression analyses were additionally adjusted for birth year. This adjustment was used because baseline-entry analyses had recruitment dates concentrated between 2006 and 2010, making age at recruitment nearly collinear with birth year, whereas progression entry dates spanned a broader calendar-time interval and birth year captured additional cohort effects.

#### Gene-based rare-variant association testing

Gene-based rare-variant association tests were performed using GATE-GENE/SAIGE-GENE+ on UK Biobank exome-sequencing data, restricted to individuals of European genetic ancestry. Null models were fitted separately for each transition analysis using the survival trait type, with event indicator and time since risk-set entry as the outcome. A sparse genetic relatedness matrix with relatedness cutoff 0.05 was used to account for relatedness. Association testing was performed with leave-one-chromosome-out disabled and with a minimum minor allele count of 0.5 for single-variant inclusion.

Each gene was tested under three functional masks: predicted loss-of-function, loss-of-function plus missense, and loss-of-function plus missense plus synonymous. Each mask was evaluated at three maximum minor allele frequency thresholds: $1 \times 10^{-4}$, $1 \times 10^{-3}$ and $1 \times 10^{-2}$. This yielded up to nine mask-by-frequency cells per gene per transition analysis. Within each cell, GATE-GENE/SAIGE-GENE+ returned burden, variance-component and Cauchy-combined test statistics.

#### Harmonised gene-level omnibus statistic

For each gene and transition, cell-level *P* values were combined into a gene-level omnibus statistic using the Cauchy combination test. We recomputed the omnibus statistic rather than relying directly on the default SAIGE-GENE+ output to ensure comparability across transition classes. The primary harmonized transition-concordance omnibus is a separate six-cell analysis (three annotations by MAF ceilings 0.1% and 1%); it must not be conflated with the nine-cell discovery omnibus.

Two harmonisation steps were required. First, the $1 \times 10^{-4}$ frequency bin was frequently unavailable in small progression risk sets because, when the risk-set size is below 5,000, a single allele can exceed a frequency of $1 \times 10^{-4}$. This bin was present in baseline-entry analyses but absent from many progression analyses. Retaining it would cause the omnibus statistic to combine a different number of mask-frequency cells across transition classes for reasons unrelated to biology. We therefore recomputed the primary harmonised omnibus statistic over the $1 \times 10^{-3}$ and $1 \times 10^{-2}$ frequency ceilings for all analyses. When no mask-frequency cells were dropped, the harmonised combination reproduced the native SAIGE-GENE+ Cauchy combination.

Second, mask-frequency cells returning $P = 1$ were excluded before Cauchy combination. These *P* values represent untestable cells rather than evidence of no association. Because the Cauchy transform is unstable near $P = 1$, retaining such cells could dominate the combined statistic and differentially affect progression analyses. The Cauchy combination was implemented with an explicit small-P branch below $P = 1 \times 10^{-15}$ to avoid numerical flooring of the combined $P$ value.

Exome-wide significance was defined as $P < 2.5 \times 10^{-6}$, corresponding to 0.05 divided by approximately 20,000 tested genes. A suggestive threshold of $P < 1 \times 10^{-4}$ was used for association-sharing and pathway-enrichment analyses.

#### Interpretation of burden-effect estimates

The BETA estimate returned by SAIGE-GENE+ for a gene-based burden test is the coefficient of a Beta(1,25)-weighted burden score. It is therefore not a per-allele log hazard ratio. Empirical calibration comparing single-variant and gene-burden standard errors indicated that the burden-score scale differs substantially from a per-allele covariate scale. Accordingly, exp(BETA) values are reported only as burden-score effects. They are comparable across transitions when the same mask is evaluated, but they should not be interpreted as per-allele or per-carrier hazard ratios. Exponentiating the weighted burden coefficient yields the hazard ratio per unit increase in the prespecified weighted score, not a per-allele hazard ratio; it is sensitive to the exact variant mask and allele weights.

For candidate progression-specific associations, we fixed the mask that carried the progression association and evaluated that same mask in the corresponding T0→T1 and T0→T2 analyses of the same trajectory. We did not compare the best-scoring mask in each transition, because the best-scoring cell of a gene can differ between transitions and would not provide a like-for-like effect comparison.

#### Gene-level association concordance across transition-specific analyses

Similarity between two transition analyses was quantified using Spearman correlation of gene-level $-\log_{10}P$ values over genes tested in both analyses, using the harmonised omnibus statistic. For each trajectory, we computed the correlation between T1→T2 and T0→T1 and the correlation between T1→T2 and T0→T2. These paired correlations were compared across trajectories using Wilcoxon signed-rank tests. We further compared progression (B; T1→T2) with the direct downstream transition (D; T0→T2 censored at incident T1). The paired B–C versus B–D analysis was restricted to 91 UKB trajectories with at least 50 direct-transition events; the B progression analysis was shared between contrasts.

We also constructed an empirical null distribution from all pairs of transitions. Because concordance depends on statistical power, transition pairs were stratified by the minimum event count of the two analyses. Because concordance also depends on transition-pair composition, transition pairs were additionally stratified by transition-pair composition, defined as the number of progression analyses in each pair. This step was necessary because progression analyses draw their risk sets from individuals who reached T1, whereas baseline-entry analyses draw their risk sets from the full cohort. In addition, two baseline-entry analyses are expected to correlate more strongly with each other than a pair containing a progression analysis.

Null-adjusted concordance was defined as the observed Spearman correlation minus the median correlation of disjoint transition pairs matched on both event-count stratum and transition composition. Disjoint transition pairs were defined as pairs sharing no disease state. Strata with fewer than 30 matched-null pairs were not reported. The same transition-pair-composition matching was used for the marginal (B-C) and direct (B-D) progression comparisons, with null-adjusted correlations obtained separately for each observed pair.

Several controls were used to assess the concordance metric. Positive controls consisted of onset-pair comparisons expected to share genetic architecture. Effect-estimate concordance was evaluated using the correlation of BETA estimates and the fraction of genes with concordant BETA direction, restricted to genes matched on gene and mask with MAC ≥20. These statistics were compared with the same quantities computed against unrelated onset analyses randomly drawn from the trajectory set. Robustness to statistical power was assessed by repeating the concordance analyses after restricting transition pairs to minimum event-count thresholds of 100, 200 and 500. Robustness to the choice of *P*-value-based ranking was assessed using fixed-rank gene overlap instead of fixed *P*-value thresholds.

#### Calibration of transition analyses

Tail calibration was summarized using $\lambda_{0.01}$, defined as the genomic-control factor at the 1st percentile of the gene-level *P*-value distribution. This statistic was computed as the observed chi-square statistic at that quantile divided by the corresponding expected chi-square statistic under the null. λ0.01 was used as the primary calibration metric because it remains defined when a transition has no genes below a stringent fixed *P*-value threshold.

Fixed-threshold observed-to-expected gene counts were retained as secondary diagnostics but were not used as the primary calibration display. These ratios are unstable in sparse progression analyses because the expected count can be very small. For example, a transition testing approximately 18,000 genes expects only 0.045 genes below $P < 2.5 \times 10^{-6}$ under the null, making the observed-to-expected ratio either zero or large for most analyses.

Because λ0.01 tracked censoring across transition classes, comparisons of $\lambda_{0.01}$ between transition classes were reported with adjustment for censoring. The three progression analyses carrying exome-wide associations were additionally checked against the progression distribution of λ0.01, censoring and carrier-count availability.

#### Per-mask sensitivity and carrier-count availability

To evaluate whether sparse progression discovery reflected weaker per-gene sensitivity or fewer testable rare-variant masks, achieved standard errors were summarized by minor allele count stratum and transition class. Because MAC is an allele count, a given MAC corresponds to a larger carrier fraction in progression analyses than in baseline-entry analyses, since progression risk sets are smaller. Comparisons at matched MAC therefore compare sensitivity at equal carrier count rather than at equal allele frequency.

For each mask-frequency cell, per-mask information was summarized as events × $p_{\text{carrier}}$ × (1 − $\lambda_{0.01}$), where $\lambda_{0.01}$ denotes the carrier fraction in the relevant transition-specific risk set. We also summarized the fraction of masks reaching prespecified MAC thresholds by transition class, including MAC ≥20 and MAC ≥50, to quantify the number of testable gene-mask cells in progression versus baseline-entry analyses.

#### Longer clinical chains

The 95 analysed trajectories were assembled into a directed graph whose nodes represented disease states and whose edges represented progression transitions. Simple directed paths of two to five edges were enumerated, yielding 947 longer clinical chains. For each pair of consecutive edges within a chain, gene-level concordance was computed using the same harmonised omnibus P-value vectors described above. In total, this analysis included 155 adjacent edge pairs drawn from 70 distinct edges sharing 16 disease states. Adjacent-edge concordance was compared with disjoint transition pairs as a non-adjacent baseline.

#### Gene-set enrichment of transition-specific associations

Genes reaching $P < 1 \times 10^{-4}$ in exactly one transition class were tested for pathway enrichment against MSigDB gene-set collections. Gene-set membership tables were deduplicated to one row per gene set and gene before enrichment testing, because the source tables can contain one row per gene set, gene and source identifier. Without deduplication, set sizes and overlap counts would be inflated. Enrichment analyses were seeded for reproducibility.

### Supplementary Note 7. Illness-death simulation and partial-sharing analyses

#### Detectability of shared effects

Because progression analyses carry far less genetic information than baseline-entry analyses, we bounded rather than estimated the genome-wide variance of rare-variant burden effects. A method-of-moments estimator $\tau^2$ = Var($\hat{\beta}$) − mean(SE²) was applied within each transition class; under the null its standard error is approximately $\sqrt{(2.mean(S^4)/G}$ for $G$ genes, which agreed with simulation to within 2%. The smallest detectable per-gene effect standard deviation scaled with events −0.80 across transitions, giving detection floors of 0.017 for progression, 0.005 for marginal downstream and 0.0036 for onset. A transition-level genetic correlation is not identifiable from these data because its denominator requires $\tau^2$ in both arms; we therefore report the near-zero onset–progression concordance as limited detectable sharing of association signals rather than as an estimate of genetic correlation.

We simulated individual-level illness-death processes (N = 150,000; 3,000 genes with sparse carrier sets, carrier frequency $2 \times 10^{-4}$ to $1.5 \times 10^{-2}$) under three architectures: (A) susceptibility-only, in which genes affect only the T0→T1 transition; (B) shared correlated effects, in which onset and progression effects are drawn with correlation $\rho$ = 0.7; and (C) stage-specific, in which distinct genes carry downstream-disease effects expressed on the T0→T2, T1→T2 and post-T2 transitions. Each scenario was run with and without EHR under-ascertainment of T1 and with index-event selection into the progression risk set, and simulated gene-level statistics were pushed through the same concordance pipeline as the real data, including the composition-matched disjoint null (**Supplementary Table 24**).

The positive progression-marginal concordance contrast arose under several simulated architectures and therefore was not uniquely diagnostic of shared causal effects. In the originally specified shared-effect architecture, onset-progression concordance was greater than observed, whereas the susceptibility-only architecture more closely matched the near-zero onset-progression null-adjusted concordance. These comparisons are conditional on the modelled effects, carrier frequencies, recording assumptions and progression event counts; they do not exclude substantial underlying causal-gene sharing in other rare-variant architectures or imply a causal decomposition of observed clinical transitions. A separate progression–direct simulation described below evaluates this detectability issue directly.

The naive ordering did not discriminate: all three scenarios produced a positive median null-adjusted concordance of $\rho$(T1→T2, T0→T2) over $\rho$(T0→T1, T1→T2) ($\Delta$0.07-0.25 under-accertainment). Two contrasts separated the architectures. First, the null adjusted onset-progression concordance itself: shared-effect architecture (B) forces a positive null-adjusted onset-progression concordance (median 0.11-0.13), whereas the observed UKB and AoU values were centred on zero (0.00), excluding substantial shared correlated effects. Second, the T1-free contrast: restricting the baseline-downstream to individuals who never develop T1 preserved the null-adjusted progression-downstream concordance only under true stage-specific effects (scenario C median 0.12-0.15, versus 0.00 under susceptibility-only A), providing the contrast against which the empirical value is read. The empirical direct and T1-free null-adjusted concordance is approximately 0.001, which corresponds to the susceptibility-only architecture (A) and not to transition-specific downstream effects (C). This is the expected result under the T2-event-overlap interpretation developed in the main text: the progression-downstream concordance of the marginal analysis is generated by the clinical route shared between the two phenotype definitions and does not persist once overlapping T2-event is removed. Shared correlated effects (B) are separately excluded by the null-adjusted onset-progression concordance, which is forced positive under B but is centred on zero in the observed data. These simulations therefore constrain which architectures are compatible with the observations; they do not establish that downstream genetic effects are absent.

#### Illness-death simulations

We simulated individual-level illness-death processes with N = 150,000 participants and 3,000 genes represented by sparse carrier sets with carrier frequencies ranging from approximately $2 \times 10^{-4}$ to $1.5 \times 10^{-2}$. Three genetic architectures were considered.

In the susceptibility-only architecture, genes affected only the T0→T1 transition. In the shared-effect architecture, onset and progression effects were correlated, with correlation 0.7. In the transition-specific architecture, distinct genes carried effects on downstream disease processes represented by T0→T2, T1→T2 and subsequent transitions.

Each architecture was simulated with and without under-ascertainment of incident T1 in the EHR and incorporated selection into the progression risk set. Simulated gene-level association statistics were passed through the same concordance pipeline used for the empirical data, including the event- and composition-matched disjoint-pair null.

The contrast between progression–marginal and onset–progression concordance was positive under several simulated architectures and therefore did not identify causal-gene sharing by itself. The original shared-effect model tended to produce more detectable onset-progression concordance than observed. The empirical near-zero progression-direct and T1-free correlations were closer to some simulated scenarios than to others, but these comparisons are scenario-specific. Because rare carrier-event information is sparse, neither a near-zero onset-progression correlation nor a near-zero progression–direct correlation can by itself establish that the corresponding causal gene sets are disjoint. The more targeted progression-direct sharing-grid simulation is reported separately below.

#### Continuum of partial sharing

The discrete simulation scenarios represent limiting cases and do not capture architectures in which only a fraction of causal genes is shared between onset and progression. We therefore extended the simulation to a continuum of shared architecture.

Sharing was parameterized as the fraction π of downstream causal genes drawn from the onset causal set. Shared genes retained their onset effect, whereas the remaining downstream causal genes were selected independently. This definition uses overlap of causal gene sets rather than correlation of per-gene effects because the primary concordance statistic is based on *P*-value ranks and is most directly influenced by which genes carry detectable association signals.

Five levels of sharing were evaluated, from $\pi = 0$ to $\pi = 1$, with 95 simulation replicates per level and the same composition-matched empirical-null procedure used in the primary concordance analyses. Median null-adjusted onset-progression concordance increased monotonically with $\pi$: −0.025 at $\pi = 0$, +0.021 at $\pi = 0.25$, +0.073 at $\pi = 0.50$, +0.108 at $\pi = 0.75$ and +0.138 at $\pi = 1.00$.

The observed UK Biobank null-adjusted onset-progression concordance was approximately 0.0008. Under this specific partial-sharing design, simulations with 25% and 50% causal-gene sharing yielded concordance at or below the observed level in approximately 22% and 1.1% of replicates, respectively. This comparison evaluates compatibility with prespecified effect sizes and carrier architectures; it is not an estimate or a confidence interval for the true shared-causal-gene fraction. The simulated architecture differs from the separate progression–direct experiment below.

To evaluate whether incomplete ascertainment of T1 could produce the same pattern, each level of π was additionally simulated with EHR under-ascertainment of the intermediate state. Under-ascertainment changed median null-adjusted concordance by less than 0.02 at every value of π, indicating that incomplete recording of T1 alone did not reproduce the empirical near-zero onset-progression pattern.

#### Progression-direct causal-sharing simulation

A separate illness-death simulation tested whether the near-zero gene-level concordance between post-diagnosis progression (B, T1→T2) and the cause-specific direct downstream transition (D, T0→T2 with censoring at incident T1) could persist when their causal genes were shared. This simulation explicitly constructed a direct arm censored at incident T1 rather than using a T1-free analysis that excluded participants with T1. The progression-direct causal-gene overlap fraction $\pi_{BD}$ was varied from 0 to 1 in increments of 0.25; at $\pi_{BD}$ = 1, direct-transition causal genes carried the same effects as the progression causal genes. These overlap fractions are distinct from the onset–progression sharing parameter in the preceding analysis.

The effect-scale parameter was calibrated to approximate the empirical B-C progression-marginal association excess and burden-effect correlation (observed approximately 0.064 and 0.287). At $\tau$ = 0.10, the corresponding simulation metrics were approximately 0.062 and 0.315, whereas at the inherited $\tau$ = 1.0 the association excess was approximately 0.21 and was not comparable to the empirical setting. At $\tau$ = 0.10, 30 replicates per sharing-grid cell were analysed, including the intermediate-state ascertainment settings. Across $\pi_{BD}$ = 0, 0.25, 0.50, 0.75 and 1.00, simulated B-D association excess was −0.0016, 0.0011, −0.0039, 0.0106 and 0.0005; the corresponding burden-effect correlations were −0.0038, −0.0137, 0.0059, −0.0049 and −0.0004. At complete sharing, pooled over ascertainment settings, the B-D association excess had median 0.0005 and burden-effect correlation had median −0.0004 (60 replicates). Thus, under the evaluated power and effects, the direct-transition metrics did not distinguish no sharing from complete sharing.

The simulation provides a detectability check, not a calibrated estimate of shared genetic architecture. Simulated B-C effect-direction concordance was 1.00 versus 0.599 in UKB, and the simulator included 3,000 genes rather than approximately 18,000 tested genes in the empirical analysis. Because few simulated genes qualified for direction-concordance comparisons, these simulated direction results were not interpreted. The overlap grid used one calibrated effect scale, 30 replicates per cell, and identical causal effects at full sharing; its conclusions should not be generalized beyond those evaluated assumptions.

### Supplementary Note 8. All of Us and cross-biobank analysis of multi-state trajectories

#### All of Us trajectory phenotype construction

Time-to-event phenotypes in All of Us were constructed from OMOP-mapped electronic health records using PheCode v1.2b1 definitions and the same T0→T1→T2 framework used in UK Biobank. For each trajectory, the upstream-onset arm (T0→T1) measured time from cohort entry to the first qualifying T1 diagnosis among participants free of T1 at entry. The marginal downstream arm (T0→T2) was defined analogously for T2 and followed participants to T2 irrespective of whether T1 occurred during follow-up. Each participant contributed the longest continuous OMOP observation interval of at least 365 days; intervals separated by a gap were not bridged, because absence of records outside an observation period cannot be read as absence of disease. Risk-set entry (baseline) was the start of that interval plus a 365-day prior-observation washout, and participants whose observation interval ended at or before baseline were dropped rather than censored at time zero. Prevalent disease was assessed against the first recorded occurrence of the PheCode over the participant's entire record, not only within the selected interval: anyone whose first occurrence fell at or before baseline was excluded. The asymmetry is deliberate, since a prior record is evidence of prevalence whereas its absence is not evidence of absence.

The progression arm (T1→T2) was restricted to participants with incident T1 who were free of T2 at the T1 date. The analysis clock was reset to the incident T1 date and measured time to first T2 diagnosis. Participants were censored at death or the end of available EHR follow-up. A 30-day landmark at cohort entry was applied to baseline-entry analyses, matching the UK Biobank incident definition. Follow-up ended at the earliest of the end of the participant's observation interval, date of death, and a site-specific administrative censoring date, taken as a minimum rather than conditionally so that a death recorded after the site cut-off cannot extend follow-up. The site-specific cut-off was the 99th percentile of condition start dates within each contributing EHR site, with each participant assigned to the site contributing most of their condition records and sites with fewer than 100 participants pooled; this is the All of Us counterpart of the per-nation hospital-inpatient cut-off used in UK Biobank.

Trajectories with at least 100 T1→T2 events within an ancestry group were included in the primary AoU trajectory analysis. Phenotypes with complete or quasi-complete separation by sex, and progression phenotypes for which the minority sex contributed fewer than 1% of events, were fitted as sex-stratified analyses with sex omitted from the covariate set. Analyses for which the null model could not be fitted were excluded and recorded in the model inventory.

#### Gene-based testing and cross-cohort harmonization

Gene-based rare-variant testing in AoU used GATE-GENE as implemented in SAIGE v1.5.2 on exonic regions of the version 8 short-read WGS callset. Models were fitted separately in EUR,

AFR and AMR using ancestry-specific sparse genetic relatedness matrices with a relatedness threshold of 0.125.

Null models were adjusted for age at transition-specific risk-set entry, sex and the first ten genetic principal components. Birth year was additionally included for progression analyses, matching the UK Biobank specification. Variant-mask definitions corresponded directly between UKB and AoU across the nine annotation-by-frequency cells used for primary gene-based association testing.

Cross-cohort phenotype harmonization required exact PheCode-pair matches. Gene symbols were harmonized using a curated HGNC alias map, including eight legacy-to-current renames such as *DFNA5* to *GSDME* and *MARCH7* to *MARCHF7*. Sixteen UK Biobank gene symbols absent from the AoU annotation were classified as untestable, and 31 symbols mapping to more than one locus in either annotation were excluded from cross-cohort analyses.

#### Cause-specific direct downstream transition

The cause-specific direct T0→T2 transition was reconstructed separately within each AoU ancestry using the same definition applied in UK Biobank. Participants entered the direct-transition risk set at cohort entry and exited at T2, censoring or incident T1, whichever occurred first. Participants with T1 prevalent at baseline were excluded. When T1 and T2 were first recorded on the same date, ordering was treated as indeterminate and the observation contributed an event to neither direct transition. A minimum of 50 direct-transition events was required.

Direct-transition models used the same GATE-GENE pipeline, covariates, variant masks and ancestry-specific sparse genetic relatedness matrices as the corresponding production analyses. Because AoU birth dates are generalized to 15 June, transition ages were calculated in days and converted using 365.25 days per year. Direct-transition results were obtained for 177, 127 and 104 trajectories in EUR, AFR and AMR, respectively; trajectories missing direct-transition results primarily fell below the event threshold after removal of intermediate-mediated events.

#### Cross-ancestry reproduction of association-concordance patterns

Within each AoU ancestry, gene-level omnibus P values were assembled into gene-by-transition matrices over genes with valid results in the relevant transitions. Association evidence was converted to ranks, and transition-pair concordance was calculated as the Pearson correlation of ranks, equivalent to Spearman's ρ. These analyses describe correlation of association statistics and do not directly estimate correlation of underlying causal genetic effects.

For each trajectory with complete results, progression-downstream concordance, $\rho$(T1→T2, T0→T2), was compared with progression-to-onset concordance, $\rho$(T1→T2, T0→T1). The paired comparison holds the progression analysis, and therefore its statistical information, constant by construction and was evaluated using the Wilcoxon signed-rank test. Eligible trajectories were also compared using B-D concordance, retaining the same progression analysis as the B-C comparison and excluding shared T2 events through censoring at incident T1.

Null-adjusted concordance was referenced to a separately constructed empirical null within each ancestry. Disjoint transition pairs sharing no disease state were matched on minimum event-count stratum and transition composition, using the same procedure as in UK Biobank. Association-concordance analyses were repeated using the cause-specific direct T0→T2 transition to test whether progression-downstream concordance attenuated after exclusion of the overlapping post-T1 T2 events.

Sensitivity analyses additionally restricted to trajectories overlapping the UK Biobank discovery set and to trajectories in which most downstream events bypassed incident T1 (**Supplementary Table 34**). Trajectory-level progression-downstream concordance estimates were also compared between UK Biobank and AoU.

#### Replication of UK Biobank associations in All of Us

The 60 trajectory-level exome-wide associations identified in UK Biobank collapsed to 20 distinct gene-transition associations because individual endpoint transitions could contribute to multiple trajectories. Replication was therefore assessed once per distinct association.

For each association, the UK Biobank-selected mask was defined as the annotation-by-frequency cell with the strongest discovery burden signal. Primary replication required concordant burden-effect direction and a one-sided $P$ <0.05 in the UK Biobank discovery direction for the same mask in AoU. A direction-agnostic Cauchy omnibus $P$ <0.05 was used as a secondary criterion but was not considered directional replication. Benjamini-Hochberg correction was applied across carried-forward associations within each ancestry.

Sensitivity analyses repeated replication using the best corresponding AoU mask and after restricting the UK Biobank-selected mask to cells meeting the prespecified minor-allele-count requirement. A fixed-effect inverse-variance meta-analysis across AoU EUR, AFR and AMR provided one trans-ancestry AoU replication test per association, with heterogeneity summarized using Cochran's $Q$ and $I^2$.

#### Cross-biobank meta-analysis

Because variant-level score statistics and linkage-disequilibrium summaries could not be exchanged across the UK Biobank and AoU research environments, cross-biobank meta-analysis used gene- and mask-level burden summaries.

For each harmonized transition analysis, gene and annotation-by-frequency mask, burden log-hazard-ratio estimates were combined across four strata–UK Biobank EUR and AoU EUR, AFR and AMR–using fixed-effect inverse-variance meta-analysis. Mask-level meta-analysis $P$ values were then combined across masks using the Cauchy method to obtain a gene-level cross-biobank burden-meta omnibus *P* value. SKAT and SKAT-O statistics were not meta-analysed.

The cross-biobank analysis was treated as combined discovery rather than replication and formed a separate multiple-testing family. Exome-wide significance was defined as $P < 2.5 \times 10^{-6}$ per transition. New progression associations were evaluated using leave-one-stratum-out fixed-effect meta-analysis, DerSimonian-Laird random-effects meta-analysis, direction concordance across contributing strata and event-count-weighted Stouffer *P*-value combination. Calibration was assessed from the cross-biobank burden-meta omnibus distribution within each transition.

Associations identified by meta-analysis were classified according to their corresponding UK Biobank evidence as already exome-wide significant, suggestive, tested without signal or untestable. Corresponding onset results were additionally examined to determine whether progression discoveries would have been identified through onset-based association analyses.

#### Index-event bias

Progression risk sets condition on reaching T1. Such conditioning can induce a spurious negative correlation between onset and progression effects, known as index-event bias, which could in principle contribute to the low concordance observed between T0→T1 and T1→T2 analyses. We therefore tested the direction of this bias in both cohorts. If index-event bias explained the separation between onset and progression, onset-progression effect correlations would be expected to be systematically negative.

In UK Biobank, the median onset-progression correlation of burden-effect estimates among jointly testable masks was 0.028 (IQR, 0.010-0.047) across 95 trajectories, and the median fraction of concordant effect signs was approximately 0.51. In AoU, among 849 UKB onset-suggestive gene-trajectory comparisons, the onset and progression burden-effect directions agreed in 53% of comparisons (51% in EUR, 56% in AFR and 52% in AMR). These observations did not reveal a systematically negative average effect correlation or sign deficit. However, negative correlation is not a necessary feature of index-event bias for every genetic architecture; these uncorrected diagnostics do not rule out selection bias, quantify its magnitude, or establish that progression effects are unbiased.

#### Power-based replication expectation and winner's-curse attenuation

For each testable UK Biobank association, the expected probability of primary replication in AoU was computed as $1 - \Phi(z_{0.95}0.95 - f \times |\beta_{UKB}\mathrm{UKB}| / \mathrm{SE}_{\mathrm{AoU}})$, where $f$ denotes attenuation of the UK Biobank discovery effect. The expected number of replications was the sum of these probabilities across analyzable associations. To assess robustness to winner's-curse inflation of discovery effects, we evaluated $f$ = 1, 0.75, 0.5 and 0.25.

Observed-to-expected replication ratios were below 1 in every ancestry and under every attenuation factor. At $f$ = 1, 0.75, 0.5 and 0.25, observed-to-expected ratios were 0.42, 0.42, 0.45

and 0.62 in EUR; 0.19, 0.20, 0.24 and 0.41 in AFR; and 0.15, 0.16, 0.20 and 0.35 in AMR. Even under $f$ = 0.25, corresponding to 75% attenuation of discovery effects, the observed-to-expected ratio remained below 1 in every ancestry, indicating that incomplete replication is not attributable to discovery-effect inflation alone.

Power-bin analyses led to the same conclusion. Across association-by-ancestry pairs deduplicated to distinct associations, 48 pairs with estimated power $f \geq 80\%$ yielded 13 observed primary replications against 47.3 expected replications, whereas two lower-power pairs yielded no observed replications against 0.9 expected. Replication counts were also robust to mask definition. Using the UK Biobank-selected mask, 8, 3 and 2 associations met the primary criterion in EUR, AFR and AMR, respectively. Restricting to mask cells with MAC $f \geq 20$ gave 8, 3 and 1 associations, whereas scoring on the best corresponding AoU mask gave 11, 7 and 4 associations. The latter analysis is anti-conservative because it permits best-mask selection in AoU, but it shows that incomplete transferability is not driven solely by exact-mask mismatch. In the trans-ancestry AoU replication meta-analysis, between-ancestry heterogeneity was modest overall, with median $I^2$ = 14%; Cochran's $Q$ was significant at $P < 0.05$ for 3 of 16 associations estimated in more than one ancestry.

#### Suggestive-tier transferability and sign-concordance artefact

We analysed the broader suggestive tier of 872 UK Biobank associations with $P < 1 \times 10^{-4}$ separately from exome-wide discoveries. Direction-concordant nominal replication, defined using one-sided $P < 0.05$ on the UK Biobank-selected mask, occurred at 3.9-fold enrichment over the 2.5% chance expectation in EUR ($P = 8.3 \times 10^{-24}$), 3.0-fold enrichment in AFR ($P = 4.0 \times 10^{-12}$) and 0.9-fold enrichment in AMR ($P$ = 0.75). Enrichment increased monotonically with the strength of UK Biobank evidence: 8.1-fold for associations with discovery-level $P < 2.5 \times 10^{-6}$, 2.6-fold for $P = 2.5 \times 10^{-6}$ to $1 \times 10^{-5}$ and 1.6-fold for $P = 1 \times 10^{-5}$ to $1 \times 10^{-4}$. Progression associations also transferred above chance (2.2-fold enrichment for T1→T2), within the range observed for onset (2.8 fold for T0→T1) and baseline-downstream (2.7-fold for T0→T2) associations.

Raw sign concordance was not used as a replication metric because, in sparse masks, effect signs reflected allele count rather than shared biology. In UK Biobank, 98% of suggestive effects were risk-increasing, consistent with winner's-curse selection at a fixed significance threshold. In AoU, sparse masks were biased toward protective estimates because masks with few or no carrier events tend to yield negative log-hazard ratios. Accordingly, only 28% of matched-mask AoU effects were positive at MAC <10, compared with 51% at MAC ≥200. A logistic regression of sign agreement on log10 MAC showed a strong trend ($P = 1.1 \rightarrow 10^{-13}$), indicating that raw sign agreement tracks allele count and is not an appropriate measure of transferability in this setting.

#### Native All of Us discovery, transition calibration and cross-biobank meta-analysis sensitivity

Native exome-wide analyses within AoU were analysed separately from both the replication look-up and the cross-biobank meta-analysis. Using the per-gene Cauchy omnibus threshold $P$ < 2.5

$\times 10^{-6}$ and restricting to calibrated analyses, native AoU discovery yielded 40, 16 and 8 associations in EUR, AFR and AMR, respectively. When classified against UK Biobank, five native AoU associations restated UK Biobank exome-wide associations involving *PKD1*, *LDLR* and *BRCA2*; ten lay in trajectories or genes that UK Biobank could not test; and 32 were tested in UK Biobank without signal. The latter two categories represent candidate cohort- or ancestry-specific associations and are reported with effect estimates from both resources side by side.

The strongest native AoU association was a positive-control-like AFR signal: protective PCSK9 burden against hypercholesterolaemia ($P = 1.8 \times 10^{-14}$; minor-allele count 1,179; burden-score log-HR = -0.025), a canonical African-ancestry signal that European-ancestry cohorts are largely unable to detect. Calibration was assessed using $\lambda_{0.01}$, as in the UK Biobank analyses, because the median genomic-control factor of sparse gene-based tests is systematically deflated and can miss tail inflation. All EUR and AFR analyses were calibrated with $\lambda_{0.01} \leq 1.2$. In AMR, eight analyses showed tail inflation and contributed 139 apparent associations; these analyses were flagged and their associations were excluded from all reported discovery counts. Analyses used transitions with successfully fitted null models; sex-specific phenotypes for which null models could not be fitted were excluded throughout.

The cross-biobank meta-analysis was well calibrated in the tail, with $\lambda_{0.01}$median 1.08 and maximum 1.17 across 155 transition pairs. Beyond the progression results reported in the main text, the joint analysis recovered eight onset associations already exome-wide in UK Biobank and identified 36 additional onset associations. Sensitivity analyses supported the 12 new progression associations from the cross-biobank meta-analysis. The maximum leave-one-stratum-out *P* value was $2.7 \times 10^{-3}$. Event-count-weighted Stouffer *P*-value combination gave $P \leq 4.3 \times 10^{-5}$ for all 12 associations and ranked associations consistently with the inverse-variance meta-analysis, with Spearman $\rho = 0.74$ among associations with joint $P < 1 \times 10^{-5}$. The three UK Biobank-only progression candidates did not reach the joint $P < 1 \times 10^{-4}$ reporting threshold, consistent with winner's-curse attenuation.

#### Cross-biobank transferability and clinical validation of progression associations

To assess whether the downstream events underlying the UKB progression candidates represented clinically meaningful events rather than surveillance or coding artefacts, we corroborated outcome records against temporally proximate objective clinical information. Among assessable myocardial infarction events, 48% were accompanied by coronary revascularization, coronary angiography or a cardiac cause of death within 90 days and 58% within one year, with procedures concentrated around the recorded event date. Among urinary tract infection events in participants with linked primary-care prescribing data, 40% were accompanied by an antibiotic prescription within 30 days and 80% within one year. Dialysis events showed similarly strong corroboration, with 64% accompanied by a renal-replacement procedure within 90 days and 81% within one year. We further tested whether differential healthcare utilization could account for the *F13A1* and *GSDME* signals by adjusting carrier models for pre-entry coding intensity and by conservatively censoring uncorroborated downstream events. Effect estimates were unchanged after adjustment for coding intensity and increased after restricting to corroborated events (*GSDME* log-HR 2.19→2.25, $P = 8.5 \times 10^{-9}$; *F13A1* 0.56→0.84, $P = 6.3 \times 10^{-4}$), indicating that carrier events were disproportionately supported by independent clinical records. These analyses argue against endpoint misclassification or surveillance intensity as the primary explanation for the UKB associations and support interpreting the lack of *GSDME* replication in AoU as limited cross-cohort transferability rather than evidence that the UKB endpoint was artefactual.

We additionally performed clinical-code sensitivity analyses to determine whether the PKD1 renal trajectory associations reflected progression of common hypertension or chronic kidney disease or instead captured underlying polycystic kidney disease. Cystic kidney disease was defined using ICD-9 753.1x and ICD-10 Q61.x codes. Excluding participants with any recorded cystic-kidney-disease code removed only 1.4-5.6% of each AoU risk set but 56-100% of qualifying *PKD1* pLoF alleles in the lead mask; applying the same exclusion in UKB eliminated the corresponding lead-mask meta-analysis signals for hypertension→CKD and CKD→dialysis ($P = 0.74$ and 0.30, respectively). In UKB, five of seven lead-mask carriers contributing to the two progression risk sets had cystic-kidney-disease codes despite background frequencies of only 0.2–1.1%, and all four carriers in the CKD→dialysis risk set progressed to dialysis. When exclusion was restricted to cystic-kidney-disease codes recorded by 90 days after T1, residual association remained ($P = 3.5 \times 10^{-4}$ and $9.0 \times 10^{-7}$, respectively), indicating that some carriers were first recognized as having cystic kidney disease only after entering the progression risk set. Carrier-level review in UKB showed that several first cystic diagnoses occurred 0.9-2.8 years after T1, including one after the downstream event itself. Together, these analyses indicate that the *PKD1* trajectory signals primarily capture previously unrecognized polycystic kidney disease rather than a general modifier of non-cystic renal progression. This finding highlights a complementary role of stage-resolved trajectory analysis: in addition to identifying progression-associated genetic effects, it can reveal latent monogenic disease embedded within common longitudinal disease pathways.

#### Ancestry-specific transferability and the AFR replication deficit

Replication of UK Biobank discoveries was consistently weakest in AFR. To distinguish technical from biological explanations, we compared the UK Biobank-selected mask between EUR and AFR for the 15 associations in which the selected mask existed in both ancestries. The AFR mask captured approximately half the cumulative minor-allele count of the EUR mask, with a median AFR/EUR MAC ratio of 0.52 (Wilcoxon $P = 0.0057$), reflecting smaller AFR risk sets. Per event, however, AFR masks were not systematically poorer: the median AFR/EUR MAC-per-event ratio was 1.5 (Wilcoxon $P = 0.17$). Because statistical power in rare-variant burden tests depends on both carrier count and event information, this pattern indicates that the AFR replication deficit principally reflects smaller progression risk sets rather than uniformly poorer variant capture per event.

Gene-specific exceptions were present. For *SRSF2*, *PATL1* and *GCK*, the UK Biobank-selected masks captured few or no qualifying AFR variants. For these genes, annotation and variant ascertainment, rather than lack of biological transferability, limit cross-ancestry replication. Thus, the overall AFR replication deficit appears to reflect smaller progression risk sets, with a subset of gene-specific cases additionally limited by sparse qualifying variation.

### Supplementary Figures

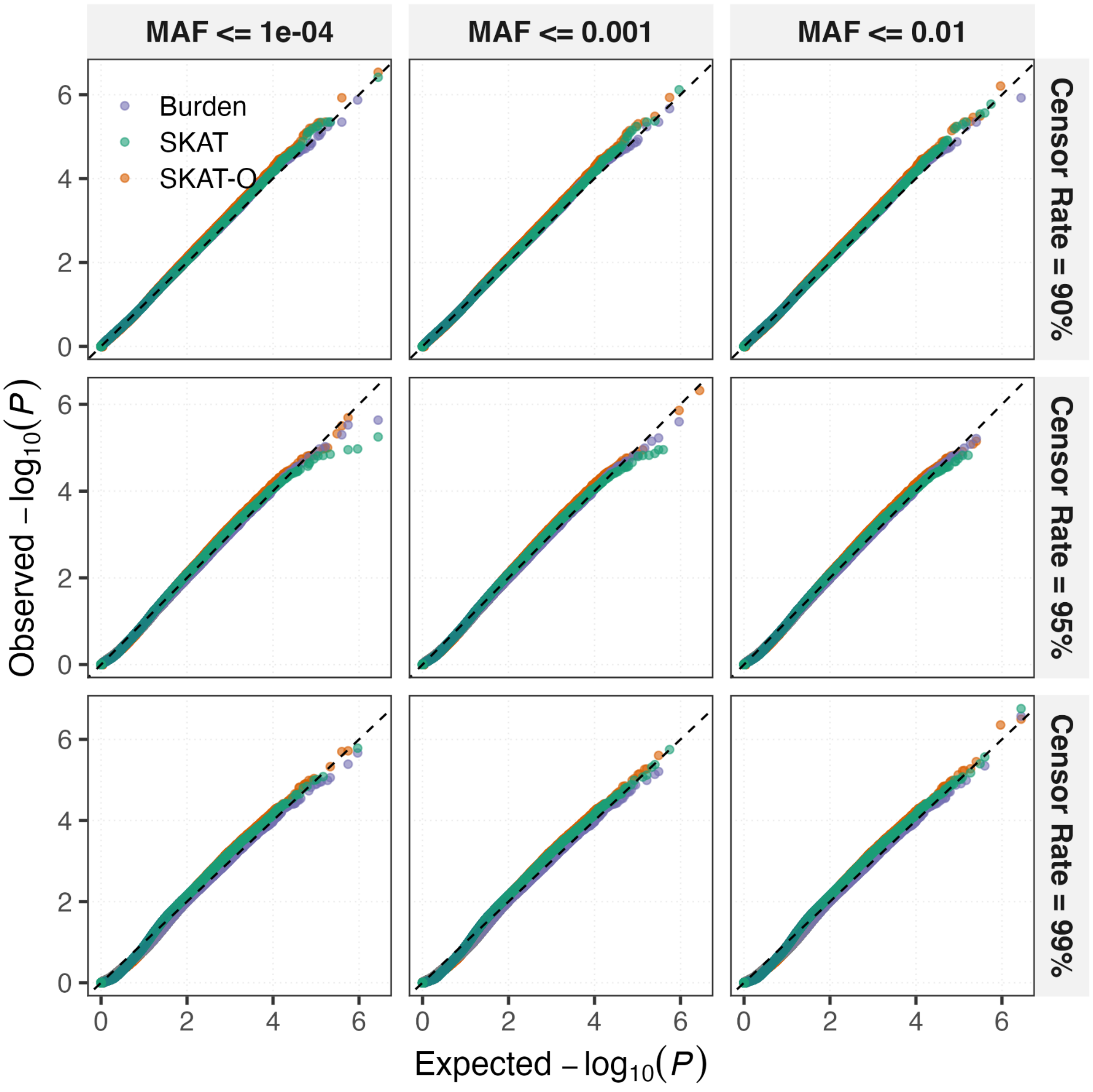


#### Supplementary Fig. 1 | Type I error calibration of GATE-GENE in simulations with saddlepoint approximation.

Quantile-quantile plots of gene-level *P*-values under the null for burden (purple), SKAT (green) and SKAT-O (orange), across mask MAF ceilings and censoring rates. Simulations used UK Biobank exome genotypes for 100,000 White British participants (Methods; Supplementary Note

2). Across 4.17 million null tests per statistic, the empirical type I error at $\alpha = 2.5 \times 10^{-6}$ was approximately nominal ($\leq 2.9 \times 10^{-6}$; interpret with exact binomial confidence intervals).

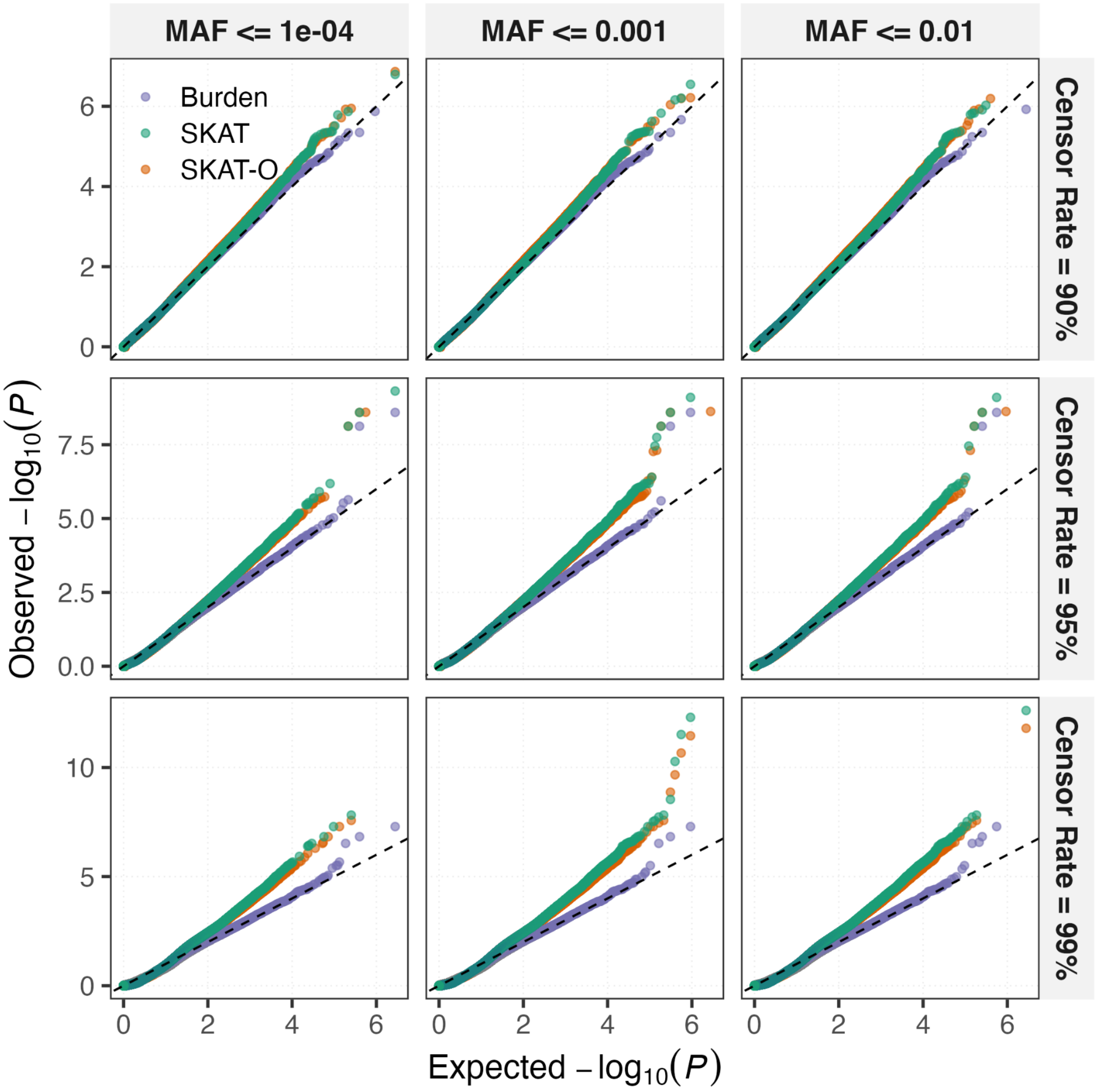


Supplementary Fig. 2 | Type I error without saddlepoint approximation.

As Supplementary Fig. 1, but with the saddlepoint approximation disabled. SKAT and SKAT-O show type I error inflation that increases with censoring, whereas burden remains approximately calibrated (empirical type I error at $\alpha = 2.5 \times 10^{-6}$; $5.0 \times 10^{-5}$ for SKAT and $3.8 \times 10^{-5}$ for SKAT-O, versus $5.0 \times 10^{-6}$ for burden), demonstrating that saddlepoint calibration is essential for variance-component tests under heavy censoring.

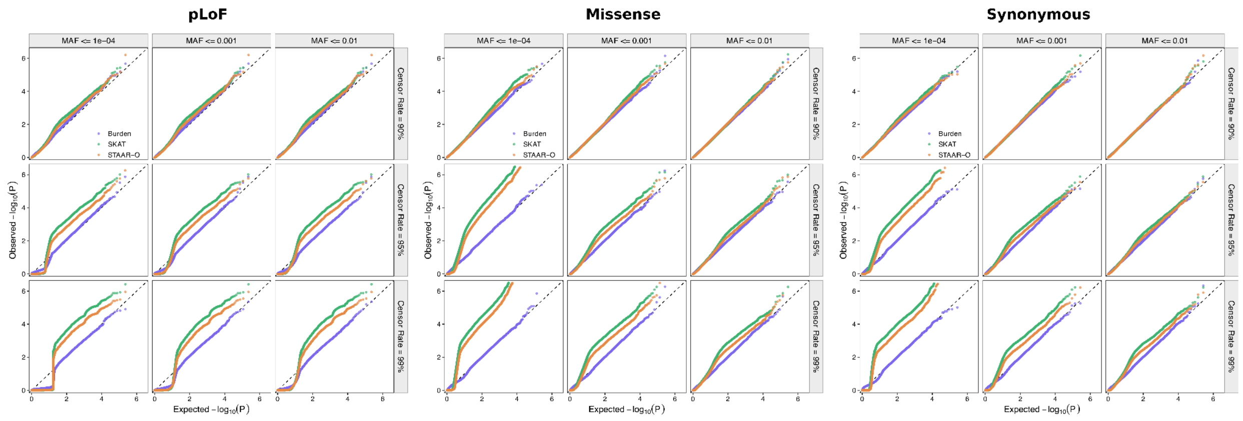


#### Supplementary Fig. 3 | Type I error of GATE-STAAR under increasing censoring.

Empirical type I error of GATE-STAAR burden, SKAT and STAAR-O tests was evaluated using UK Biobank exome genotypes from 100,000 White British participants under simulated null time-to-event phenotypes with 90%, 95% and 99% censoring. Dashed horizontal lines indicate the nominal type I error rate.

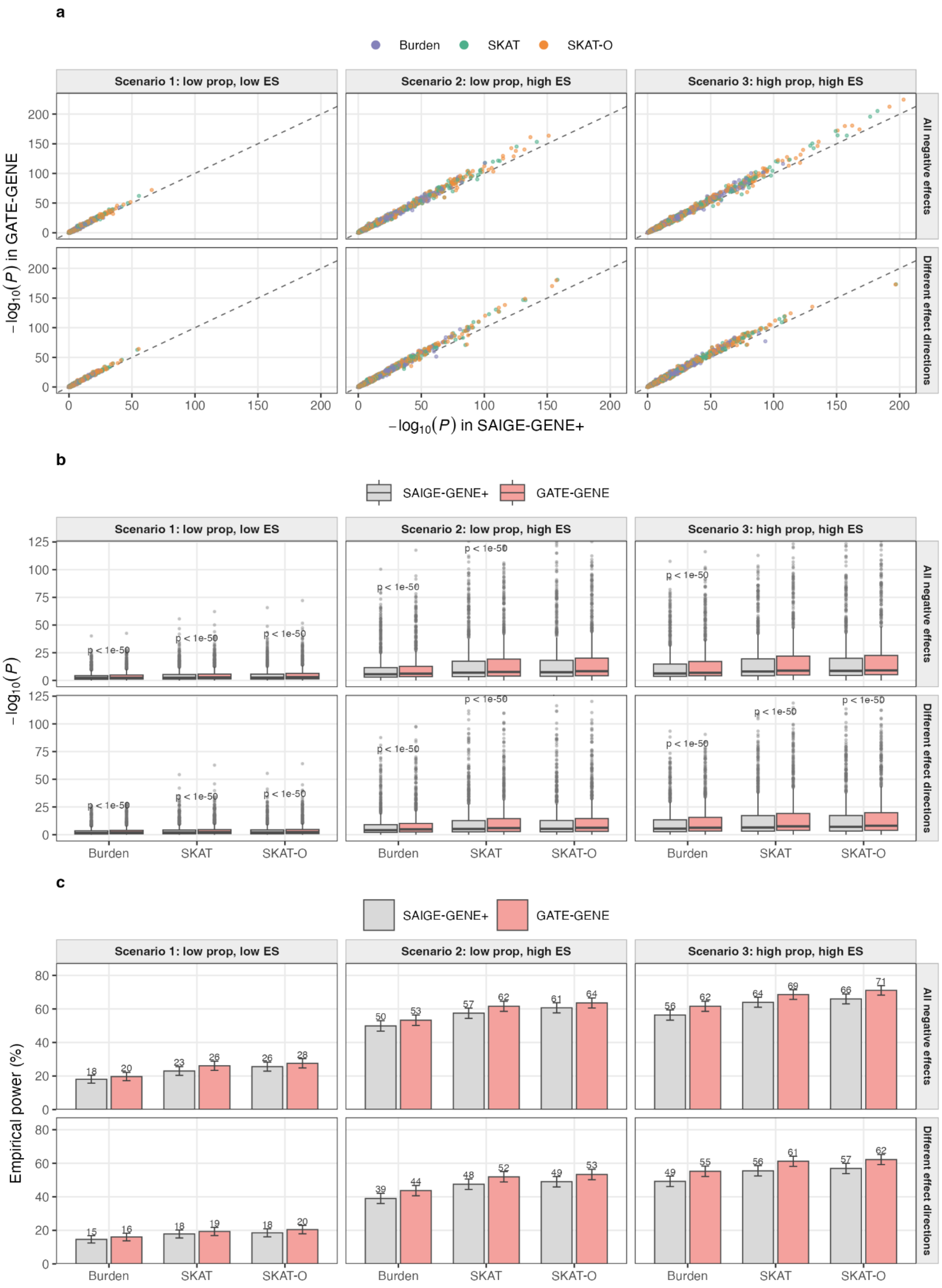
a
Burden
SKAT
SKAT-O
Scenario 1: low prop, low ES
Scenario 2: low prop, high ES
Scenario 3: high prop, high ES
All negative effects
Different effect directions
$-\log_{10}(P)$ in GATE-GENE
$-\log_{10}(P)$ in SAIGE-GENE+
b
SAIGE-GENE+
GATE-GENE
$-\log_{10}(P)$
p < 1e-50
Burden
SKAT
SKAT-O
c
SAIGE-GENE+
GATE-GENE
Empirical power (%)
Burden
SKAT
SKAT-O

#### Supplementary Fig. 4 | Empirical power of GATE-GENE versus SAIGE-GENE+ at 90% censoring.

a, Per-gene $-\log_{10}(P)$ for GATE-GENE against SAIGE-GENE+ for burden (purple), SKAT (green) and SKAT-O (orange) at causal genes, under three causal-architecture scenarios (columns; **Supplementary Table 5**) and two effect-direction settings (rows); the dashed line marks equality.

b, Distributions of $-\log_{10}(P)$ by test for SAIGE-GENE+ (grey) and GATE-GENE (red); annotations give Wilcoxon rank-sum tests comparing the two methods (all $P < 1 \times 10^{-50}$). Median $-\log_{10}(P)$ values are summarized in **Supplementary Table** 6. c, Empirical power, the proportion of causal-gene tests reaching the exome-wide threshold ($P < 2.5 \times 10^{-6}$), for each test, scenario and effect-direction setting (1,000 causal-gene tests per cell); bars show the point estimate with 95% confidence intervals and labels give the percentage. Power values and paired tests are given in **Supplementary Table 6**.

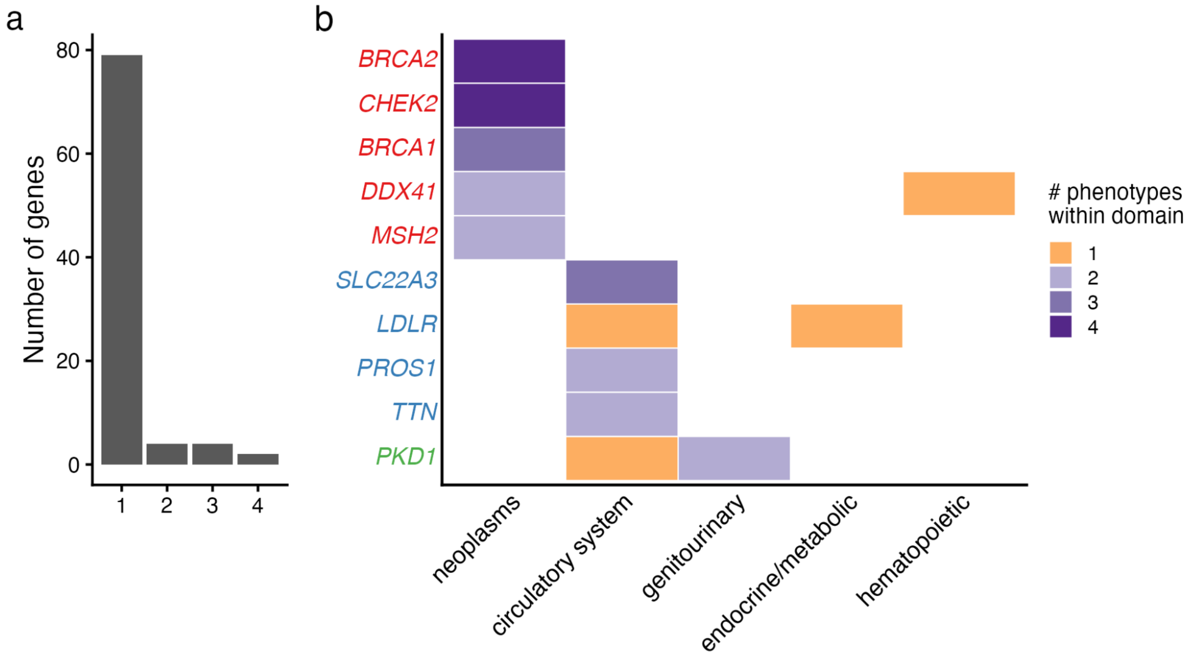


#### Supplementary Fig. 5 | Pleiotropy of rare-variant onset associations across the UK Biobank phenome.

a, Distribution of gene-level pleiotropy among exome-wide significant onset associations. Bars show the number of genes associated with a given number of onset phenotypes. b, Clinical "fingerprints" of highly pleiotropic genes. The heatmap shows, for selected genes with the largest number of associated phenotypes, the number of associated onset phenotypes within each PheCode category (binned as 1, 2, 3, 4; blank indicates none). Gene labels are colored by broad gene groups inferred from the dominant category of associated phenotypes.

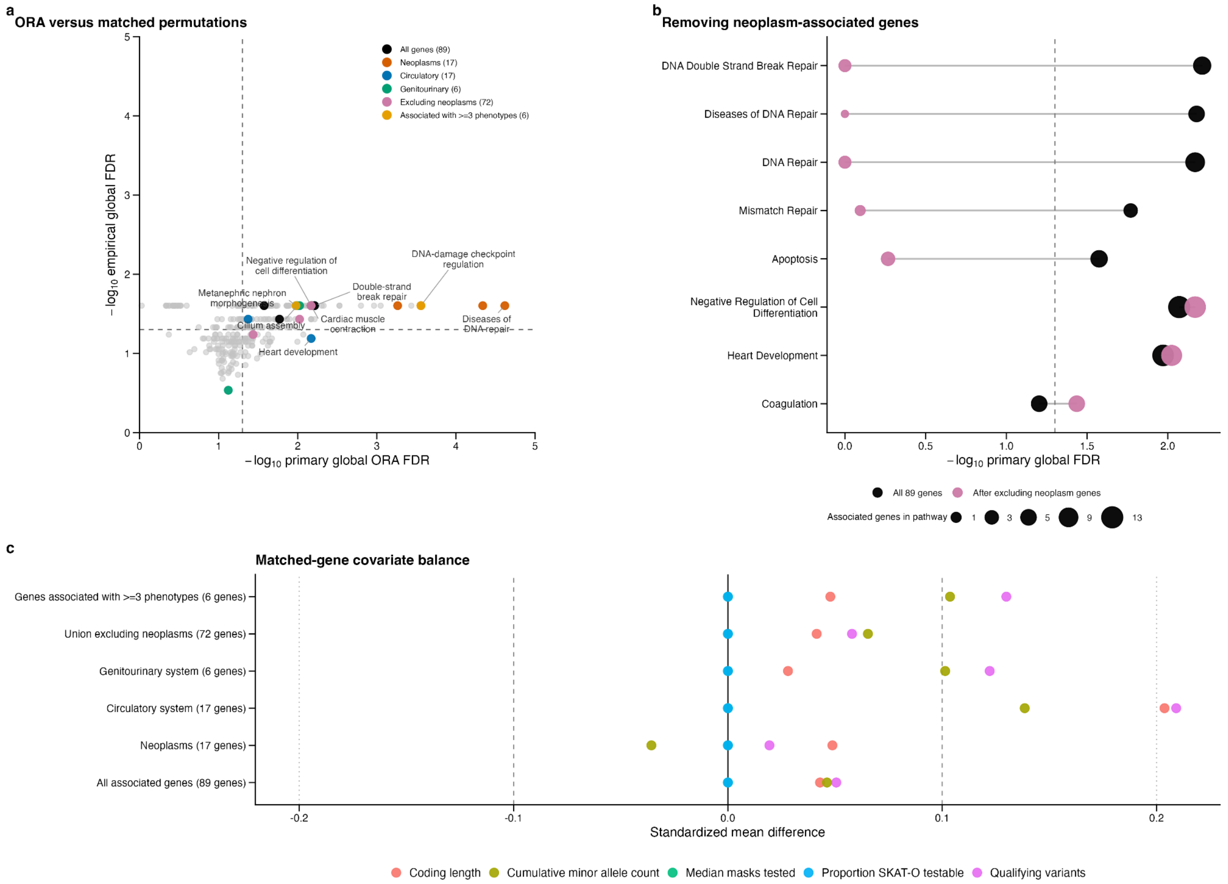


#### Supplementary Fig. 6 | Robustness and calibration of pathway enrichment analyses

a, Comparison of pathway enrichment evidence from the primary over-representation analysis and covariate-matched gene-set permutations. Each point represents a pathway-gene-list combination with at least one associated gene in the pathway and global FDR < 0.10 in either analysis. The x-axis shows $-\log_{10}$(primary global ORA FDR) and the y-axis shows $-\log_{10}$(empirical global FDR) from the matched-gene permutations. Colored points denote highlighted pathway-gene-list combinations, with colors indicating the associated-gene list; other qualifying combinations are shown in grey. Selected representative pathways are labelled. Dashed lines indicate FDR = 0.05. b, Sensitivity of selected union-level pathway enrichments to exclusion of the 17 neoplasm-associated genes. Black points show results for the complete 89-gene union and pink points show results after excluding neoplasm-associated genes. Points representing the same pathway are connected by grey lines. The x-axis shows $-\log_{10}$(primary globa FDR), and point area represents the number of associated genes assigned to the pathway. The dashed line indicates FDR = 0.05. c, Balance of gene-level characteristics between each associated-gene list and its matched control-gene sets. Points show standardized mean differences for coding length, qualifying-variant count, cumulative minor allele count, median number of masks tested and the proportion of phenotypes with a testable SKAT-O result. The solid vertical line indicates perfect balance, dashed lines indicate standardized mean differences of $\pm 0.10$ and dotted lines indicate $\pm 0.20$. Values closer to zero indicate better covariate balance.

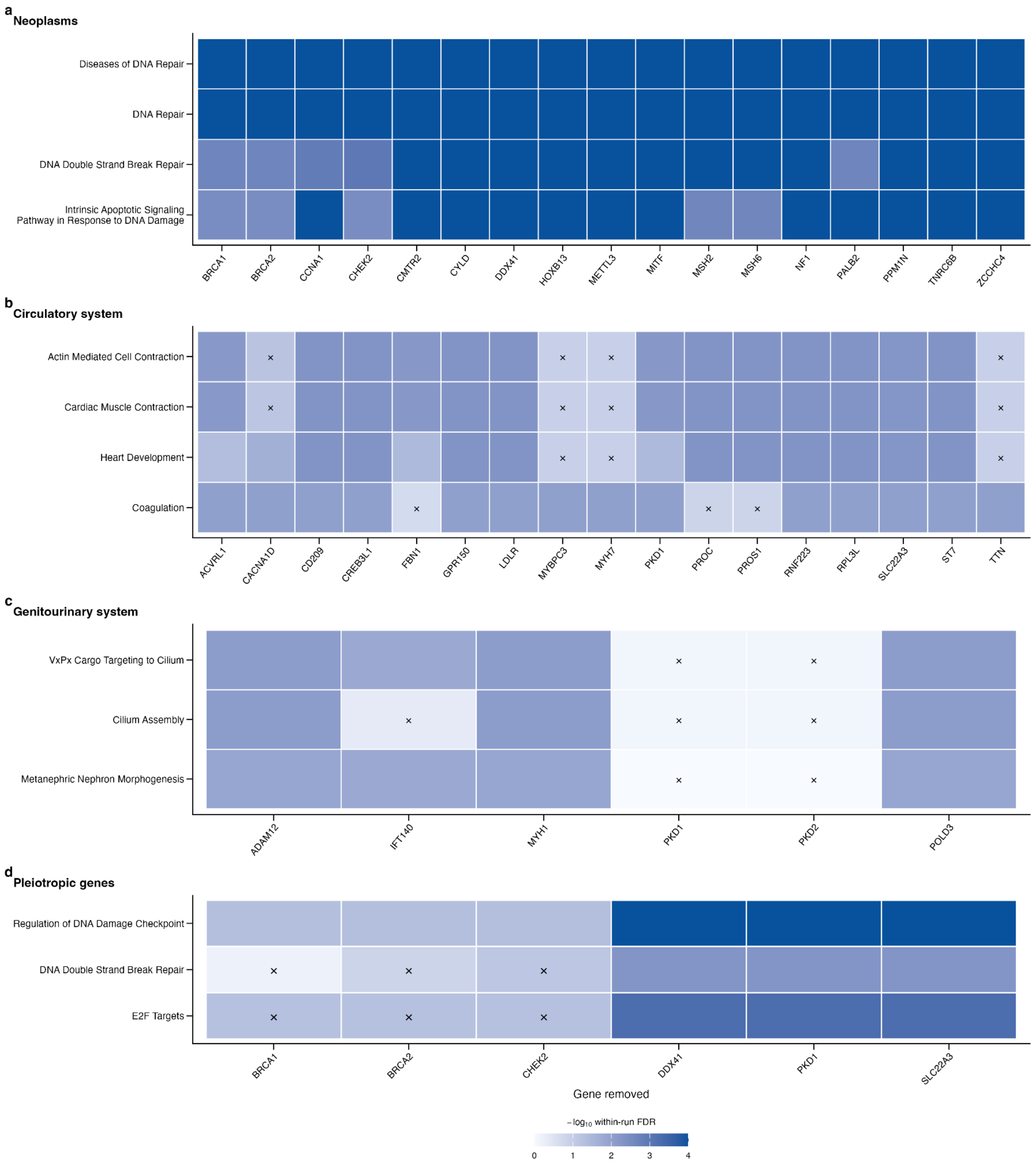


Supplementary Fig. 7 | Leave-one-gene-out assessment of pathway enrichment robustness.

a-d, Leave-one-gene-out sensitivity analyses for pathways enriched among genes associated with neoplasm phenotypes (a), circulatory-system phenotypes (b), genitourinary-system phenotypes (c) and at least three phenotypes (d). Each column represents an analysis in which the indicated gene was removed from the corresponding associated-gene list before repeating the over-representation analysis. Rows show representative pathways reported in the main analysis. Tile color represents $-\log_{10}$ of the within-run FDR, with darker colors indicating stronger evidence of enrichment. Crosses indicate pathway results with within-run FDR $\geq 0.05$ after removal of the indicated gene. Persistence of significant enrichment across columns indicates that the pathway signal is not dependent on any single gene, whereas loss of significance after removal of particular genes indicates that the enrichment is concentrated in those genes.

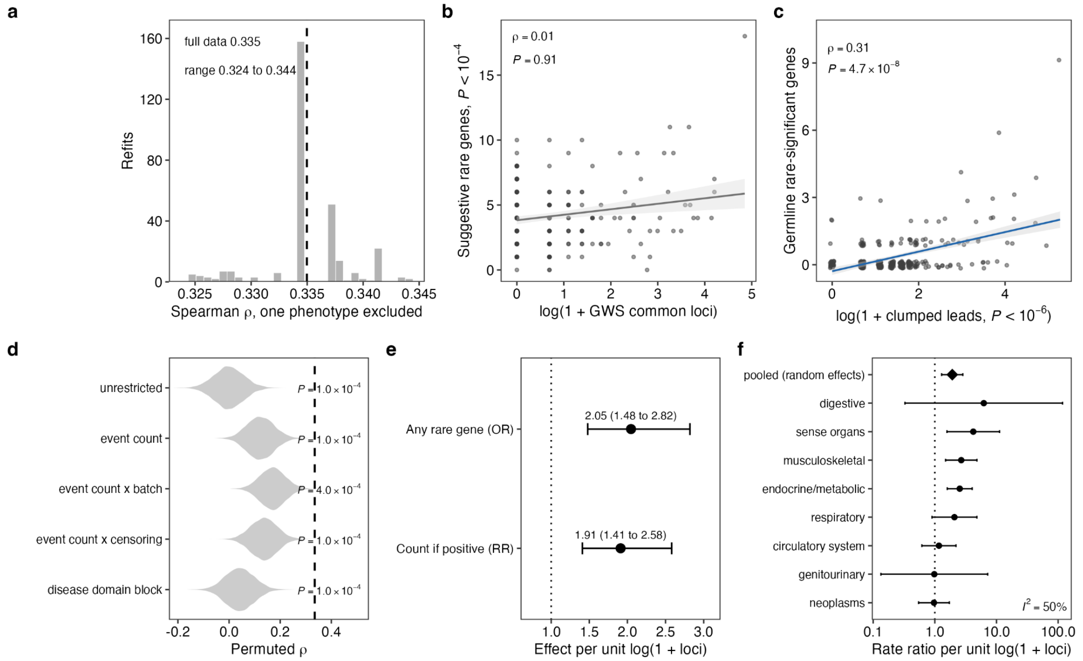


#### Supplementary Fig. 8 | Robustness of the phenotype-level convergence between common-variant locus burden and germline rare-gene discovery.

a, Distribution of the Spearman correlation between genome-wide significant (GWS) common-locus count and germline rare-significant gene count across 300 leave-one-phenotype-out refits (range 0.324 to 0.344, median 0.334); dashed line, full-data value ($\rho$ = 0.335). b, Germline rare-gene counts at the suggestive threshold ($P < 1 \times 10^{-4}$) against GWS common-locus count ($\rho$ = 0.01, $P$ = 0.91); line, linear fit; shading, 95% confidence band. c, Germline rare-significant gene count against all clumped common lead variants ($P < 1 \times 10^{-6}$) rather than GWS leads only ($\rho$ = 0.31, $P = 4.7 \times 10^{-8}$, n = 300); points are jittered; line, linear fit; shading, 95% confidence band. d, Null distributions of the Spearman correlation under five permutation designs of increasing stringency, each 10,000 permutations: unrestricted label permutation; permutation within event-count quintile; within event-count quintile crossed with analysis batch; within event-count quintile crossed with censoring-rate tertile; and block permutation within PheCode disease domain. Dashed line, observed correlation; $P_{perm}$ is the one-sided empirical $P$, floored at $1 \times 10^{-4}$ by the number of permutations. e, Hurdle-model components: the odds ratio for yielding any germline rare-significant gene (logistic regression, n = 300) and the rate ratio for the number of genes among rare-positive phenotypes (zero-truncated negative binomial, n = 67). Both are per one-unit increase in log(1 + GWS common-locus count) and adjusted for log(1 + event count). Points, point estimates; horizontal bars, 95% Wald confidence intervals; dotted line, no effect. f, Negative binomial rate ratios estimated separately within each of the eight disease domains with sufficient events, with 95% confidence intervals, and the DerSimonian-Laird random-effects pooled estimate (diamond; rate ratio 1.92, 95% CI 1.29 to 2.85, $P = 1.2 \times 10^{-3}$). Heterogeneity $I^2 = 50\%$ ($Q = 14.0$ on 7 degrees of freedom). Dotted line, no effect; x axis on a log scale.

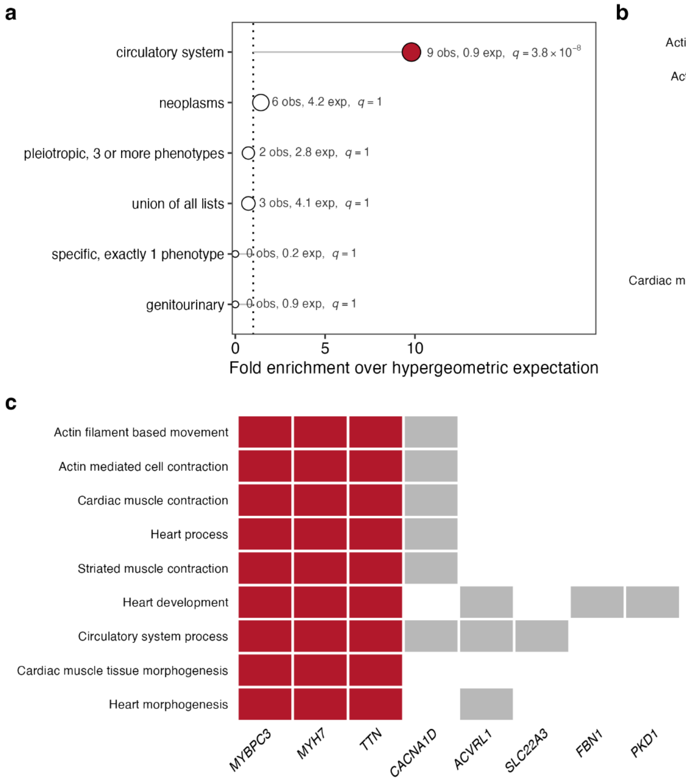


#### Supplementary Fig. 9 | Pathway convergence is confined to circulatory phenotypes and to a small sarcomeric gene core.

a, Overlap between rare- and common-variant Gene Ontology Biological Process enrichment for each prespecified rare-variant gene list, showing the observed and hypergeometric expected number of shared terms and the Benjamini-Hochberg $q$ value. Point size, number of shared terms; filled red, $q < 0.05$; dotted line, no enrichment. Six of the ten prespecified lists yielded a testable overlap and are shown; the remaining four produced no significant rare-side term and contribute $P = 1$ to the correction. b, The nine Gene Ontology Biological Process terms shared between the rare- and common-variant circulatory analyses, grouped into six redundancy clusters; blue, contraction; orange, development. c, Gene-by-term membership for the eight genes contributing to the rare-variant enrichment of the shared terms; red, *TTN*, *MYH7* and *MYBPC3*, which account for 27 of the 39 memberships. d, Number of redundancy clusters retaining at least two contributing rare-side genes with all genes included, after removing *TTN*, and after removing *TTN*, *MYH7* and *MYBPC3*.

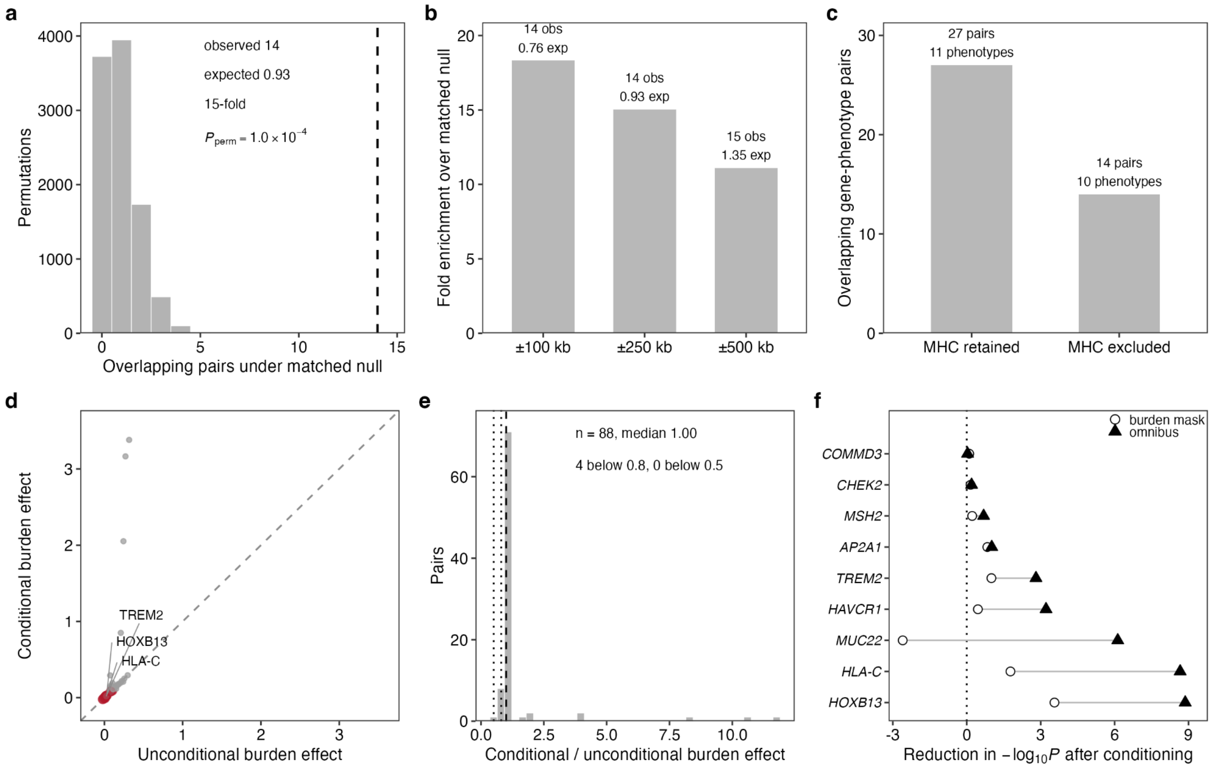


#### Supplementary Fig. 10 | Robustness of the positional overlap and of the conditional analysis.

a, Covariate-matched permutation null for the number of overlapping gene-phenotype pairs within ±250 kb of a genome-wide significant (GWS) common lead variant, extended MHC excluded. Dashed line, observed count. b, Observed and expected overlaps and the resulting fold enrichment over the matched null at ±100 kb, ±250 kb and ±500 kb. c, Number of overlapping gene-phenotype pairs with the extended MHC (chr6:25 to 34 Mb) retained and excluded; 13 of the 27 pairs recovered when the MHC is retained lie within it and arise from two phenotypes. d, Conditional against unconditional rare-variant burden effect estimates for all 90 germline-likely gene-phenotype pairs with stable conditional fits (Spearman $\rho$ = 0.98); red, the 12 pairs that also show positional overlap; dashed line, identity. e, Distribution of the 88 defined conditional-to-unconditional effect ratios, that is, the 90 stable pairs excluding two whose effect changed sign (median 1.00, interquartile range 0.98 to 1.02; four ratios below 0.8 and none below 0.5). Dashed line at 1.0; dotted lines at 0.8 and 0.5. f, Reduction in $-\log_{10}P$ after conditioning for the selected burden mask (open circles) and for the omnibus Cauchy statistic (filled triangles), paired within gene-phenotype pair, for the nine stable pairs that fell below the exome-wide threshold. The omnibus statistic weakened more than the burden statistic in eight of nine pairs (median reduction 2.81 versus 0.45); dotted line, no change.

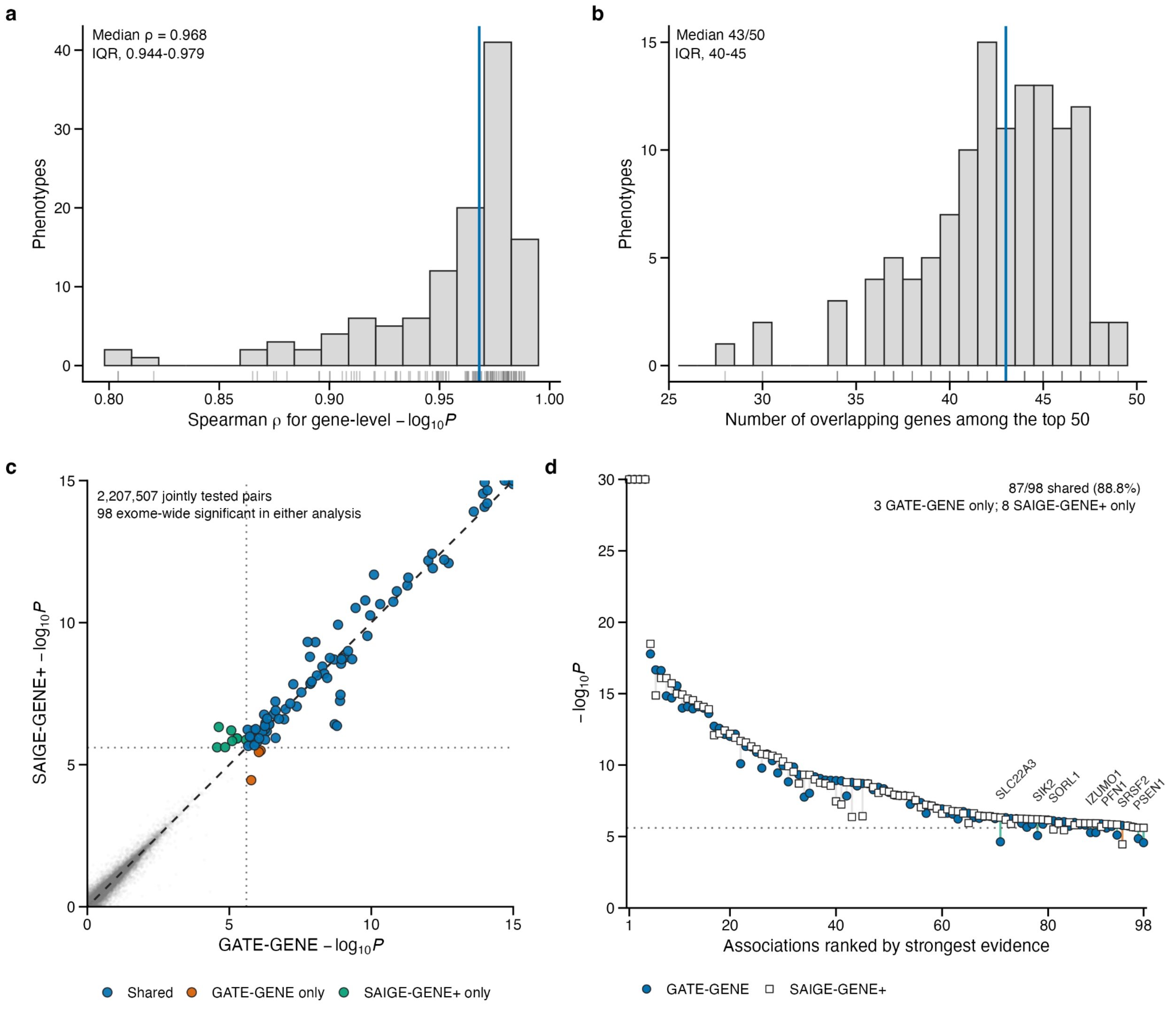


#### Supplementary Fig. 11 | Concordance of incident rare-variant association signals between GATE-GENE and SAIGE-GENE+.

a, Distribution of phenotype-specific Spearman correlations between gene-level values from GATE-GENE and matched incident case-control analyses using SAIGE-GENE+ across 120 phenotypes. The blue line denotes the median correlation (ρ = 0.968; IQR, 0.944-0.979), and rug marks denote individual phenotypes. b, Distribution of the number of genes shared between the top 50 genes ranked by GATE-GENE and SAIGE-GENE+ for each phenotype. The blue line denotes the median overlap of 43 genes (IQR, 40-45), and rug marks denote individual phenotypes. c, Gene-level association evidence from GATE-GENE and SAIGE-GENE+ across jointly tested gene-phenotype pairs. A reproducible random sample of 100,000 associations that were not exome-wide significant in either analysis is shown in grey, and all 98 associations reaching exome-wide significance in at least one analysis are overlaid. Blue, orange and green points denote associations significant in both analyses, in GATE-GENE only and in SAIGE-GENE+ only, respectively. The dashed line indicates equality, and dotted lines denote the exome-wide significance threshold. Values are capped at -$\log_{10}P$ = 15 for visualization. D, Paired GATE-GENE and SAIGE-GENE+ association evidence for the 98 gene-phenotype associations reaching exome-wide significance in either analysis. Associations are ordered by the stronger of the two signals; paired values for the same association are connected. Filled circles denote GATE-GENE results and open squares denote SAIGE-GENE+ results. Of the 98 associations, 87 were significant in both analyses, 3 were significant only in GATE-GENE and 8 only in SAIGE-GENE+. Discordant associations are highlighted and labeled by gene. The dotted line denotes the exome-wide significance threshold, and values are capped at -$\log_{10}P$ = 30 for visualization.

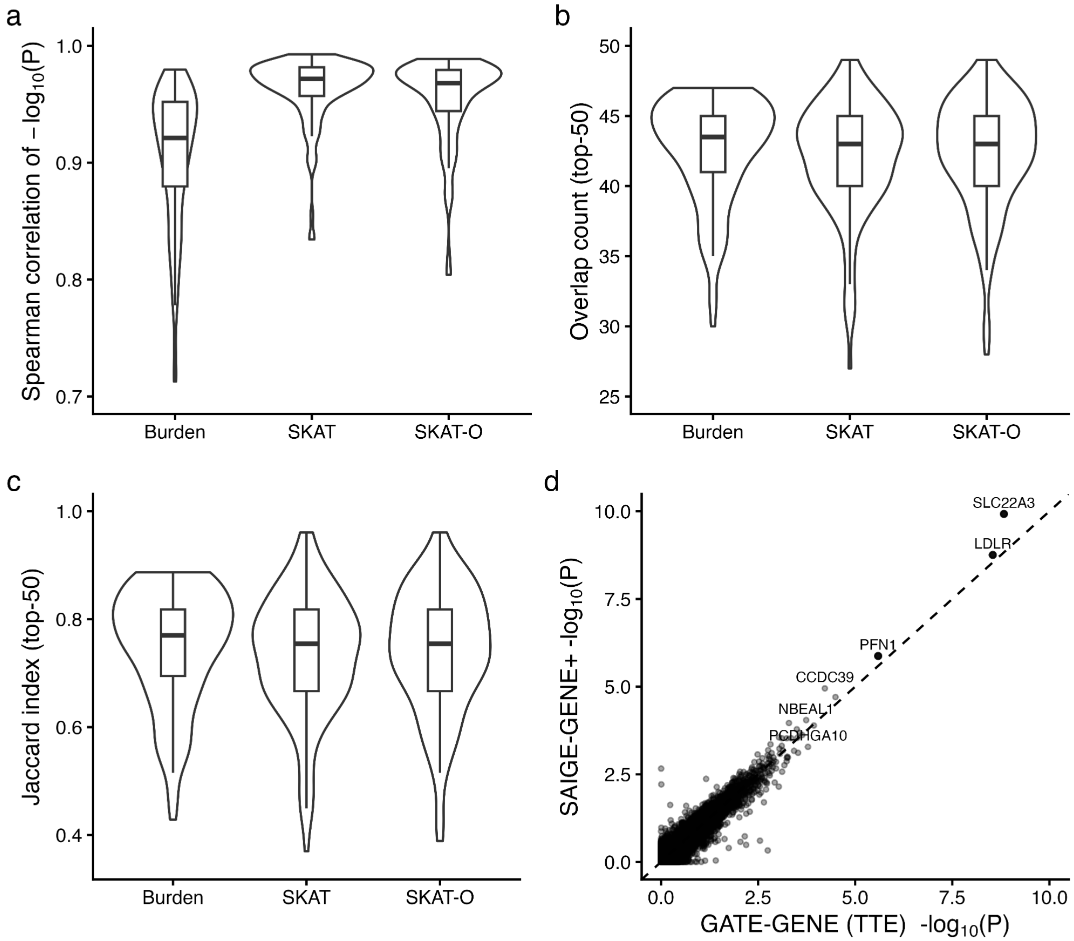


#### Supplementary Fig. 12 | Concordance of survival and incident case-control gene-based rare-variant associations in UK Biobank.

We compared gene-level association evidence from GATE-GENE time-to-event (TTE) analyses and matched incident case-control analyses using SAIGE-GENE+ across 120 onset phenotypes in UKB-EUR. a, Distribution across phenotypes of Spearman rank correlation between gene-level $-\log_{10}P$ values from TTE and incident case-control analyses for burden, SKAT and SKAT-O tests (Cauchy-combined across nine variant masks). b, Overlap of the top 50 ranked genes between the two frameworks for each phenotype. c, Jaccard index of the top 50 ranked genes between frameworks for each phenotype (intersection over union). Violin plots show the full distribution across phenotypes; center line, median; box, interquartile range; whiskers, 1.5× IQR. d, Example gene-level comparison for a representative phenotype (Coronary atherosclerosis), showing concordance of $-\log_{10}P$ between GATE-GENE and SAIGE-GENE+; dashed line denotes y=x. Selected genes are labeled for reference.

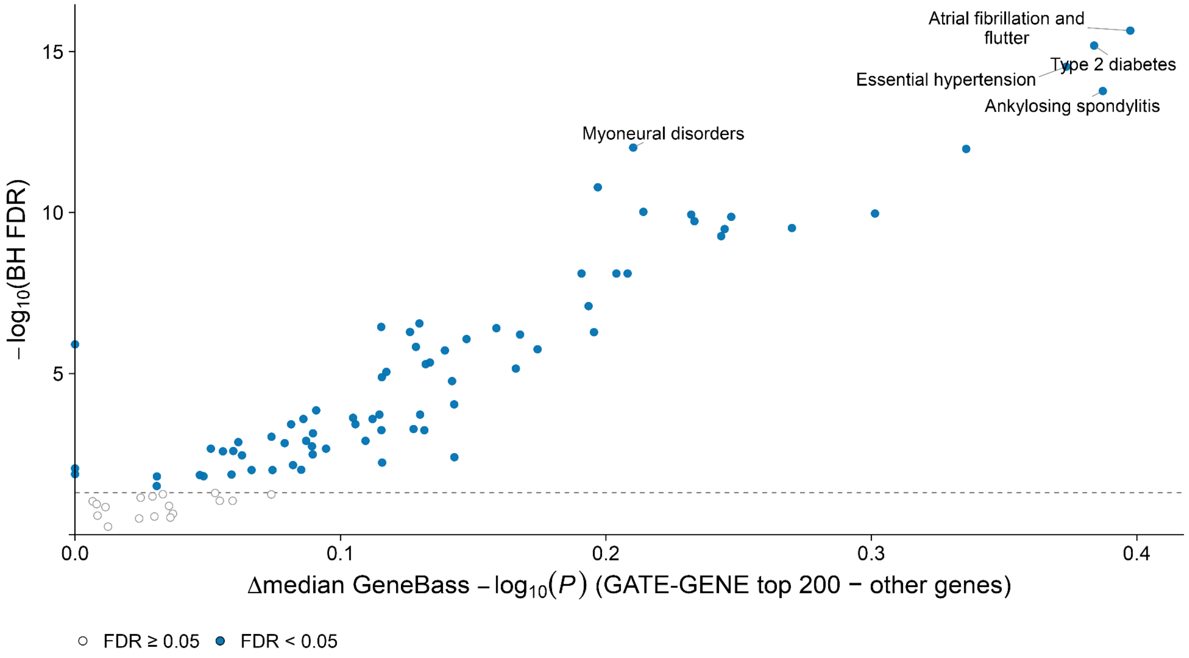


#### Supplementary Fig. 13 | Enrichment of GeneBass association evidence among GATE-GENE-prioritized genes

Each point represents one of 99 core phenotypes with matched GATE-GENE and GeneBass results. For each phenotype, analyses were restricted to the phenotype-specific intersection of genes with valid association results in both resources. The 200 genes with the strongest GATE-GENE evidence were compared with the remaining intersecting genes using a one-sided Wilcoxon rank-sum test. The x axis shows the difference in median GeneBass $-\log_{10}P$ between the GATE-GENE top-200 genes and the remaining genes; positive values indicate stronger GeneBass evidence among GATE-GENE-prioritized genes. The y axis shows $-\log_{10}P$ of the Benjamini-Hochberg FDR calculated across the 99 phenotypes. The horizontal dashed line denotes FDR = 0.05. Filled blue points indicate significant enrichment at FDR < 0.05, whereas open circles indicate FDR ≥ 0.05. GATE-GENE top-200 genes showed significantly stronger GeneBass evidence in 74 of 99 phenotypes (74.7%). Selected phenotypes with the strongest enrichment evidence are labeled.

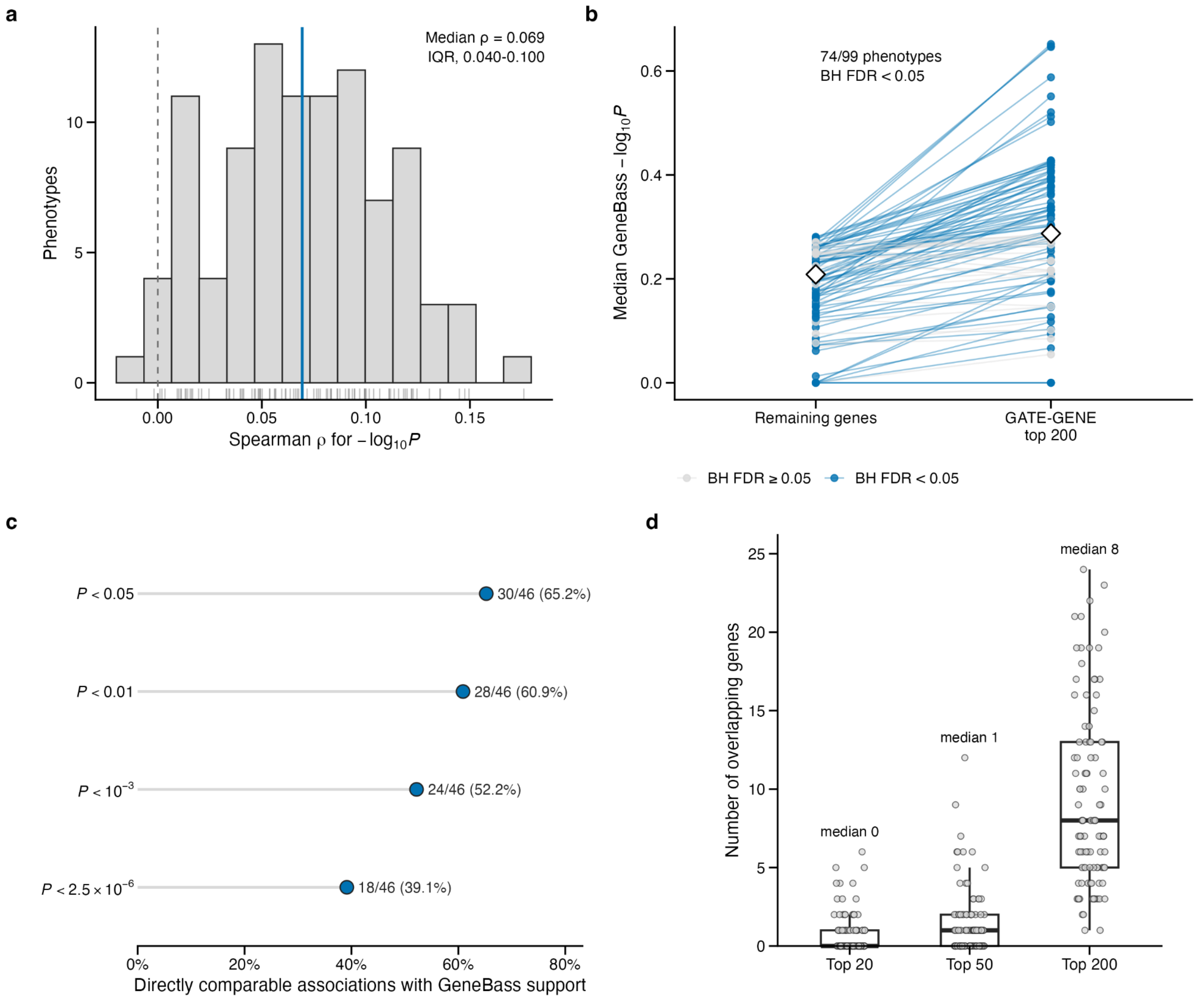


#### Supplementary Fig. 14 | Cross-analysis concordance of GATE-GENE onset associations with GeneBass prevalent case-control exome results.

a, Distribution of phenotype-specific Spearman correlations between GATE-GENE and GeneBass $-\log_{10}P$ values across 99 core phenotypes with valid gene-level results in both analyses. Correlations were calculated across the phenotype-specific intersection of genes tested in both resources. The solid blue line denotes the median correlation ($\rho = 0.069$), the dashed grey line denotes $\rho = 0$, and rug marks denote individual phenotypes. b, Paired median GeneBass $-\log_{10}P$ values for the 200 genes most strongly prioritized by GATE-GENE and the remaining genes in each phenotype-specific joint-gene universe. Lines connect values from the same phenotype; blue denotes phenotypes in which the GATE-GENE top-200 genes showed stronger GeneBass evidence at Benjamini-Hochberg FDR < 0.05 in a one-sided Wilcoxon rank-sum test, with correction across the 99 phenotypes. White diamonds denote medians across phenotypes. Significant enrichment was observed for 74 of 99 phenotypes. c, GeneBass support among the 46 directly comparable germline-likely, exome-wide significant GATE-GENE associations. Points and labels show the number and percentage of associations below each GeneBass *P*-value threshold. Thirty of 46 associations also remained significant after Benjamini-Hochberg correction across the directly comparable set. d, Number of genes shared between the top 20, top 50 and top 200 genes ranked by GATE-GENE and GeneBass across the 99 mapped phenotypes. Box plots show the median and interquartile range, with whiskers extending to 1.5 times the interquartile range; points denote individual phenotypes. Median overlaps are indicated above each distribution. Because both analyses use UK Biobank exome data, these comparisons provide cross-analysis support rather than independent replication.

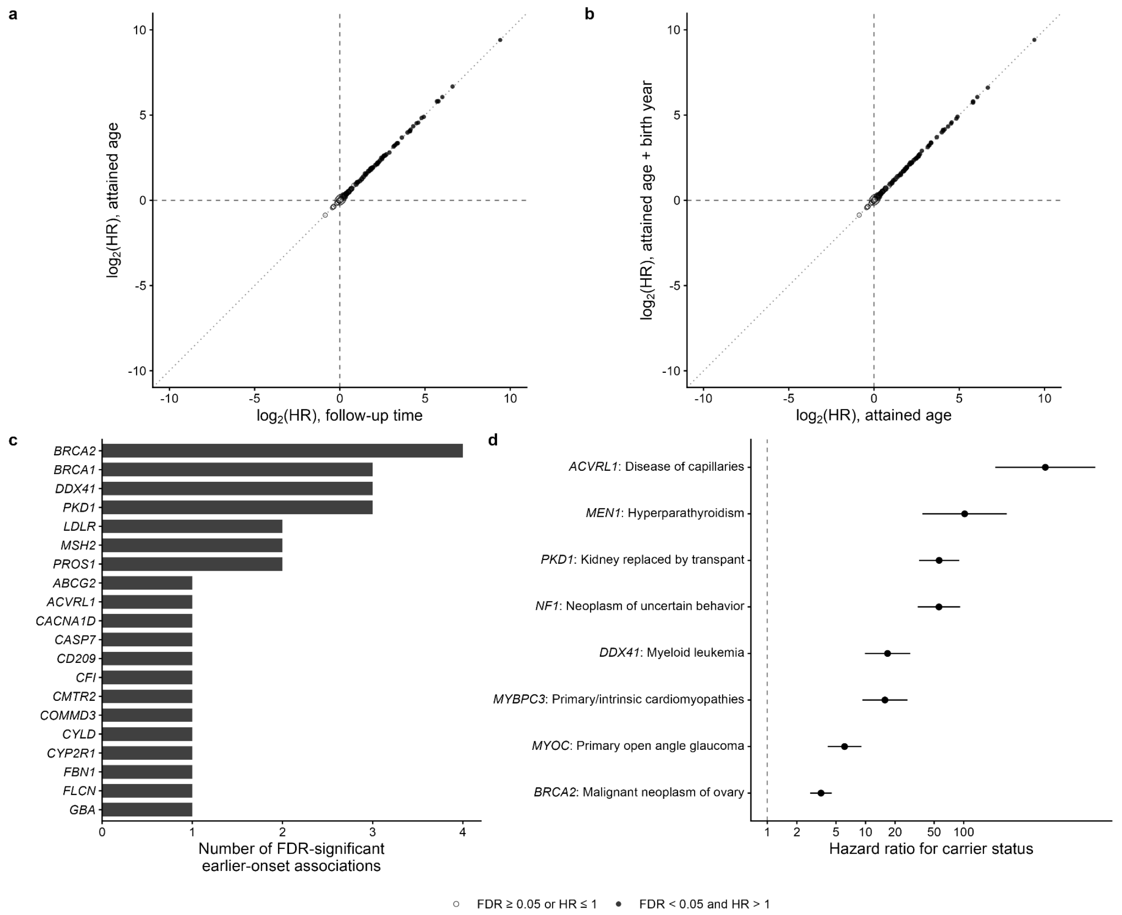


#### Supplementary Fig. 15 | Carrier-based timing analyses for core exome-wide significant gene-phenotype associations.

a, Concordance of carrier-associated hazard-ratio estimates between the discovery-aligned follow-up-time Cox model and the attained-age delayed-entry Cox model across analyzable core associations. Filled points indicate associations with FDR < 0.05 and burden-score HR > 1 in the attained-age model; open points indicate all other associations. Point size is proportional to the number of carrier events. The diagonal line denotes equality of hazard-ratio estimates. b, Concordance of carrier-associated hazard-ratio estimates between the primary attained-age model and a sensitivity model additionally adjusted for birth year. Point definitions are as in a. c, Genes ranked by the number of FDR-significant earlier-onset associations in the attained-age model among the 107 germline-likely core associations; up to 20 genes are shown. d, Attained-age hazard-ratio estimates and 95% confidence intervals for the strongest FDR-significant earlier-onset association for each of the eight most strongly associated distinct genes. The vertical dashed line denotes HR = 1. FDR values were calculated using the Benjamini-Hochberg procedure across the 107 core associations.

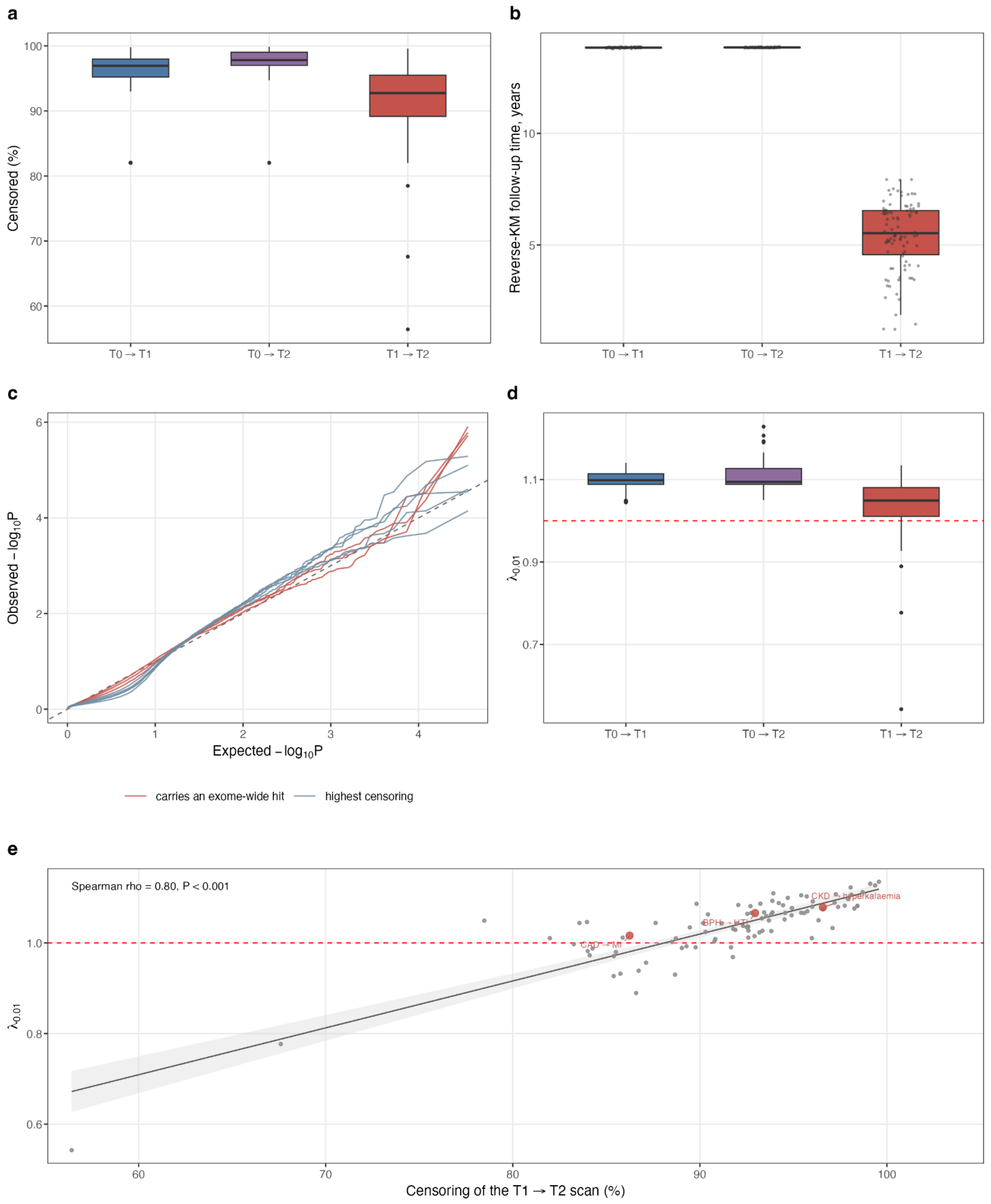


#### Supplementary Fig. 16 | Risk-set information and tail calibration of transition analyses in the UK Biobank.

a, Censoring by transition class across the 95 trajectories. b, Reverse Kaplan-Meier follow-up time by transition class; points are individual trajectories. Follow-up is shorter for T1→T2 than for both baseline-entry arms in all 95 trajectories, because progression time is clocked from the incident T1 date rather than from baseline; the two baseline arms are set by the administrative follow-up window (median 13.84 and 13.85 years, interquartile range 0.01 years each). c,

Quantile-quantile plots of gene-level *P* for the three progression analyses carrying an exome-wide association and the five most heavily censored progression analyses; dashed line, null expectation. d, Calibration in the 1st percentile tail of the gene-level P distribution ($\lambda_{0.01}$) summarised by the genomic-control factor and transition class; dashed line, null expectation of 1. e, $\lambda_{0.01}$ against censoring for the 95 T1→T2 analyses; analyses carrying an exome-wide progression association are highlighted and labelled; grey line and shaded band, linear fit with 95% confidence interval.

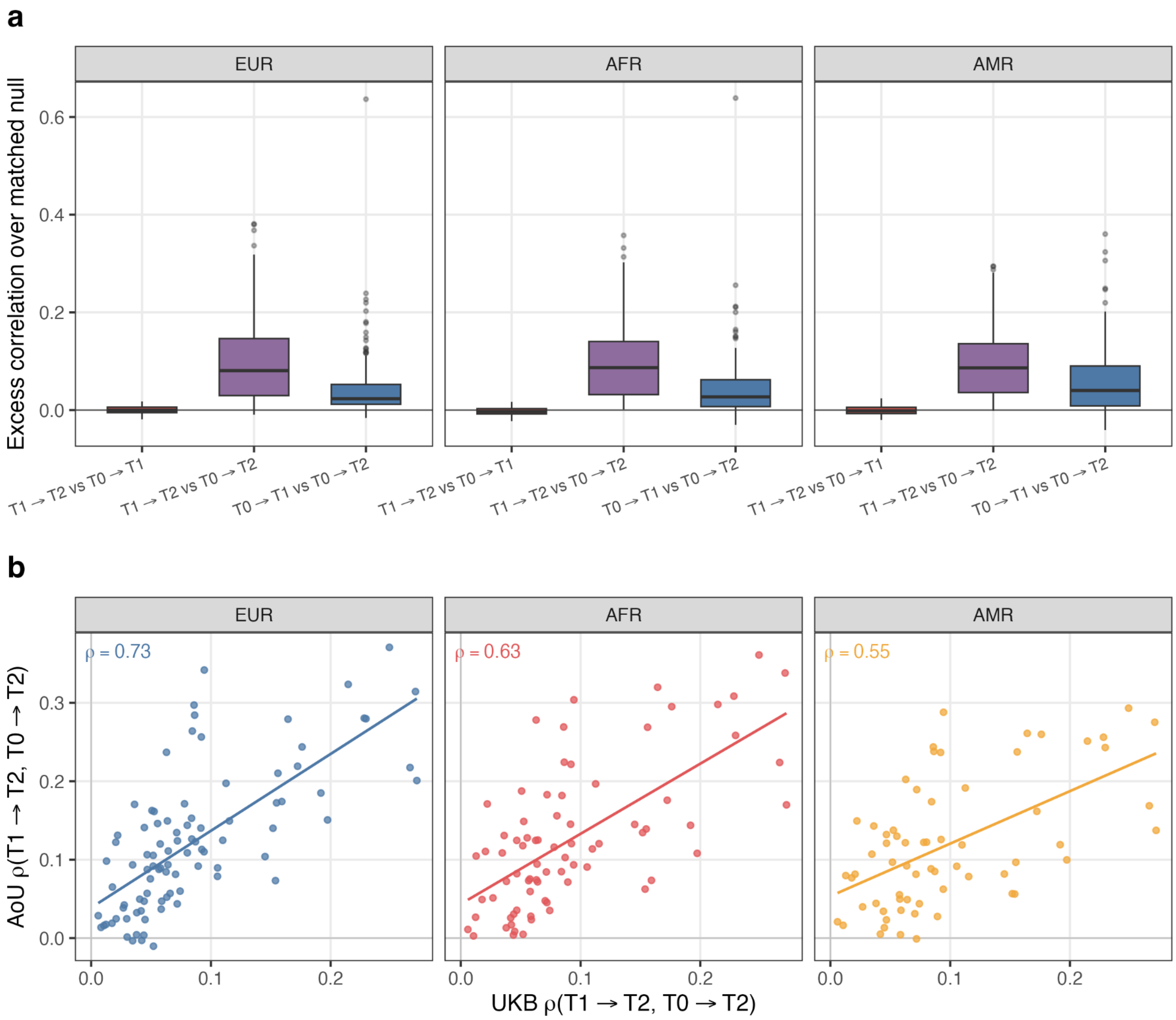


Supplementary Fig. 17 | Cross-transition concordance relative to the composition-matched null in All of Us, and trajectory-level agreement with UK Biobank.

a, Excess gene-level concordance over the composition- and event-matched null in All of Us, by transition-pair class and ancestry (179 EUR, 134 AFR and 116 AMR trajectories with all three transition arms fitted). Values are the observed Spearman $\rho$ between gene-level $-\log_{10} P$ vectors of the two transitions minus the median ρ of disjoint transition pairs matched on both minimum event count and transition composition; zero corresponds to the matched null. Null cells with fewer than 30 disjoint pairs were excluded. Boxes show the median and interquartile range; whiskers extend to 1.5× the interquartile range; points beyond are shown individually. b, Per-trajectory architecture agreement across biobanks: UK Biobank versus All of Us $\rho$(T1→T2, T0→T2) for each trajectory analysed in both cohorts (n = 91 EUR, 76 AFR and 65 AMR). Lines are linear fits; annotated values are Spearman correlations ($\rho$ = 0.73, 0.63 and 0.55; $P = 3.6 \times 10^{-16}$, $1.3 \times 10^{-9}$ and $2.7 \times 10^{-6}$).

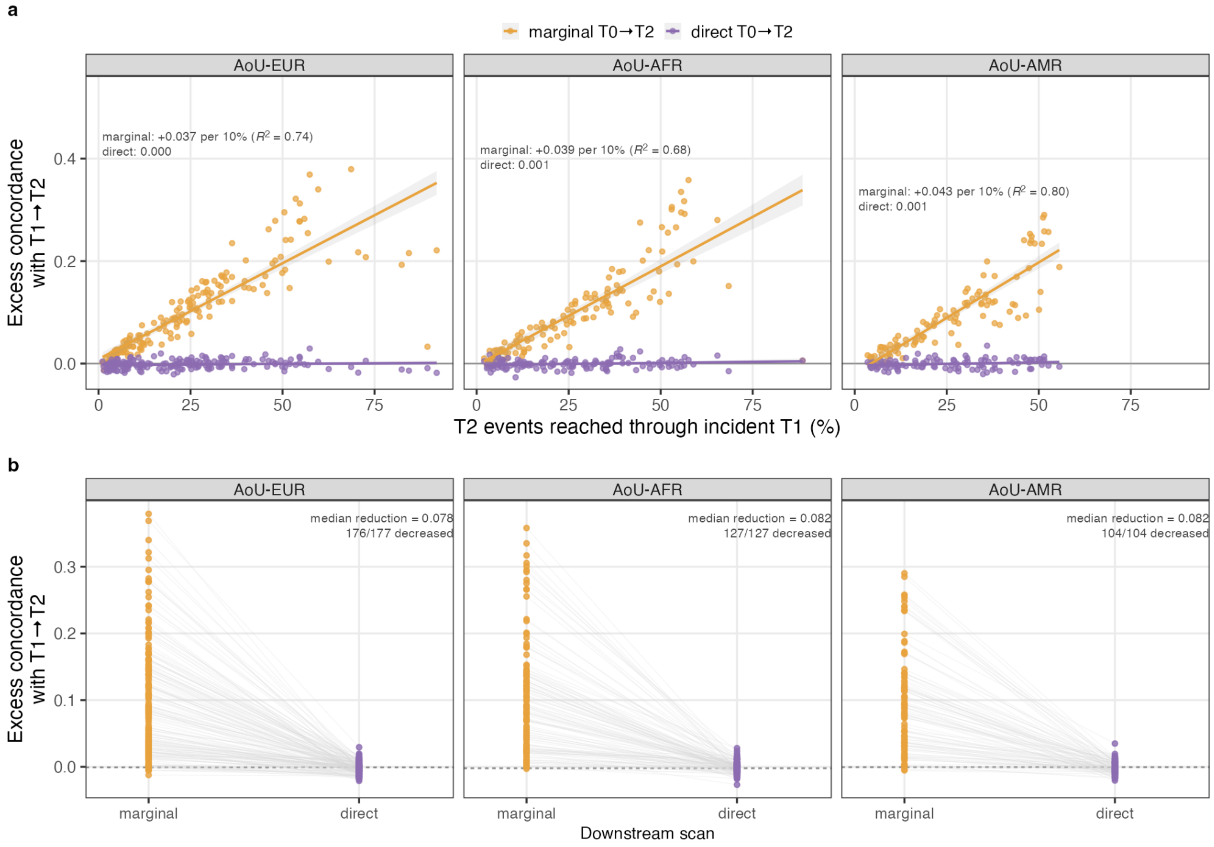


#### Supplementary Fig. 18 | T2-event overlap gradient and marginal-to-direct attenuation in All of Us, per ancestry.

a, Progression-downstream concordance as a function of T2-event overlap in All of Us (AoU) European (EUR), African (AFR) and Admixed-American (AMR) ancestry groups. The x-axis shows the fraction of downstream T2 events reached through incident T1, and the y axis shows excess gene-level concordance with the corresponding T1→T2 progression relative to matched-null expectation. Orange denotes marginal T0→T2 analyses, which follow participants to T2 irrespective of whether T1 occurs first; purple denotes direct T0→T2 transitions, in which participants exit the direct-transition risk set at incident T1. Points represent individual trajectories and lines show ordinary least-squares fits with 95% confidence intervals. b, Within-trajectory attenuation of progression-downstream concordance after replacing marginal with direct T0→T2 analyses. Paired estimates are shown for 177 EUR, 127 AFR and 104 AMR trajectories; grey lines connect the same trajectory. The dashed horizontal line denotes zero excess concordance

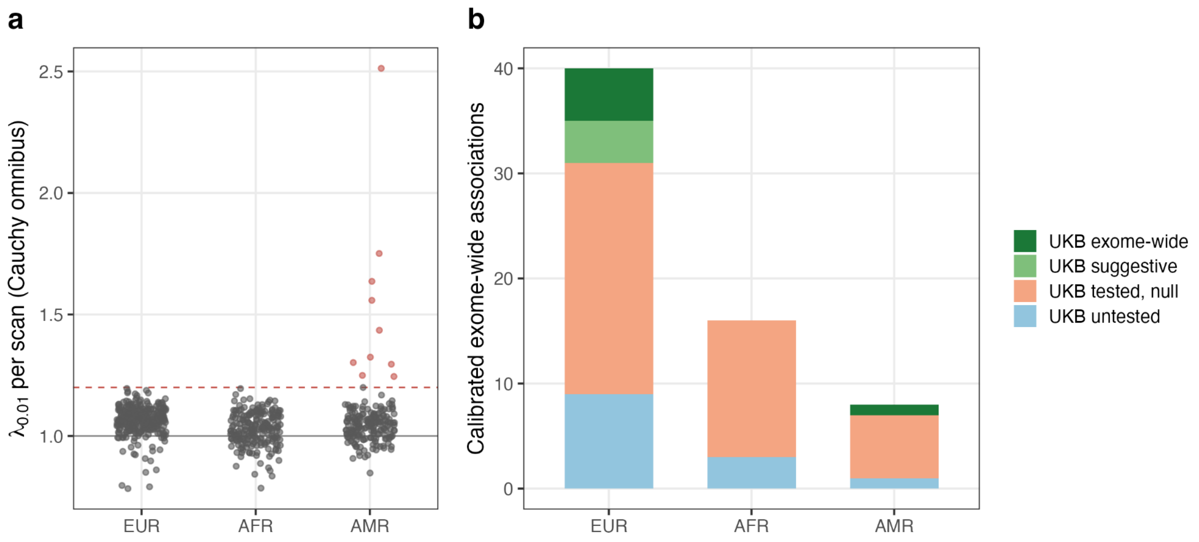


#### Supplementary Fig. 19 | Native All of Us calibration and discovery classification.

a, Tail-based genomic-control factor $\lambda_{0.01}$ of the per-gene Cauchy omnibus for every analysed transition (279 EUR, 213 AFR and 188 AMR), by ancestry. $\lambda_{0.01}$ compares the observed and expected statistic at the 1st percentile of the gene-level *P* distribution; the solid line marks 1 and the dashed line the exclusion threshold of 1.2. All EUR and AFR analyses were calibrated; eight AMR transitions exceeded the threshold (red) and their apparent associations were excluded from all discovery counts. b, Calibrated native exome-wide associations (Cauchy omnibus $P < 2.5 \times 10^{-6}$; 40 EUR, 16 AFR and 8 AMR) per ancestry, classified against UK Biobank as exome-wide, suggestive ($P < 1 \times 10^{-4}$), tested without signal, or untested.

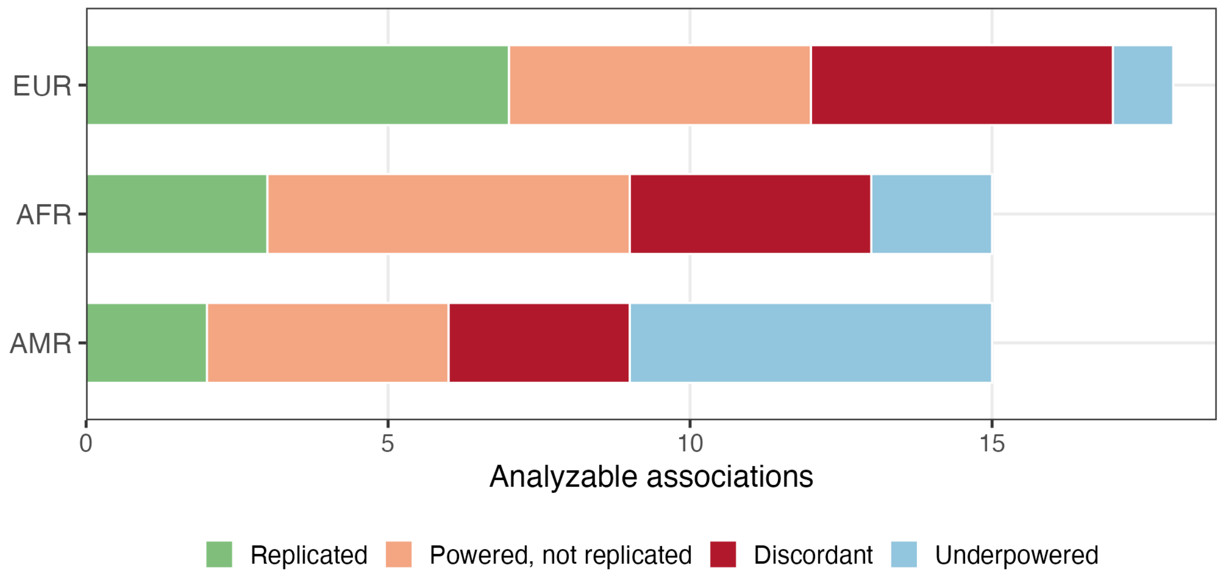


**Supplementary Fig. 20 | Power-based classification of replication outcomes.**

Analyzable UK Biobank exome-wide associations in each All of Us ancestry (18 EUR, 15 AFR and 15 AMR), classified as replicated (primary criterion); powered but not replicated; directionally discordant; or directionally consistent but underpowered. “powered” denotes estimated power ≥ 80% and “discordant” requires estimated power ≥ 20% with an opposite effect direction. Most non-replications occurred despite adequate power, implicating cohort and phenotype heterogeneity rather than sample size alone.
